# Probabilistic inference of the genetic architecture underlying functional enrichment of complex traits

**DOI:** 10.1101/2020.09.04.20188433

**Authors:** Marion Patxot, Daniel Trejo Banos, Athanasios Kousathanas, Etienne J. Orliac, Sven E. Ojavee, Gerhard Moser, Alexander Holloway, Julia Sidorenko, Zoltan Kutalik, Reedik Mägi, Peter M. Visscher, Lars Rönnegård, Matthew R. Robinson

## Abstract

Due to the complexity of linkage disequilibrium (LD) and gene regulation, understanding the genetic basis of common complex traits remains a major challenge. We develop a Bayesian model (BayesRR-RC) implemented in a hybrid-parallel algorithm that scales to whole-genome sequence data on many hundreds of thousands of individuals, taking 22 seconds per iteration to estimate the inclusion probabilities and effect sizes of 8.4 million markers and 78 SNP-heritability parameters in the UK Biobank. We show in theory and simulation that BayesRR-RC provides robust variance component and enrichment estimates, improved marker discovery and effect estimates over mixed-linear model association approaches, and accurate genomic prediction. Of the genetic variation captured for height, body mass index, cardiovascular disease, and type-2 diabetes in the UK Biobank, only ≤ 10% is attributable to proximal regulatory regions within 10kb upstream of genes, while 12-25% is attributed to coding regions, 32-44% to intronic regions, and 22-28% to distal 10-500kb upstream regions. ≥ 60% of the variance contributed by these exonic, intronic and distal 10-500kb regions is underlain by many thousands of common variants, which on average have larger effect sizes than for other annotation groups. Up to 24% of all cis and coding regions of each chromosome are associated with each trait, with over 3,100 independent exonic and intronic regions and over 5,400 independent regulatory regions having ≥ 95% probability of contributing ≥ 0.001% to the genetic variance of these four traits. Thus, these quantitative and disease traits are truly complex. The BayesRR-RC prior gives robust model performance across the data analysed, providing an alternative to current approaches.

---

As whole-genomes are collected for hundreds of thousands of individuals, we require regression methods that are not only computationally efficient, but which also provide improved inference. Rather than relying on subsets of the SNPs, methods should fully utilize the data, exploiting computational power to facilitate discovery of additional genomic regions, to improve understanding of the genomic architecture of common disease, and to provide more informative genomic prediction.

Recent studies [1–4] highlight the importance of accounting for minor allele frequency (MAF) and LD structure of the genomic data when estimating the proportion of phenotypic variance attributable to different categories of genetic markers (the SNP-heritability,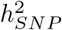 of a genomic region). Assessment of the relative contribution of different genomic regions is currently made assuming that markers within a category all contribute to the variance, with enrichment defined as the estimated share of the variance explained divided by its expected share [5, 6]. However ideally, the estimated distribution of marker effects for each category would be directly obtained, accounting for MAF and LD structure and allowing for some of the marker effects to be zero, as this would yield a better understanding of the polygenicity of genomic effects across different genomic regions.

Current mixed-linear association models such as those implemented in the software fastGWA [7], boltLMM [8], and REGENIE [9], use a two-step approach, first estimating the variance contributed by the SNP markers without the use of MAF-LD-annotation information, which is then used to estimate the marker effect sizes in a second step, essentially assuming effects in the model come from a single distribution [7, 8, 10]. Summary statistic approaches such as LDSC [11] and SumHer [6], then use these baseline effect estimates coupled with independent LD reference panels to then alter the weightings of the marker effects allowing for annotation differences, showing improved genomic prediction. However, no model currently provides joint estimates of the marker effects and tests for association whilst accounting for effect size differences across MAF, LD, or annotation groups.

Here, we outline the fastest Bayesian penalised regression model to date, with a hybrid-parallel algorithm for analysing large-scale genomic data that: (i) provides unbiased MAF-LD annotation-specific genetic effect size estimates and 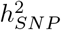 of different annotations in a single step, allowing for a contrasting of the genetic architectures of complex traits under a flexible prior formulation, (ii) yields the probability that each marker, genomic region, annotation, gene-coding region, or SNP is associated with a phenotype, alongside the proportion of phenotypic variation contributed by each describing the *gene* architecture of complex traits, (iii) gives a posterior predictive distribution for each individual at each genomic region.

## A Bayesian model for large-scale genomic data

The model we derive (which we call BayesRR-RC) is based on grouped effects with mixture priors, improving on the formulations of [12, 13] and [14]. Like these former methods, we consider a spike probability at zero (Dirac delta function), and a scale mixture of Gaussian distributions as a slab probability density. Unlike these models, we have genetic markers grouped into MAF-LD-annotation specific sets, with independent hyper-parameters for the phenotypic variance attributable to each group. Assuming *N* individuals and *p* genetic markers, our model of an observed phenotype vector **y** is:

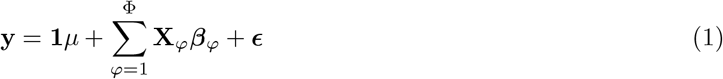

where there is a single intercept term **1***μ* and a single error term, a vector (*N* × 1) of residuals *ϵ*, with 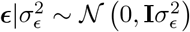. An *N* by *p* matrix of single nucleotide polymorphism (SNP) genetic markers, centered and scaled to unit variance, which we denote as **X**_*ϕ*_. The effects are allocated into groups (1, …, Φ). Each group has a set of model parameters 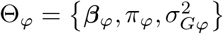, with ***β***_*ϕ*_ as a *p*_*ϕ*_ × 1 vector of partial regression coefficients, where 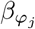 is the effect of a 1 SD change in the *j*^*th*^ covariate within the *ϕ*^*th*^ group. The spike and slab prior, contains what is called a Dirac spike [15, 16] for ***β***_*ϕ*_, which induces sparsity in the model through a Dirac-delta at zero, excluding variables from the model by setting their coefficients to zero. A finite scale mixture of normal distributions centered at zero constitute the slab component. The slab shrinks the non-zero coefficients towards zero according to the slab’s width, and by having a scale mixture of Gaussians, the distribution has heavier tails and can accommodate big and small effects [17]. Therefore, each 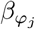 is distributed according to:

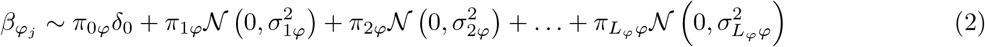

where for each SNP marker group 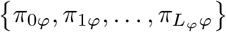 are the mixture proportions and 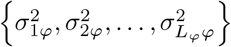 are the mixture-specific variances proportional to

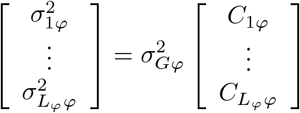

with 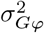 the phenotypic variance associated with the SNPs in group *φ*, which, like all the other parameters, is estimated directly from the data. Thus, related approaches of BayesRC and BayesRS that are heavily utilized in animal and plant breeding [18, 19] are extended as the mixture proportions, the variance explained by the SNP markers, and the mixture constants are all unique and independent across SNP marker groups. This enables estimation of the amount of phenotypic variance attributable to the group-specific effects, and differences in the underlying distribution of the ***β***_*φ*_ effects among MAF-LD-annotation groups, with different degrees of sparsity.

### Simulation study

We wished to highlight the performance of BayesRR-RC for the following tasks: (i) estimation of the phenotypic variance attributable to genetic markers, both genome-wide, and for segments of the genome; (ii) “discovery” of associated genomic regions and identification of candidate SNP groups; and (iii) phenotypic prediction. We developed a large-scale simulation study, randomly selecting 40,000 unrelated individuals and all 596,741 imputed (version 3) genetic markers from chromosomes 19 through 22 from the UK Biobank. Using this data, we simulated a wide-range of different possible underlying genetic effect size distributions as described in Table 1 and Methods. This range covers generative genetic models discussed in the literature and provides data that both fit and violate the assumptions of the range of variance component models, both individual-level and summary statistic approaches, that are currently applied in the literature for estimation of the variance attributable to the SNP markers, for testing association of genetic markers with phenotypes genome-wide, and for genomic prediction.

**Table 1.** The generative genetic models used in the simulation study. Imputed SNP marker data from chromosomes 19, 20, 21 and 22 of 40,000 randomly selected UK Biobank participants were selected, giving 596,741 markers in total. Marker effects were simulated according to the 20 generative models in two ways: (i) a single distribution of marker effects, and (ii) 13 distributions of marker effects for 13 different genomic annotation groups with different proportions of SNP heritability 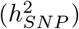 explained for exonic variants 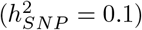, intronic variants 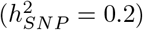, 1kb promotor variants 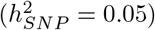, 1-10kb enhancer variants (0.025), 1-10kb transcription factor binding sites 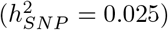, 1-10kb other variants 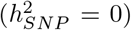, 10-500kb enhancers 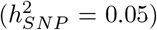, 10-500kb transcription factor binding sites 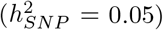, 10-500kb other variants 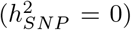, 500kb-1Mb enhancers 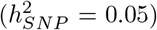, 500kb-1Mb transcription factor binding sites 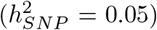, 500kb-1Mb other variants 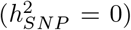, and other non-annotated SNPs 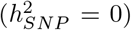. 10 simulation replicates were created for both (i) and (ii) giving a total set of 400 simulated phenotypes.

| generative model | causal variant allocation | causal variants | effect size (b), LD (w), MAF (p) relationship |
| --- | --- | --- | --- |
| 1 | highest MAF per LD block | 10,000 | $b \propto N(0, w^{-0.25}[p(1-p)]^{-0.25})$ |
| 2 | highest MAF per LD block | 10,000 | $b \propto N(0, w^{0.25}[p(1-p)]^{-0.25})$ |
| 3 | highest MAF per LD block | 10,000 | $b \propto N(0, w^{-0.25}[p(1-p)]^{0.75})$ |
| 4 | highest MAF per LD block | 10,000 | $b \propto N(0, w^{0.25}[p(1-p)]^{0.75})$ |
| 5 | highest MAF per LD block | 10,000 | $b \propto N(0, w^0[p(1-p)]^0)$ |
| 6 | highest MAF per LD block | 5,000 | $b \propto N(0, w^{-0.25}[p(1-p)]^{-0.25})$ |
| 7 | highest MAF per LD block | 5,000 | $b \propto N(0, w^{0.25}[p(1-p)]^{-0.25})$ |
| 8 | highest MAF per LD block | 5,000 | $b \propto N(0, w^{-0.25}[p(1-p)]^{0.75})$ |
| 9 | highest MAF per LD block | 5,000 | $b \propto N(0, w^{0.25}[p(1-p)]^{0.75})$ |
| 10 | highest MAF per LD block | 5,000 | $b \propto N(0, w^0[p(1-p)]^0)$ |
| 11 | random | 10,000 | $b \propto N(0, w^{-0.25}[p(1-p)]^{-0.25})$ |
| 12 | random | 10,000 | $b \propto N(0, w^{0.25}[p(1-p)]^{-0.25})$ |
| 13 | random | 10,000 | $b \propto N(0, w^{-0.25}[p(1-p)]^{0.75})$ |
| 14 | random | 10,000 | $b \propto N(0, w^{0.25}[p(1-p)]^{0.75})$ |
| 15 | random | 10,000 | $b \propto N(0, w^0[p(1-p)]^0)$ |
| 16 | random | 5,000 | $b \propto N(0, w^{-0.25}[p(1-p)]^{-0.25})$ |
| 17 | random | 5,000 | $b \propto N(0, w^{0.25}[p(1-p)]^{-0.25})$ |
| 18 | random | 5,000 | $b \propto N(0, w^{-0.25}[p(1-p)]^{0.75})$ |
| 19 | random | 5,000 | $b \propto N(0, w^{0.25}[p(1-p)]^{0.75})$ |
| 20 | random | 5,000 | $b \propto N(0, w^0[p(1-p)]^0)$ |

Throughout the simulation study and UK Biobank data analysis, we apply our BayesRR-RC model using 78 MAF-LD-annotation SNP marker groups. SNPs are partitioned into 7 location annotations preferentially to coding (exonic) regions first, then to intronic regions, then to 1kb upstream regions, then to 1-10kb regions, then to 10-500kb regions, then to 500-1Mb regions. Remaining SNPs were grouped in a category labelled “others” and also included in the model so that variance is partitioned relative to these also. Thus, we assigned SNPs to their closest upstream region, for example if a SNP is 1kb upstream of gene X, but also 10-500kb upstream of gene Y and 5kb downstream for gene Z, then it was assigned to be a 1kb region SNP. This ensures that SNPs 10-500kb and 500kb-1Mb upstream are distal from any known gene. We further partition upstream regions to experimentally validated promoters, transcription factor binding sites (tfbs) and enhancers (enh) using the HACER, snp2tfbs databases (see Code Availability). All SNP markers assigned to 1kb regions map to promoters; 1-10kb SNPs, 10-500kb SNPs, 500kb-1Mb SNPs are then split into enh, tfbs and others (unmapped SNPs) extending the model to 13 annotation groups. Within each of these annotations, we have three minor allele frequency groups (MAF ≤ 0.01, 0.01<MAF ≤ 0.05, and MAF>0.05), and then each MAF group is further split into 2 based on median LD score. This gives 78 non-overlapping groups for which our BayesRR-RC model jointly estimates the phenotypic variation attributable to, and the SNP marker effects within, each group. For each of the 78 groups, SNPs were modelled using five mixture groups with variance equal to the phenotypic variance attributable to the group multiplied by constants (mixture 0 = 0, mixture 1 = 0.0001, 2 = 0.001, 3 = 0.01, 4 = 0.1). In the following sections, we explore the model performance of BayesRR-RC and compare to existing approaches across the range of different simulation scenarios described in Table 1.

### Variance component estimation

We compare our BayesRR-RC model to the following statistical models: (i) a restricted maximum likelihood (REML) model implemented in the software GCTA with a single relationship matrix providing an estimate of the variance attributable to SNPs genome-wide, (ii) a REML model implemented in the software BoltREML [20] with the same 78 MAF-LD-annotation groups used for BayesRR-RC enabling a direct comparison, (iii) a Haseman-Elston (HE) regression using the same 78 group model implemented in the software RHEmc [21],(i) summary statistic linkage disequilibrium score regression (LDSC) [11], with LD scores calculated using the same data, and the same 78 non-overlapping annotations in a 78 component LDSC annotation model, and and summary statistic SumHer [6] with the same 78 non-overlapping annotations. Across the range of generative genetic models, both BayesR and BayesRR-RC estimated the variance attributable to SNP markers with high accuracy, with the distribution of BayesRR-RC model posterior mean estimates across simulation replicates, calculated as a % difference from the simulated value, containing zero across all simulation scenarios (Figure 1a). We find that BayesRR-RC estimates the variance attributable to different genomic regions on the correct scale, with lower RMSE than other approaches (Figure 1b), and that this results in the estimated average effect size for each annotation group having high correlation with the simulated value (Figure 1c). This indicates that BayesRR-RC provides accurate estimates of the underlying effect size distribution for different genomic groups, across a wide range of different underlying generative genetic models.

**Figure 1.**
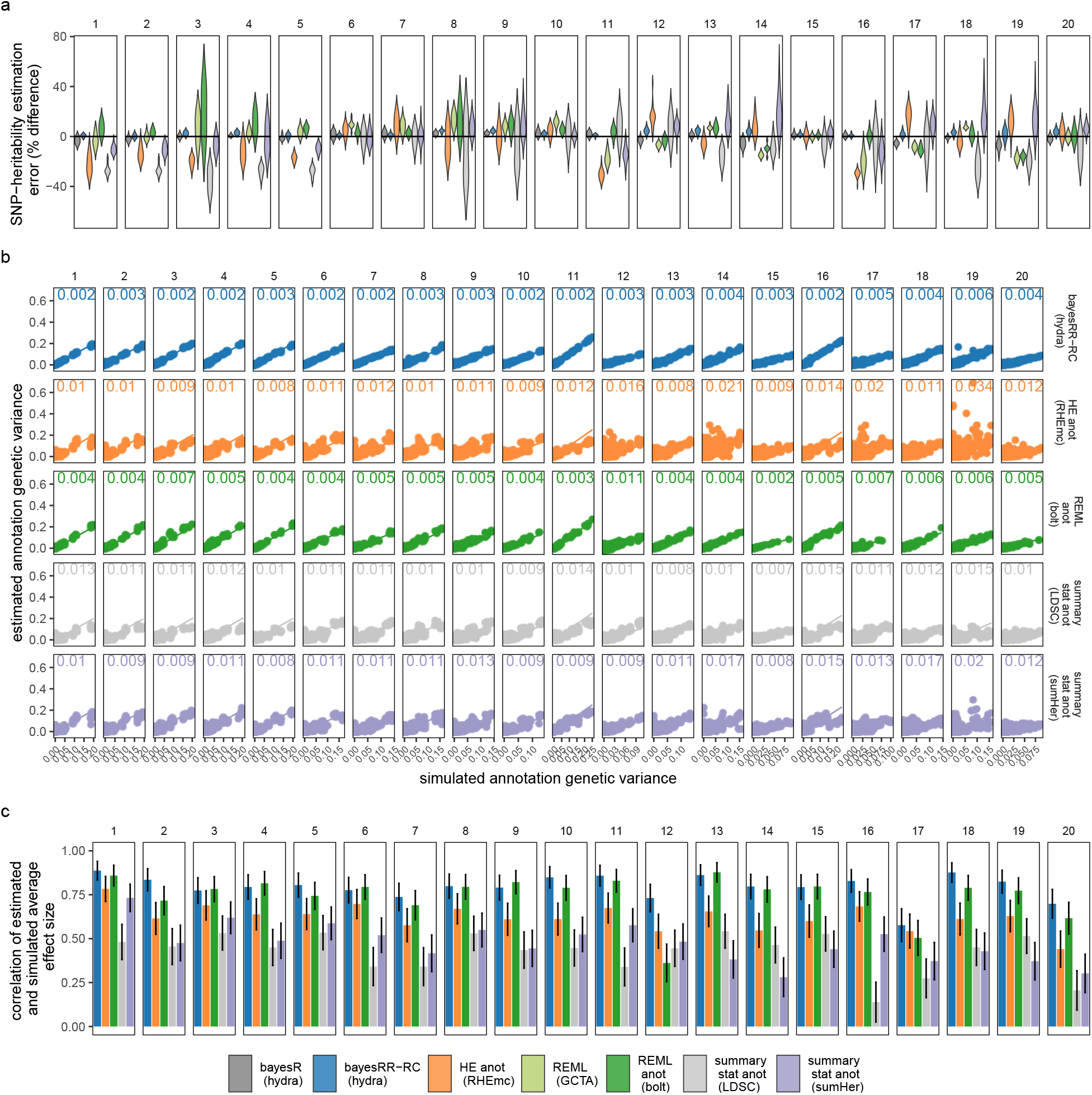
Simulation study of the variance component estimation performance of BayesRR-RC. (a) Violin-plot of the genome-wide SNP-heritability estimates as a percentage difference from the simulated value for 40 replicates, for each of 20 different generative genetic models described in Table 1. For each generative genetic model we compare seven different statistical models: a mixture of regression model with a single global variance component known as “bayesR” implemented in our hydra software (bayesR hydra), the mixture of regression model with multiple group-specific variance components described in this work implemented in our hydra software (bayesRR-RC hydra), Haseman-Elston regression with annotation-specific relationship matrices implemented in the RHEmc software (HE anot RHEmc), a single component REML model implemented in the software GCTA (REML GCTA), a multiple group-specific variance component REML model implemented in the software bolt (REML anot bolt), and two annotation summary statistic models implemented in the software LDSC and sumHer. (b) The estimated genetic variance for each of 13 genetic annotation groups plotted against the simulated genetic variance for the five statistical approaches which enable annotation-specific estimation. Root mean square error values are shown alongside lines representing the 1:1 relationship with the simulated value. (c) Bar-plots of the correlation of the estimated and simulated average effect size of each annotation. Error bars give the SD.

For estimation of the variance attributable to different genomic regions, we find that all statistical models other than BayesRR-RC are sensitive to the underlying generative genetic model, with no other approach providing consistent estimates across the 20 generative genetic models (Figure 1a). The individual-level BoltREML model estimates the variance attributable to different genomic regions on the correct scale, with low RMSE similar to BayesRR-RC (Figure 1b), but with slightly lower correlation of the estimated and simulated average effect sizes in many simulation scenarios (Figure 1c). Generally, summary statistic approaches perform poorly for both variance component estimation (Figure 1b) and quantification of enrichment as compared to individual-level methods, often even incorrectly selecting the group of highest average effect size (Figure 1c). While comparisons like this of different approaches have been made under different simulation scenarios, direct comparisons of individual-level and summary statistic approaches are lacking, and more importantly we have limited understanding of why approaches differ. We show in theory (see Methods Eq.23) and in additional simulations (see Methods, Figure S1 to Figure S3) the importance of our model formulation for accurate estimation of the variance components and the SNP regression coefficients, which we explore in more detail below.

### Estimation of SNP marker effects and discovery

While direct comparisons of frequentist and Bayesian approaches are difficult and often ill advised, we wished to show that BayesRR-RC provides accurate effect size estimation in the presence of LD and that this leads to more ‘discovery’ of associated variants. Here, we provide three simple comparable metrics to assess model performance of BayesRR-RC against frequentist mixed linear association models (MLMA) applied as two-stage approaches, where either the SNP is fitted twice as it is included in both the fixed and random terms (MLMAi implemented in GCTA), or the SNP to be tested as fixed is removed from the random term alongside those on the same chromosome (MLMA implemented in BoltLMM and Regenie).

First, we calculated z-scores of the marker effect estimates from their true simulated value. As MLMA approaches estimate marker effects one-at-a-time, we calculated the z-score of the estimate from the true simulated value for the causal variants in each simulation replicate, across generative genetic models. For the Bayesian methods, at any one iteration LD among the markers is controlled for (see Eq. 35 where we describe these properties). However across iterations as the chain mixes, markers in LD will enter and leave the model, with their posterior inclusion probabilities reflecting their association with the trait (Eq. 35). Thus, we summed the squared regression coefficient estimates of SNPs in the model at each iteration for each LD block (markers in LD *R*^2^ ≥ 0.1 within 1MB) of each simulation replicate, took the posterior mean across iterations, and then calculated the z-score of the estimate from the simulated value. This metric provides an assessment of the ability of BayesRR-RC to accurately estimate the contribution of a genomic region to the phenotypic variance as compared to MLMA approaches. We find that the z-scores of the estimated BayesRR-RC effects are generally stable across generative genetic models and comparable to those obtained from BayesR but with slightly elevated variance in many cases as the model is less sparse (Figure 2a). We find that SNP effect size estimates from MLMA models have higher estimation error, especially when the causal variant is rare, or in high-LD with many other SNPs (Figure 2a). MLMAi models show lower estimation error than MLMA approaches, likely as they control for both distant and local LD (Figure 2a). We show in theory (see Methods) and in additional simulations that penalized regression or mixed linear model approaches can inaccurately estimate the effects of highly correlated variants (under multicollinearity, Figure S1, Figure S2, and Methods Eq.23) due to the assumption made by these models that effects come from a single Gaussian distribution. We show how SNP marker regression coefficient estimation accuracy improves when using MAF-LD specific shrinkage in sparser BayesR and BayesRR-RC models, because estimated common variant effects in high LD are shrunk to a greater degree, alleviating the influence of multicollinearity (Figure S1, Figure S2).

**Figure 2.**
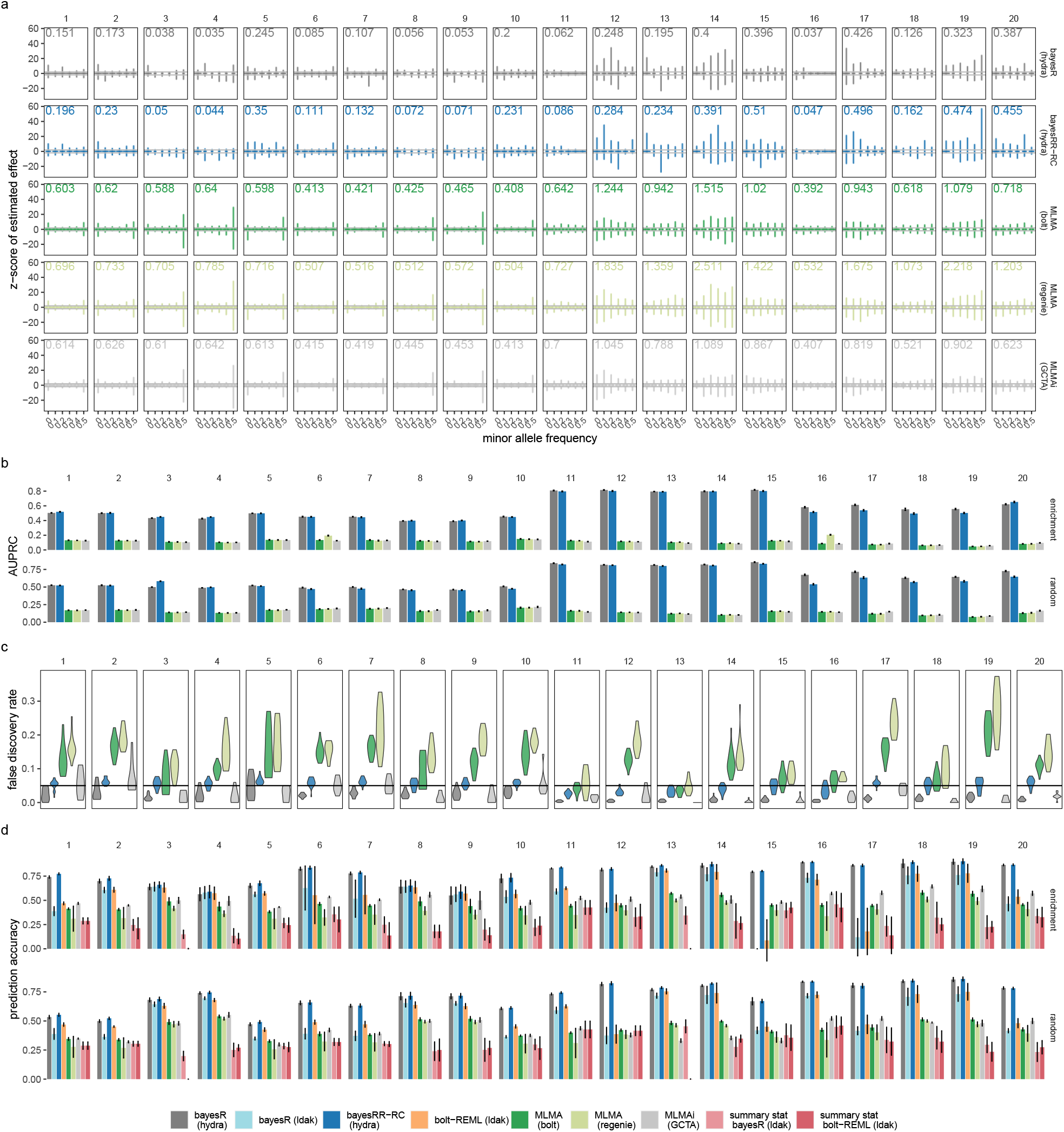
Simulation study of the effect size estimation and prediction performance of BayesRR-RC. (a) For 20 different generative genetic models we compare model performance of Bayesian models (bayesR and bayesRR-RC) and frequentist mixed-linear association models (MLMA) where the genetic marker tested for association is removed from the relationship matrix (implemented in software bolt and regenie), or fitted both as fixed and random (MLMi implemented in the software GCTA). For bayesR and bayesRR-RC we summed the squared regression coefficient estimates of all SNPs in LD with each causal variant (markers in LD *R*^2^ ≥ 0.1), took the posterior mean, and calculated the z-score from the simulated value. For the MLMA approaches, we calculated the z-score of the causal marker estimate from the simulated value. Violin plots for groups of minor allele frequency of the causal variant are shown, with values giving the variance. (b) Area-under the precision-recall curve (AUPRC). For Bayesian methods we use our PPWV metric (see Methods), with true positives defined as LD blocks that contain a causal variant and false positives defined as LD blocks that did not contain a causal variant. For MLMA methods we LD-clumped the results (LD *R*^2^ ≥ 0.01) using the p-value of the chi-squared statistics. Markers in *R*^2^ ≥ 0.01 with simulated causal variants were defined as true positives and those not in LD *R*^2^ ≥ 0.01 as false positives. (c) False discovery rate (FDR), with the line giving the 5% threshold. For the MLMA methods, FDR was calculated as the proportion of LD independent SNPs with p-value ≤5×10^−8^ that were not in LD *R*^2^ ≥ 0.01 with causal variants. For the Bayesian methods, we defined FDR as the proportion of LD blocks with posterior probability of window variance (PPWV), of ≥ 95% at 0.001% variance threshold that did not contain a causal variant. (d) Average prediction accuracy in an independent sample, defined as the squared correlation of the predicted and simulated genetic value, with error bars giving the SD.

Second, we then compared the area under the precision-recall curve (AUPRC) for MLMA methods, BayesR and BayesRR-RC. For BayesR and BayesRR-RC we propose a posterior probability window variance (PPWV) approach [22], which provides a probabilistic determination of association of a given LD block, genomic window, gene, or upstream region, relative to the amount of phenotypic variation attributable to that window. Our PPWV approach determines the posterior inclusion probability that each region and each gene contributes at least 0.001% to the 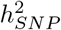, with theory outlined in the Methods suggesting that the FDR is well controlled. Across the 20 simulated generative models, we calculated precision-recall curves, where associations are defined as LD blocks (markers in LD *R*^2^ ≥ 0.1 within 1MB) with PPWV of ≥ 95% at 0.001% proportion of variance explained. True positives were the number of identified LD blocks that contained a causal variant. False positives were the number of identified LD blocks that did not contain a causal variant. For frequentist MLMA methods, which estimate SNP effects and test for association one marker at a time, results are typically reduced using LD patterns to a subset of the strongest associated LD-independent variables (a clumping approach). Thus, we first selected the strongest associated markers through LD clumping (LD *R*^2^ ≥ 0.01 within 1MB). True associations were then defined as clumped LD-independent selected SNPs that were in LD (LD *R*^2^ ≥ 0.01) with a simulated causal variant. We find that BayesR and BayesRR-RC have roughly equivalent precision-recall performance, and we find that our PPWV approach outperforms MLMA methods in their precision-recall curves across all 20 generative genetic models (Figure 2b). This demonstrates that our proposed PPWV approach can improve the identification of associated genomic regions as compared to MLMA approaches.

Third, we then wished to compare the FDR control across all generative models at the thresholds used for discovery (Figure 2). For MLMA approaches, we set the threshold at 5×10^−8^ and defined false associations as selected SNPs that were not in high LD (LD *R*^2^ ≥ 0.01) with simulated causal variants. For BayesR and BayesRR-RC, the false discovery rate was defined as the proportion of LD blocks (LD *R*^2^ ≥ 0.1 within 1MB) with PPWV of ≥ 95% at 0.001% proportion of variance explained that did not contain a causal variant. This is not a strict definition of FDR, but instead captures the ability of MLMA and Bayesian approaches to localise an effect to within *R*^2^ ≥ 0.01, and *R*^2^ ≥ 0.1 respectively, providing a direct comparison of regions identified with our PPWV approach and those identified using p-value based clumping to give LD-independent associations. BayesR, BayesRR-RC, and MLMAi provided control of this FDR metric across the range of generative models (Figure 2c). MLMA approaches provided poor control of the FDR when causal variants were high-LD (Figure 2c), suggesting that not controlling for local LD leads to regions on the same chromosome as the causal variant, but with very low LD (LD *R*^2^ ≤ 0.01) with underlying causal variants, entering the model. The BayesRR-RC prior allows more markers to enter the model due to the greater number of smaller variance components and thus while the FDR is always ~ 5% or less on average, it is slightly elevated as compared to the sparser BayesR model (Figure 2c). We also explore this approach in additional simulations for regions of the DNA (Figure S4), confirming also that population stratification and relatedness are well-controlled for in the BayesRR-RC model as compared to a MLMA models with the leading PCs of the genomic data included (Figure S5). Thus, our results suggest that BayesRR-RC is appropriate for GWAS, providing accurate estimation and discovery of associated genomic regions, across a full range of underlying generative genetic models.

Additionally, we also focused on the ability of our approach to identify candidate SNPs and to provide a probabilistic assessment of the most likely associated set of SNP markers (to fine-map associated regions). We show that our PPWV approach is analogous to the approach suggested in a recent paper (SuSiE [23]) of selecting credible sets of markers with high probability of association (Figure S4), finding that BayesRR-RC has higher power to localise associations to sets of SNP markers. The advantage of BayesRR-RC is also that assessment of associated regions is done genome-wide, with estimates obtained through simple summary of the posterior distribution rather than running numerous statistical models at different genomic regions. We also show that taking annotation information into account in BayesRR-RC can assist in marker selection among markers in LD (Figure S3).

### Genomic prediction

Finally, we used our simulation study design to explore the performance of BayesR and BayesRR-RC for genomic prediction. We randomly selected 10,000 individuals from the UK Biobank that were unrelated to those used in the simulation, we calculated the predicted genetic value and determined the correlation with the simulated genetic value. We compared four different genomic prediction models proposed in a recent paper (MegaPRS [24]) as it is suggested that these outperform a BayesR model, and all other approaches. We find that BayesR and BayesRR-RC outperforms all MegaPRS methods across all generative models, with BayesRR-RC very marginally outperforming BayesR in the enrichment simulations of each of the 20 generative genetic models (Figure 2d). In these simulation settings, recently proposed summary statistic prediction approaches [24] rarely outperform the best selected MLMAi genetic predictor at the best p-value clumping and thresholding settings (Figure 2d). Thus, we find that genomic predictors created from BayesR and BayesRR-RC models show improved prediction performance across a full range of underlying generative genetic models.

### A hybrid parallel Gibbs sampling scheme for large-scale genomic data

One of the major limitations preventing the application of Bayesian approaches to large-scale genomic data is the view that the computation of a posterior distribution is too expensive. Thus, having demonstrated the effectiveness of BayesRR-RC we then overcome a major-hurdle limiting the application of penalized regression approaches to large-scale biobank data, by deriving a Bulk Synchronous hybrid-parallel (BSP) sampling scheme for Eq.(1) that allows both the data and the compute tasks to be split within and across compute nodes in a series of message-passing interface (MPI) tasks. This BSP Gibbs sampling scheme, implemented based on a hybrid MPI + OpenMP model with residual updating and message interfacing, allows the MCMC Gibbs sampling simulations to retain accuracy of the estimation of the partial regression coefficients of each SNP marker ***β***_*φ*_ (the joint effect of each marker, conditional on all other markers), whilst allowing the marker effects to be updated in parallel (see Methods and simulation study of Figure S6).

Our Gibbs sampling algorithm enables all sampling steps to utilize genetic data stored in mixed binary/sparse-index representation, reducing computational complexity of a single Gibbs step from *𝒪*(*n*) to *𝒪*(*n*_*z*_), with *n*_*z*_ the number of non-zero genotypes. This provides a highly vectorizable mixed representation of genomic marker data as a series of indices (Figure S7) and this facilitates highly vectorized and highly parallel SNP-phenotype covariance estimation (dot product calculation) in a series of look-up tables which greatly extends previous sparse residual updating schemes. We provide publicly available open source software (see Code Availability) with capacity to easily extend to a wider range of models than that demonstrated here (see Methods). Our software requires as little as 22 seconds per MCMC sample to estimate 78 group-specific 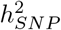 parameters, and the inclusion probabilities and effect sizes of 8,433,421 markers in 382,466 individuals on standard Intel Xeon CPU processors (Figure S7, see Code Availability for hardware specifications).

### The genetic architecture of four complex traits in the UK Biobank

This sampling scheme enabled us to apply the model to cardiovascular disease outcomes (CAD), type-2 diabetes (T2D), body mass index (BMI) and height measured for 382,466 unrelated individuals from the UK Biobank data genotyped at 8,433,421 imputed SNP markers. These markers were selected as they overlap with the Estonian Genome Centre data (see Methods) and have minor allele frequency >0.0002. Although the model can account for relatedness and data structure automatically [14, 25] (Figure S5), we wished to contrast the genetic architecture of different phenotypes and estimate the phenotypic variance contributed by MAF-LD-annotation groups from markers that enter the model only due to LD with underlying causal variants (as closely as we can in a correlational study). Thus, we also adjust each phenotype for age, sex, year of birth, genotype batch effects, UK Biobank assessment centre, and the leading 20 principal components of the SNP data. We conducted a series of convergence diagnostic analyses of the posterior distributions to ensure we obtained estimates from a converged set of four Gibbs chains, each run for 6,000 iterations with a thin of 5 for each trait (Figure S8 to S11).

We find that 32-44% of the 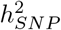 is attributable to intronic regions, 12-25% is attributable to exonic regions, 22-28% is attributable to markers 10-500kb upstream of genes, with proximal (within 10kb) promotors, enhancers and transcription factor binding sites cumulatively contributing <10% (Figure 3b, Figure S12, with estimates summed across MAF and LD groups Table 1, and full results in Table S1). The large contribution of exonic and intronic annotations to variation is in-line with the fact that these annotations account for ~ 40% of the total genome length. All four traits show the same pattern of group-specific variation, with the exception of height, where the proportion of 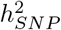 attributable to exons is almost twice as large as the other phenotypes (Figure 3b, Figure S12, Table 1, and Table S1). For all annotation groups in exons, introns, and within 500kb of genes across all traits, ≥ 60% of the 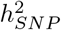 attributable to these groups is contributed by many thousands of common variants, each of small effect (Figure 3b, Figures S12 and S13).

**Figure 3.**
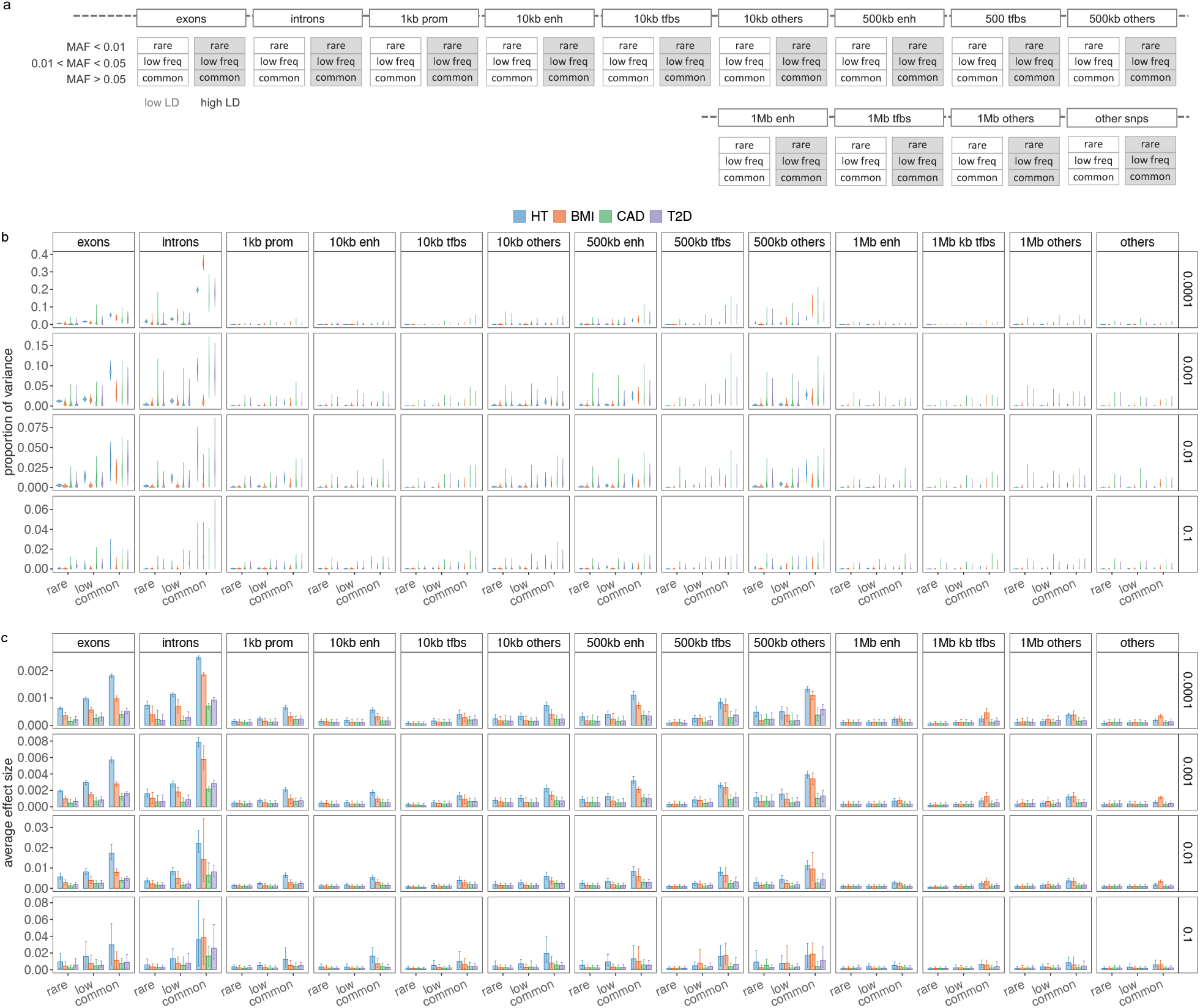
Genetic architecture of enrichment for height (HT), body mass index (BMI), cardiovascular disease (CAD) and type-2 diabetes (T2D) for 382,466 unrelated European ancestry UK Biobank individuals genotyped at 8,430,446 SNP markers. (a) We partition SNP markers into 7 location annotations (coding regions, intronic regions, and windows 1kb, 1-10kb, 10-500kb and 500kb-1Mb upstream of genes, with other SNPs grouped in a category labelled “others”). Windows 1-10kb, 10-500kb and 500kb-1Mb upstream of genes are further split into SNPs mapped to enhancers (enh), transcription factor binding sites (tfbs) and others. Within each of the 13 annotations, we have three minor allele frequency groups (MAF≤0.01, 0.01<MAF≤0.05, and MAF>0.05), and then each MAF group is further split into 2 based on median LD score. This gives 78 groups for which our BayesRR-RC model jointly estimates the phenotypic variation attributable to, and the SNP marker effects within, each group. For each of the 78 groups, SNPs were modelled using five mixture groups with variance equal to the phenotypic variance attributable to the group multiplied by constants (mixture 0 = 0, mixture 1 = 0.0001, 2 = 0.001, 3 = 0.01, 4 = 0.1). (b) Posterior distribution of the proportion of the total phenotypic variance attributable to the SNP markers that is contributed by each of the four non-zero mixtures within each MAF-annotation group for HT, BMI, CAD and T2D. Within these, are boxplots of the posterior mean and 95% credible intervals. Values are summed over LD groups. (c) Bar plots with error bars giving the 95% credible intervals for the average effect size of markers in the model for each MAF-annotation group, split by mixture.

Our estimates compare similarly to those obtained by RHEmc and SumHer, but differ to those obtained by LDSC (Table 2 and Tables S8, S9 and S10 for full results). In addition to providing variance component estimates, our model facilitates assessment of differences in the underlying effect size distribution across annotation groups. For each group, we modelled the SNP effects as coming from a series of five Gaussian mixtures, and we find that at least 45% of the 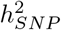 attributable to both introns and 500kb upstream regions is underlain by many thousands of SNPs that on average each contribute 0.001% (estimates summed across MAF and LD groups in Figure 3b, Figures S12 and S13). In contrast, the variance is spread more evenly across the mixtures for the other groups, implying that 10-500kb upstream regions and introns are more polygenic than other groups. This is especially so for BMI where 35% of the 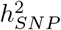 is attributable to many thousands of intronic variants (Figure 3 and Figure S13). Therefore, we find that the polygenicity of the genetic effects varies across different genomic regions, with remarkably consistent patterns across traits in the partitioning of 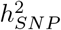 across the genome.

**Table 2.** Proportion of genetic variance genome-wide and predominantly explained by common SNPs located 10-500kb upstream of genes and coding regions for height (HT), body mass index (BMI), type-2 diabetes (T2D) and cardiovascular disease (CAD). ∗ RHEmc [21], sLDSC [11] and SumHer [6] provide the total SNP heritability observed (%) and single heritability estimates per genetic component (see Supplementary Tables 8,9,10) that we summarised to obtain the proportion of genetic variance attributed to exonic regions, intronic regions and windows 1kb, 1-10kb and 10-500kb upstream of genes.

| group | trait | BayesRR-RC<br>posterior mean (95% CI) | RHE-mc*<br>$h_{obs}^2$ (se) % | sLDSC*<br>$h_{obs}^2$ (se) % | SumHer*<br>$h_{obs}^2$ (se) % |
| --- | --- | --- | --- | --- | --- |
| *variance attributable to SNP markers genome-wide* | HT | 57.66 (56.09,59.14) | 63.28 (3.57) | 64.16 (2.86) | 98.58 (0.69) |
|  | BMI | 28.74 (27.62,30.0) | 26.76 (1.06) | 31.03 (0.9) | 44.98 (0.53) |
|  | CAD | 5.94 (5.30,6.67) | 4.49 (>100) | 4.73 (0.28) | 7.33 (0.43) |
|  | T2D | 8.45 (7.83,9.18) | 6.90 (0.47) | 6.53 (0.3) | 11.65 (0.44) |
| proportion of genetic variance attributable to exonic regions of genes | HT | 24.75 (23.39,26.071) | 27.09 | 3.00 | 16.74 |
|  | BMI | 12.98 (10.98,14.84) | 12.62 | 4.37 | 7.60 |
|  | CAD | 13.23 (8.40,18.84) | 18.68 | 1.69 | 15.34 |
|  | T2D | 14.49 (10.74,18.54) | 14.60 | 2.46 | 10.12 |
| proportion of genetic variance attributable to intronic regions of genes | HT | 41.54 (39.91,43.39) | 41.60 | 46.07 | 43.03 |
|  | BMI | 44.17 (41.36,47.25) | 47.87 | 44.61 | 48.19 |
|  | CAD | 32.05 (24.98,39.51) | 41.15 | 47.22 | 41.94 |
|  | T2D | 37.28 (32.22,42.57) | 48.66 | 38.52 | 48.02 |
| proportion of genetic variance attributable to snps 1kb upstream of genes | HT | 2.81 (2.24,3.42) | 1.76 | 1.46 | 1.74 |
|  | BMI | 1.62 (0.75,2.69) | 0.36 | 1.90 | 1.15 |
|  | CAD | 4.20 (1.71,7.55) | 2.49 | <0.00 | 1.26 |
|  | T2D | 3.58 (1.77,5.86) | 3.40 | <0.00 | 1.57 |
| proportion of genetic variance attributable to snps 10kb upstream of genes | HT | 6.60 (5.84,7.40) | 6.73 | 4.29 | 12.87 |
|  | BMI | 5.28 (3.92,6.87) | 3.19 | 6.58 | 4.10 |
|  | CAD | 13.06 (8.70,18.16) | 5.70 | 6.02 | 8.91 |
|  | T2D | 9.08 (5.90,13.28) | 4.02 | 20.44 | 7.56 |
| proportion of genetic variance attributable to snps 500kb upstream of genes | HT | 22.13 (21.00,23.40) | 21.53 | 37.23 | 24.14 |
|  | BMI | 28.58 (26.41,31.01) | 28.81 | 35.86 | 31.17 |
|  | CAD | 28.02 (21.24,35.04) | 30.23 | 38.90 | 29.58 |
|  | T2D | 27.42 (22.68,32.36) | 24.33 | 32.49 | 27.47 |
| *proportion of genetic variance attributable to exonic regions that is explained by common variants* | HT | 72.09 (69.77,74.14) | 62.62 | 75.35 | 51.22 |
|  | BMI | 69.41 (62.60,76.42) | 59.67 | 16.43 | 54.31 |
|  | CAD | 64.97 (43.08,83.16) | 61.72 | >100 | 49.17 |
|  | T2D | 68.57 (56.00,79.82) | 66.33 | >100 | 64.11 |
| *proportion of genetic variance attributable to intronic regions that is explained by common variants* | HT | 81.19 (79.30,83.02) | 79.96 | 70.88 | 66.12 |
|  | BMI | 85.05 (78.28,91.49) | 86.10 | 70.62 | 69.68 |
|  | CAD | 84.68 (65.64,95.91) | 96.55 | 61.11 | 78.17 |
|  | T2D | 87.62 (75.65,94.85) | 87.63 | 67.93 | 71.39 |
| *proportion of genetic variance attributable to snps 500kb upstream of genes that is explained by common variants* | HT | 81.59 (78.91,83.96) | 80.66 | 71.86 | 77.28 |
|  | BMI | 86.78 (80.56,91.60) | 89.95 | 67.38 | 74.81 |
|  | CAD | 66.49 (49.11,81.79) | 88.51 | 60.52 | 79.91 |
|  | T2D | 72.35 (58.71,83.75) | 94.91 | 69.48 | 75.12 |

We then directly assessed the magnitude of the effect sizes within each group, calculating the average effect size of markers in the model, for each mixture, within each group, at each iteration of the model. Across traits, effect sizes scale to their differences in 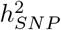, and we find that exonic and intronic region effect sizes were higher than the rest of the genome, across all mixture groups, followed by 10-500kb upstream regions (Figure 3c). We find little evidence that SNPs located in proximal promotors, enhancers, and transcription factor binding sites within 10kb of genes showed average effect sizes that were higher than SNPs located 1MB away from genes, or those that were not mapped to a specific category, with perhaps the exception of high MAF variants (Figure 3c). Generally, all phenotypes simply appear to be predominantly underlain by very many common variants, with SNPs within distal regulatory regions, coding and intronic regions each contributing more to the phenotypic variance and having higher allele substitution effects. As these results are for the effect sizes of standardized markers, it represents the square root of the average contribution of a marker to the total variance. Thus, we also re-scaled the marker effects by the standard deviation of each marker, to give effect sizes on the allele substitution effect size scale. Again, average effect sizes scaled to the 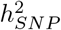 of the traits and we find that rare variants have higher average allele substitution effects than common variants for exonic, intronic, promotors and enhancers (Figure S13b). An exception to these patterns were BMI-associated intronic and 10-500kb group SNPs, where we find no evidence that the allele substitution effect size differs across frequency groups (Figure S13b). We also did not find evidence that the allele substitution effect size differed across frequency groups for transcription factor binding sites, distal SNPs 1MB upstream of genes, or those not mapping to an annotation group (Figure S13b). These results support our simulation results that both poor estimation accuracy of other approaches and an assumption that assuming an equal contribution of each marker within each annotation group may give misleading results when determining SNP enrichment. Evolutionary theory predicts that selection should result in higher effect sizes for rare variants and our results imply that selection pressures vary both across traits, but also across genomic regions with exons, promotors, and enhancers showing the strongest differentiation of effect sizes across frequency groups as compared to the rest of the genome.

### Discovery of associated genomic regions

We then partitioned the variance attributed to SNP markers across 50kb regions of the genome, then across SNPs annotated to genes, and then to LD blocks of the DNA using our PPWV approach. We find 1660 50kb regions for height with ≥ 95% posterior probability of explaining 0.001% of the 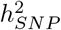, 520 regions for BMI, 70 regions for CAD and 87 regions for T2D (Figure 4a and Table 3). We then map SNPs to their closest gene (+/− 50kb from SNP position) and we use our annotations to label them (see Methods). We find 243 independent coding regions for height with ≥ 95% posterior probability of explaining at least 0.001% of the 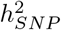, 29 independent coding regions for BMI, 5 for CAD and 13 for T2D. We find many more associations in the cis region of genes with 1254 independent cis-regions for height with ≥ 95% posterior probability of explaining 0.001% of the 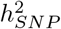, 1765 independent cis-regions for BMI, 1166 for CAD and 1221 for T2D. We additionally find 9 independent promoter regions and 1072 independent introns for height with ≥ 95% posterior probability of explaining at least 0.001% of the 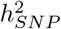, 1162 independent intronic gene regions for BMI, 307 for CAD and 347 for T2D. When we calculate the number of exons, introns, promotors and cis regions with ≥ 95% posterior probability of explaining 0.001% of the 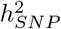, as a proportion of the total number within each chromosome, we find that up to 24% of the genes on each chromosome are associated with each of the four traits (Figure 4b). Generally, we find that only 1% or less of the available exons and promotor regions of genes per chromosome show an association with each of the phenotypes, but up to 14% of the available intronic regions and up to 10% of the cis-regions surrounding genes contribute to the phenotypic variance with ≥ 95% probability (Figure 4b). The variance contributed by each exonic, intronic, promotor, or cis region is typically only a small fraction of a percent, with largest effect sizes being the exonic region of GDF5 contributing 0.26% (95% CI 0.21, 0.32) to the phenotypic variance of height, the intronic region of FTO contributing 0.48% (95% CI 0.29, 1.12) to BMI, both the exonic- and intronic-region of LPA contributing a combined 0.08% (95% CI 0.04, 0.13) to the risk of CAD, and the intronic region of TCF7L2 contributing 0.28% (95% CI 0.23, 0.35) to the risk of T2D (Figure 4c, full results in Table S2 to S5). Taken together, these results support an infinitesimal contribution of many thousands of genes to common complex trait variation and give joint estimates of the proportions of variance contributed by each gene and their probability of association.

**Table 3.** Summary of findings for height (HT), body mass index (BMI), type-2 diabetes (T2D) and cardiovascular disease (CAD). *∗* SNPs located up to +/− 50kb from the closest gene.

| findings | method | HT | BMI | CAD | T2D |
| --- | --- | --- | --- | --- | --- |
| associated SNPs | COJO-plink2 | 1673 | 517 | 34 | 85 |
|  | COJO-BoltLMM | 2131 | 565 | 34 | 84 |
|  | COJO-Regenie | 2134 | 555 | 34 | 82 |
| 50kb regions (PPWV$\geq 95\%$) | BayesRR-RC | 1660 | 520 | 70 | 87 |
| genic regions (PPWV$\geq 95\%$) | BayesRR-RC | 2578 | 2956 | 1478 | 1581 |
| exons |  | 243 | 29 | 5 | 13 |
| introns |  | 1072 | 1162 | 307 | 347 |
| cis* |  | 1254 | 1765 | 1166 | 1221 |
| SNPs (PIP$\geq 95\%$) | BayesRR-RC | 360 | 20 | 2 | 9 |
| exons |  | 216 | 16 | 1 | 4 |
| introns |  | 73 | 2 | 1 | 5 |
| 10-500kb |  | 48 | 1 | 0 | 0 |
| LD clumps with $r^2 = 0.1$ (PPWV$\geq 95\%$) | BayesRR-RC | 1220 | 206 | 16 | 19 |

**Figure 4.**
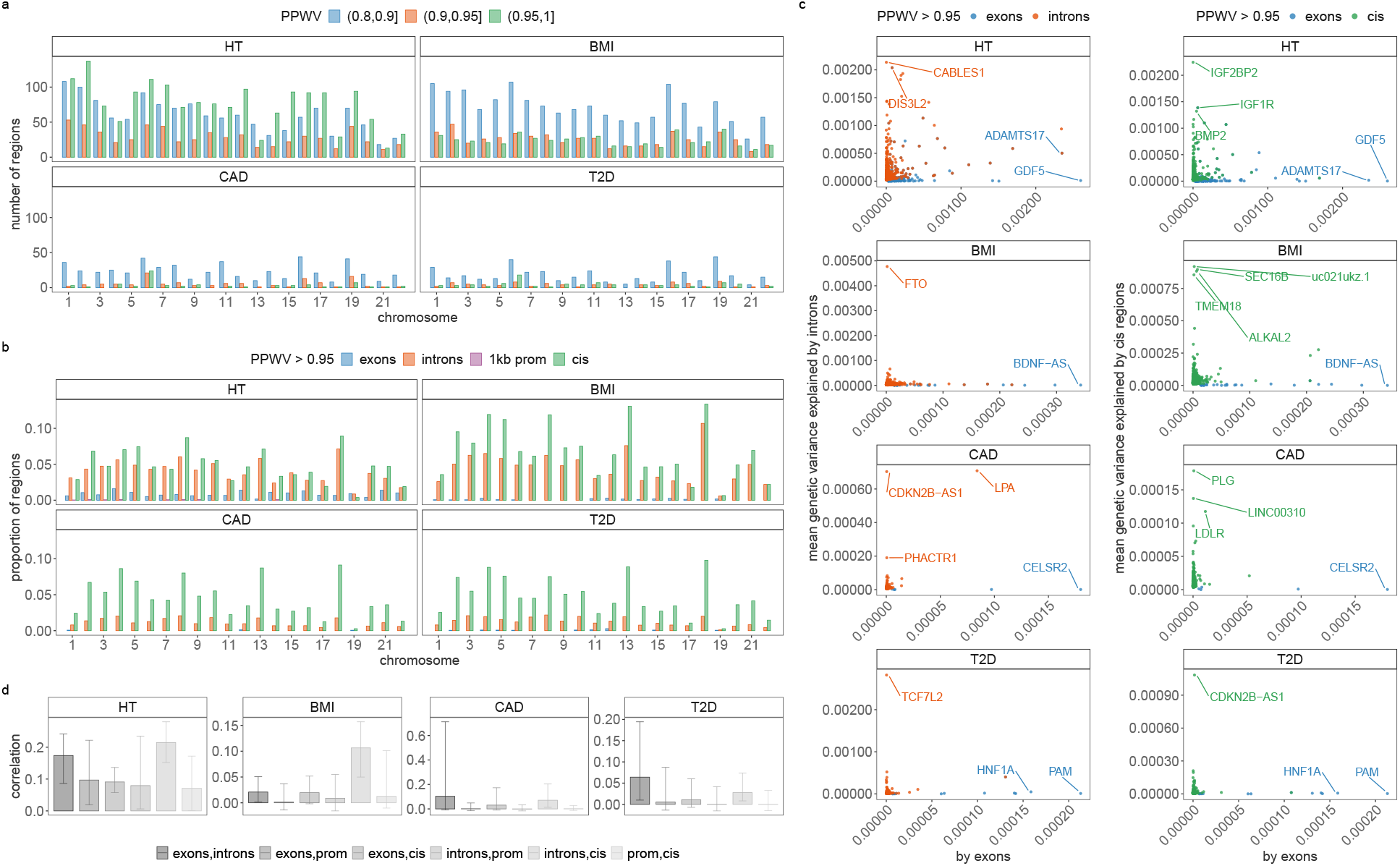
Contribution of genes and 50kb regions to height (HT), body-mass-index (BMI), cardiovas-cular disease (CAD) and type-2-diabetes (T2D). (a) We grouped SNPs in 50kb-regions genome-wide and estimated the sum of the squared regression coefficient estimates for each 50kb-region. We then select the number of 50kb regions that explain at least 0.001 % of the variance attributed to all SNP markers in 80%, 90% and 95% of the iterations. This gives a measure called the posterior probability that the window variance (PPWV) [22] exceeds 1/10,000 of the phenotypic variation attributed to SNP markers. (b) We mapped SNPs to the closest gene +/− 50kb from the SNP position and labelled them as located in a coding region, an intron, 1kb upstream of a gene using our functional annotations (Figure 3a). Remaining snps are labelled as located in a cis-region (up to +/− 50kb from a gene). We then select the number of regions where PPWV is higher than 95% and explains at least 0.001 % of the phenotypic variance attributed to all SNP markers. We then calculate the number of significant coding regions, introns, 1kb regions and cis regions as a proportion of the total number of genes for each chromosome. Genic associations that explain at least 0.001% of the phenotypic variance attributed to all SNP markers are again spread across chromosomes according to the chromosome length. (c) Shows the mean of the phenotypic variance attributed to intron and cis regions (y-axis) and coding regions (x-axis) that explain at least 0.001 % of the phenotypic variance attributable to SNP markers in ≥ 95% of the iterations (PPWV>0.95). These results provide joint estimates of the proportions of variance contributed by different gene bodies and automatic fine-mapping of gene bodies and their cis-regulatory regions. For example, introns and cis-regulatory regions of FTO respectively contribute 0.48% (95% CI 0.29, 1.12) and 0.01% (95% CI 0, 0.01) to the phenotypic variance of BMI.

For each gene, we also calculated the phenotypic variance contributed by exonic, intronic, promotor region, and cis SNPs and then calculated the correlation among the variances explained by the groups across genes. Across traits, we find small positive correlations of the variance attributable to exonic and intronic regions of 0.17 (0.09, 0.24 95% CI) for height, 0.02 (0.001, 0.05 95% CI) for BMI, 0.103 (−0.007, 0.71 95% CI) for CAD, and 0.064 (0.01, 0.19 95% CI) for T2D. Similarly, we find small positive correlations between introns and cis regions(Figure 4d). With the exception of height, there was no evidence for a relationship among the following groups: (i) SNPs in the exons of each gene and SNPs +/− 50kb outside of the exon and promotor regions; (ii) SNPs in the exons of each gene and SNPs in proximal promotors; and (iii) intronic SNPs and SNPs in promotor regions (Figure 4d). This implies that trait associated SNPs in proximal and distal regulatory regions are largely independent of the effects of SNPs in their closest exon, as they do not align in terms of the variance they explain (Figure 4d). For height, small weakly positive correlations across all gene regions in their contribution to variance, implies a degree of alignment across genes in regulatory variants and the closest exon (Figure 4d). These results suggest a regulatory link between introns and distal cis regions outside of the promotor, or that introns may be correlated with structural variation. They also imply that the variance contributed by regulatory regions and those in the closest coding regions are not strongly coupled for these common complex traits.

Finally, our approach provides automatic fine-mapping of SNP loci, and of these region- and gene-level associations, 360 SNPs for height, 20 for BMI, 2 for CAD and 9 for T2D could be mapped to a single SNP with greater than 95% inclusion probability across all 4 chains (Supplementary Table S6, Figure S14). Of these fine-mapped SNPs, only 53.45% are top loci with a p-value < 5×10^−8^ from the fastGWAS UK Biobank summary statistic data for standing height, BMI, angina / heart attack and type-2 diabetes (fastGWA, see Code Availability). This highlights that selecting on the top SNP markers identified by standard association studies would give a different set of variants than those obtained from selecting high PIP SNPs.

### Out-of-sample prediction into another European healthcare system

Finally, we then generated a full posterior predictive distribution for each trait in each of 32,500 individuals from the Estonian Genome Centre data, which allows the transmission of uncertainty in the marker effect estimates from the UK Biobank to the genomic predictors created in Estonia. First, despite this study having almost half the sample size, we show improved genomic prediction as compared to recently proposed summary statistic approaches [26], when taking the mean of the predictor across iterations and correlating this with the phenotype with correlation of 0.62 for height, 0.34 for BMI, 0.16 for T2D, and 0.07 for CAD (Figure S15a). The area under the receiver operator curve (AUC) for T2D was 0.67 and 0.57 for CAD. In comparison, using the 64 BLD-LDAK annotations recommended by a recent study [24], the highest prediction accuracy obtained from MegaPRS was 0.55 for height, 0.32 for BMI, 0.10 for T2D, and 0.05 for CAD.

We then estimated the distribution of the partial correlations between the trait and genomic predictors created from our different annotation groups and find that exonic, intronic, and 10-500kb upstream regions contribute proportionally more to the prediction accuracy than other genomic groups, replicating our results from the UK Biobank (Figure S15). We find evidence for zero/low correlations of genomic predictors created from different annotation groups, which supports our results from the UK Biobank (Figure S15e). This suggests that individuals have a different portfolio of risk variants, with different genomic regions contributing for different individuals to their overall genetic value, as expected under a highly polygenic model.

Additionally, for height and BMI we also determined the proportion of the posterior predictive distribution for each individual that was within +/−1 SD of their true phenotypic value. On average 67.5% of an individuals posterior predictive distribution is within +/−1 SD of their true phenotype for BMI and 75% for height, with similar prediction accuracy across individuals (Figure S15c). For T2D and CAD, we extended the PCF metric, typically defined as the proportion of cases with larger estimated risk the then top *p*^*th*^ percentile of the distribution of genetic risk in the general population. For each individual, we calculated the proportion of their posterior predictive distribution that falls above the top 25% of the distribution of genetic risk in the general population. The distribution of these probabilities is shown for confirmed cases and those without diagnosis in the Estonian Biobank (Figure S15d). We find 25 individuals for T2D and 15 individuals for CAD where ≥90% of their posterior predictive distribution is within the high risk group of which 40% and 18% are currently defined as cases for T2D and CAD respectively based on recent medical records. This is compared to 1% and 2% case rate for those with ≤10% probability of being in the high risk group for T2D and CAD respectively, giving an odds ratio of 20 and 18 between the ≤90% and ≥10% groups. However, our results clearly show that the individual-level sensitivity and specificity of genomic prediction for these common complex diseases is very poor, as 75% of T2D cases and 92% of CAD cases have ≤50% of their distribution within the high-risk category. These results highlight how variation contained within a posterior predictive distribution that is typically ignored in human genomic prediction can be used. We show that genomic prediction for personalized medicine with patient-specific predictions or stratification of patients is currently extremely limited.

## Discussion

There is no single statistical model appropriate for all settings and thus there will always be a situation where a model poorly fits the data. We have provided theoretical and empirical evidence that a grouped Dirac spike-and-slab model (which we term BayesRR-RC), has a prior that is flexible enough to show robust model performance across the data analysed here, improving inference over commonly applied approaches. We develop a range of computational and statistical approaches which allow this, or any similar Gibbs sampling algorithm, to scale to whole genome sequence data on many hundreds of thousands of individuals. This has enabled us to compare and contrast the inferred underlying genetic distribution for four complex phenotypes under this prior, providing novel insight into the genetic architecture of these traits. We observe that all phenotypes simply appear to be predominantly underlain by very many common variants, with SNPs within distal regulatory regions, coding and intronic regions each contributing more to the phenotypic variance and having higher allele substitution effects.

There has been debate on how to best estimate SNP heritability [1, 3, 4] and here we validate that one approach could be to split SNP markers by LD to improve genetic effect size estimates. We demonstrate through theory and simulation why penalized regression models inaccurately estimate effects under multi-collinearity and how differential shrinking of SNPs can correct this bias. Recent studies have also attempted to quantify the gene architecture of complex traits, in terms of the number and contribution to phenotypic variance of markers either in coding regions, or directly involved in the expression of genes [27, 28]. Our results suggest that the proportion of genomic variation attributable to mutations in regulatory regions and mutations in the closest genic regions are largely independent. Additionally our model tests association within groups in a probabilistic way and we find 290 independent coding, 2,888 independent intronic, and 5,406 independent cis regions with ≥ 95% probability of of contributing at least 0.001% of the SNP heritability. Understand how these coding, intronic and proximal and distal regulatory regions combine to contribute to phenotypic variance remains a substantial challenge and our results suggest a predominant role for introns and for distal, and thus likely more global enhancers, rather than locally dominant proximal expression QTL. The recent “omnigenic” model [29], suggests that trait-associated variants in regulatory regions influence a local gene which is not directly causal to the disease, and also co-regulate other disease causal genes (or “core” gene). Our findings of little correlation of exonic and proximal regulatory variance and a large number of trait-associated intronic and cis regions do not rule this out, but suggest a more complex infinitesimal picture with differences occurring among traits, potentially due to their evolutionary history.

There are important caveats and limitations to consider. In this work, we do not extend past a limited number of functional annotations and thus we do not provide a model capable of further partitioning the variation into specific regulatory functions (eQTL, mQTL, pQTL etc.) or directly modelling the relationships among components. Doing this requires the use of more information in the prior, allowing more groups, potentially allowing markers to swap groups with a prior probability of function, and allowing for correlations in marker effects across groups. While our future work is in this direction, a first requirement is an improvement in annotations as MAF-LD multicollinearity biases have to be removed from studies of eQTL, mQTL, pQTL etc. before these annotations can be reliably used, as otherwise marker function will likely be biased by the data structure (e.g. common, high LD variants may be more likely to be allocated as eQTL). LDSC functional methods take the approach that SNPs can be assigned to different categories (e.g. both coding and conserved), with the categories competing against each other to explain the signal, with the downside that enrichment is relative and that the total variance is not partitioned. Here, the total variance is partitioned but this is based on preferential allocation of SNPs to coding regions, introns, and then to their nearest upstream gene position. Coding regions, introns and 10-500kb distal regions could contribute the most variance as these SNPs are most likely to be allocated accurately, with 1kb and 1-10kb groups being more ambiguous in high gene density regions and likely mislabelled. However, if this was the case then variance would still be partitioned to these mislabelled groups and it would just be evenly split across them, with experimentally validated promotor, enhancer and tfbs regions assisting to some degree in alleviating this. This was not the case, and here we see a clear pattern of increasing variance contributed, increasing average effect size, and an increasing pattern of higher rare allele substitution effects by individual markers as distance from the nearest gene increases. 10-500kb distal regions may contribute more variance as marker density and marker coverage is higher in these regions, with missing variation within 10kb upstream as causal variants are poorly correlated with SNPs. The posterior distributions for the variance explained by 1kb, 1-10kb regions, and 10-500kb regions are negatively correlated (Figure S11, meaning that these groups are competing with each other, as if variance goes to one then it is being taken away from the other (as they are in LD), and thus there is the risk that the model cannot separate these effectively. However, this is true of any enrichment analysis conducted to date and we can only make inference in the data that we have currently available. Resolving this requires the application of this model to whole genome sequence data where the total variance can be partitioned across upstream regions without marker coverage concerns. Irrespective of exactly which upstream region variance is allocated to, our inference that genic regions are uncorrelated in their contribution to variance with the promotor and upstream regions still holds as does our probabilistic inference on the associations of each gene and their contribution to the phenotypic variation.

Other approaches may also provide continuous SNP shrinkage, regularising each SNP differently, such as a Finnish horseshoe model [30], and we are working to place a grouped version of this model within our computational framework to explore this possibility. Recent work has shown that tree sequence algorithms can also be used to massively increase the scalability of methods for genomic data, making it possible to infer trees for millions of samples [31] and to conduct regression models using tools such as TreeLD [32] or inferred ancestral recombination graphs [33]. We expect that our current algorithm combining sparse dot products and highly vectorized look-up tables to outperform these methods in terms of performance as there are costs to tree-traversal and tree-calculation. However, a tree-approach would provide benefits in terms of memory usage and future work to computationally engineer the tree-structure data may be beneficial. Finally, our focus is limited to two common complex diseases with case proportions 11.6% for CAD and 7.2% for T2D within the UK Biobank. Less prevalent complex diseases, likely require additional model extensions to the prevent effect size bias as reported elsewhere [10] and this will also be a focus of future work.

## Summary

Our results provide evidence for an infinitesimal contribution of many thousands of common genomic regions to common complex trait variation and for a predominant role of intronic, exonic, and distal regulatory regions. This highlights the immense challenge of understanding the molecular underpinning of each association and the difficulties in improving the estimation of many tens of thousands of small-effect associations that are required to improve genomic prediction. This work represents a step toward maximising the probabilistic inference that can be obtained from large-scale Biobank studies.

## Methods

### Model Specification

We begin by outlining the basic model BayesR, before then presenting our extensions. Consider *p* single nucleotide polymorphism (SNP) markers. If we gather samples for *i* = 1, …*N* subjects in an *N*× *p* matrix, **G**, in which the elements are coded as 0 for homozygous individuals at the major allele, 1 for heterozygous individuals and 2 for minor allele homozygotes. Now, we wish to model their linear association with the phenotype **y** = (*y*_*i*_) of subjects *i* = 1, …, *N* in a standard linear regression model:

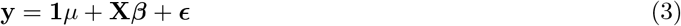

We assume that the genotypes are standardized so that 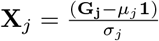 is the vector of genotypes for the *j*^*th*^ marker (*j* = 1, *p*) with zero mean and unit variance, i.e. the centered and scaled *j*^*th*^ column of **G**. The column’s mean *µ*_*j*_ *≈* 2*f*_*j*_ and the column’s standard deviation 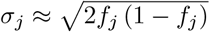 being *f*_*j*_ the minor allele frequency(MAF) of the SNP. We define ***β*** as a *p* × 1 vector of partial regression coefficients with *β*_*j*_ the effect of a 1 SD change in the *j*^*th*^ covariate, and ***E*** is a vector (*N* x 1) of residuals.

We estimate the model’s parameters using Bayesian inference, assuming that the error term 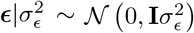. The log-likelihood of this model can be written as

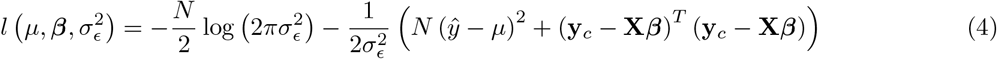

with 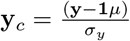 a vector of centred and scaled responses(SD 1).

As we adopt a Bayesian approach, we place priors over the model parameters. For the covariate effects, ***β***, we use a mixture prior with Dirac spike and slab components, which have been extensively used for variable selection [15, 16]. The prior induces sparsity in the model through a Dirac-delta at zero, excluding variables from the model by setting their coefficients to zero. A slab component is centered at zero and shrinks the non-zero coefficients towards zero according to the slab’s width. In our approach, the slab component is a scale mixtures of normals and thus each *β*_*j*_ ∈ ***β*** is distributed according to:

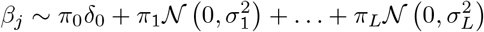

where *π*_*β*_ = (*π*_0_, *π*_1_, …, *π*_*L*_) are the mixture proportions, 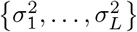 are the mixture-specific variances, and *δ*_0_ is a discrete probability mass at zero. We further constrain the prior by assuming a single parameter representing the total variance explained by the effects 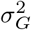, with the component-specific variances proportional to 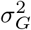 multiplied by a constant *{C*_*1*_, …, *C*_*L*_ *}* so that

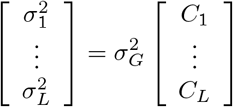

The remaining prior structure for the model is then

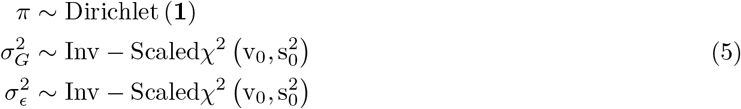

with weakly informative parameters for hyperparameters 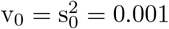.

For notational convenience, we will refer to the mixture membership labels as (*l*_0_, *l*_1_, …, *l*_*L*_) and we define a latent indicator of each SNP, *j, γ* = (*γ*_*j*_, …, *γ*_*p*_)^*T*^ with *γ*_*j,l*_ = 0 or 1, indicating whether or not the effect of SNP *j* falls into the zeroth mixture *γ*_*j,l*_ = 0, or follows a normal distribution with variance 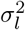. We define the “active set of coefficients” as those *β*_*j*_ such that *β*_*j*_ 0 denoted as ***β***_*γ*=0_ with cardinality ‖*γ*_*ϕ*_‖_0_. Thus the objective of our inference scheme is to compute an estimate of the posterior distribution 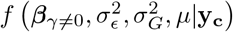 This model has been termed BayesR [12, 13] and an effective proposed Gibbs sampling scheme [13] follows the following steps:

i. Sample *µ* from 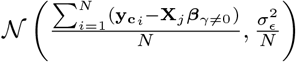
ii. sample ***β***_*γ* =0_ from its conditional as described below
iii. sample 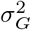 from 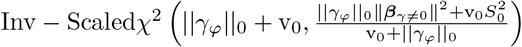
iv. sample 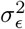 from 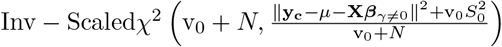

From the former algorithm, steps (i), and (iv) are straight-forward applications of conjugacy and are common to many Gibbs sampling algorithms for linear regression. Step (iii) follows from conjugacy and the assumption that the individual mixtures represent fractions of the total variance explained by the coefficients. Step (ii) is the biggest bottleneck in any linear regression problem, and in the next section we will proceed to detail the derivations of the sampling scheme for this step.

While it is not uncommon to use non-proper priors for the residual’s variance 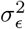, in our case we chose to keep a proper prior for algorithmic and modeling reasons as: (a) conjugacy is amenable to Gibbs sampling (b) we assume 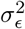 and 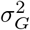 are not nuisance parameters, and in some cases we possess prior information on its distribution. It is also common to specify the distribution of *β*_*j*_ having a variance depending on the residual’s variance 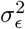, which would make the estimates transformation-invariant. Recent results suggest the estimates for 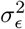 in this latter transformation-invariant formulation are biased [34]. Another concern may be that the prior’s hyperparameters induce biased estimates for small variances [35], we acknowledge that may be an issue, and allow parameters 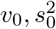 to be adjusted if deemed necessary. The scale mixture of Gaussians, allows the prior distribution to have heavier tails than a single Gaussian, which allows big effects to be shrunk to a lesser degree than small effects [17]. Finally, the original formulation of [12, 13] assumes 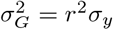 which for centered and scaled phenotypes and genotypes, with heritability *h*^2^ equal to reliability 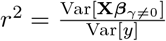 would mean 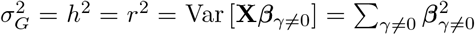, but there is no constraint in the model ensuring 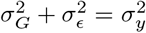. As we will see, further assumptions are necessary for having unbiased estimates of 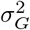 and *h*^*2*^ under varying LD and MAF. These estimates will achieve the equivalence 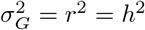 without relying in either using a point estimate of *r*^2^ [12], informative priors on 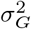, or normalising the posterior variances by 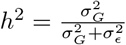 [14].

### Sampling the effects

For sampling ***β***, the challenge is two-fold: (a) determining if the effect *β*_*j*_ is part of ***β***_*γ* =0_, and if so, to which component it belongs; and then (b) sampling the vector ***β***_*γ* ≠ 0_ from a multivariate2 Gaussian with covariance matrix 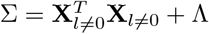 where Λ is the diagonal matrix with entries 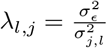, with 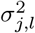the variance of the mixture component to which marker *β*_*j*_ was assigned. For (a), marginalization of each effect individually is required to compute the membership probability, which requires solving a determinant of the size of ‖*γ*_*ϕ*_‖_0_ −1 [16]. For (b), either a system of size ‖*γ*_*ϕ*_‖_0_ must be solved through LU decomposition, or Cholesky decomposition of size ‖*γ*_*ϕ*_‖_0_, and both operations are resource intensive when the size of ‖ *γ*_*ϕ*_‖_0_ is large. Instead, we determine the inclusion of a marker in the active set, along with its mixture membership and its partial regression coefficient *β*_*j*_, in single-site updates. Single-site Gibbs sampling is also known as stochastic relaxation [36] has a long history given its equivalence to iterative Gauss Siedel methods to solve matrix equations [37]. Although we choose to use the BayesR model, many alternative models can easily be placed within the iterative solving and computational framework we outline here.

In this scheme, we sample each element, *j*, of ***β*** from the full conditional posterior *f* (*β*_*j*_|***β***_*\j*_, **y**) ∝ (*α* function of *β*_*j*_, ***β***_*\j*_, **y** which can be written as *f β*_*j*_, ***β***_*\j*_, **y** = *f* (**y**|***β***) *f* (*β*_*j*_) *f* ***β***_*\j*_ where *f* (**y**|***β***) is the density the conditional distribution of **y *β*** and *f* (*β*_*j*_) and *f* ***β***_*j*_ are the densities of the prior distributions of *β*_*j*_ and ***β***_*j*_ respectively, with notation *j* representing all other covariates except *j*. The kernel of the full conditional posterior for *β*_*j*_ is proportional to the product of the likelihood, the prior distribution for *β*_*j*_ and the prior distributions of the variances, and thus ignoring factors that are constant with respect to *β*_*j*_ gives

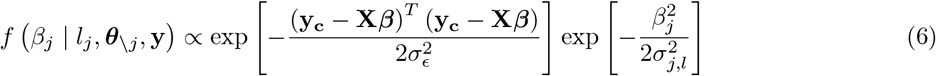

where *l*_*j*_ represents the mixture *β*_*j*_ is assigned, 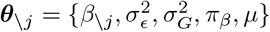 and 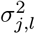 the corresponding mixture variance. We can reduce the expanded form and drop terms that are free from *β*_*j*_ as

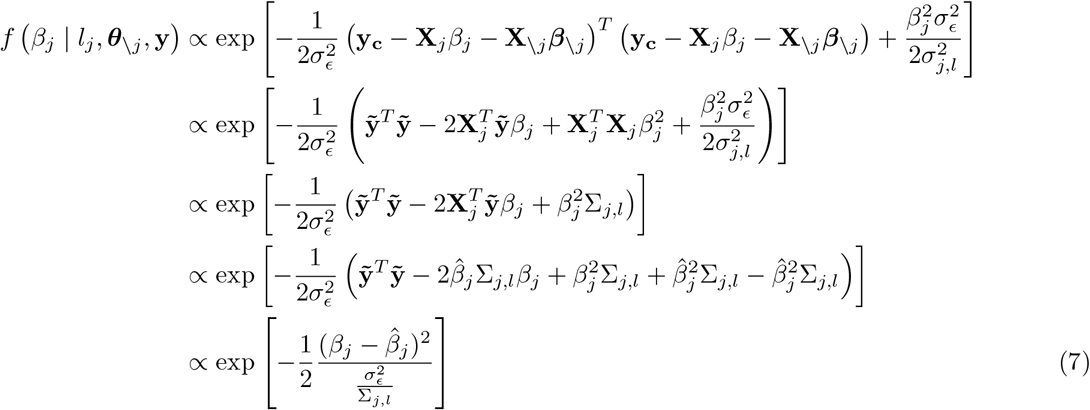

with 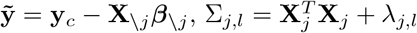 and 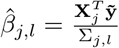. This gives the Gibbs sampling update for *β*_*j*_

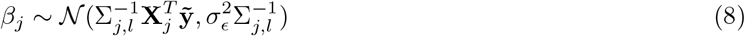

To avoid reducibility of the Markov chain, prior to drawing the effect *β*_*j*_, we first need to select the mixture *K* for each covariate *j*, and as above we can condition on the individual coordinates and to obtain the probability that a coefficient *j* belongs to a given mixture.

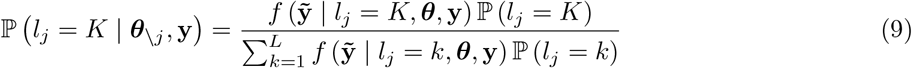

We integrate out the *β*_*j*_ coordinate following the equations above with

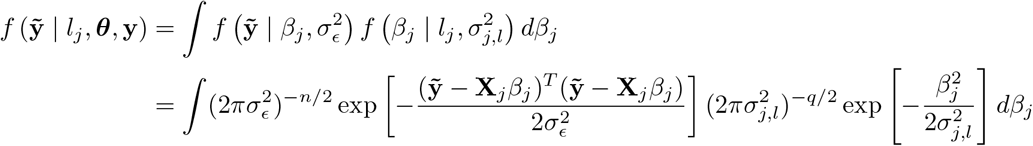

where *q* = 2. We then expand this equation using the relationship 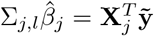 from Eq. 8 and complete the squares

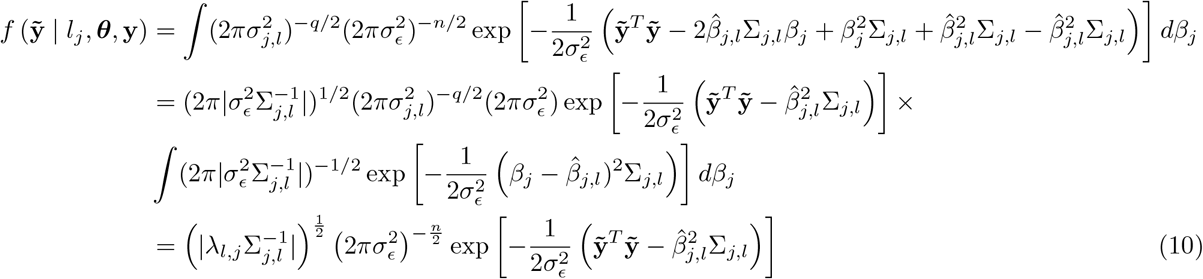

where the final reduction in Eq. 10 occurs as the integral component is now a normal distribution that integrates to 1 and then terms are removed that do not contain, nor depend upon Σ_*j,l*_ nor 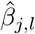. The probability for inclusion in the model in the first mixture, as compared to the spike, then depends upon the ratio

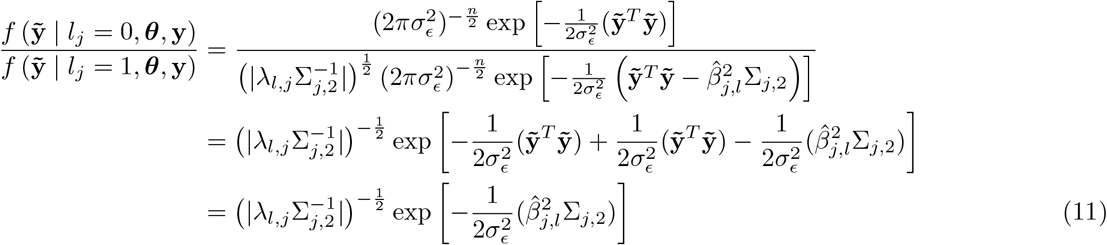

Analogous to equation 11, any comparison between mixtures has the same form and allows us to omit the 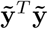 term. Thus placing Eq.11 into Eq.9 and re-arranging to a numerically more stable version [12] gives

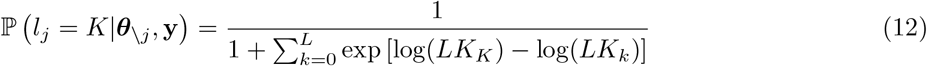

with log(*LK*_0_) = log(*π*_0_) and 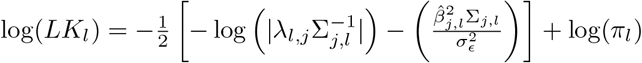 for *l* in (1 …*L*).

Having derived the regression coefficients and their inclusion probabilities, fully specifying the BayesR model, we now proceed in the following sections to: (1) derive the properties of the model parameters when applied to highly correlated genomic data (under multicollinearity) and compare these to estimates made by other approaches in the field; (2) extend the model to account for genomic annotations, minor allele frequency (MAF) and linkage disequilibrium (LD) among markers; and finally (3) derive a computational implementation that facilitate the application of the model to biobank sized data.

### Comparison to other approaches under collinearity

Genome-wide association studies have predominantly been conducted using single marker regression via ordinary least squares (OLS). Recently, it has been proposed that if aggregation due to familial or molecular similarity (e.g. population stratification) exists in the data, a better estimation approach is generalized least squares (GLS), as it poses a more general covariance structure than OLS. GLS estimates can be obtained within mixed-linear association models, which first declare all marker effects as random variables, for example, assuming that 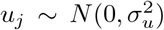, or from a mixture of distributions, with all markers in the set taken as independently and identically distributed random variables. Second, when the markers are evaluated for association, they are then treated as a fixed effect. The resulting model can be written as

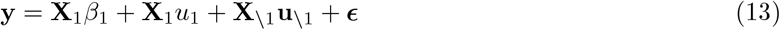

where a focal genetic marker, here **X**_1_ is fitted twice, first as a fixed effect to estimate the regression coefficient *β*_1_, and also as part of all of the other markers with their effects, *u*, estimated as random (note here \1 indicates all markers other than marker 1). Under this model the phenotypic covariance structure is

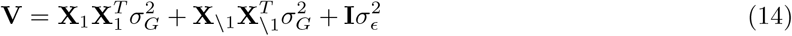

With orthogonal covariates, the estimated variance components that compose **V** can remain constant when testing each marker in turn. However, with collinearity among markers the situation becomes more complex. Below, we first describe the impact of multicollinearity on ridge regression estimates. We then outline the equivalence of a ridge regression and a mixed linear model, before then demonstrating increased variance of the estimates obtained from Eq. (13) under multicollinearity. Finally, we then go on to show that estimates from BayesR are less subject to inflated variance, except under extensive multicollinearity, before then describing how extending the model to provide minor allele frequency and LD specific hyperparameters provides estimates with improved properties across a range of underlying generative data models.

In Eq. (13) if markers were all simply estimated as random, following a single distribution, then a ridge regression estimator of Hoerl and Kennard 1970 [38] would be obtained, which was proposed to replace **X**^*T*^ **X** in the OLS solutions by **X**^*T*^ **X** + *λ***I**, with *λ* ∈ [0, *∞*] a tuning or penalty parameter. This gives the ridge regression estimator

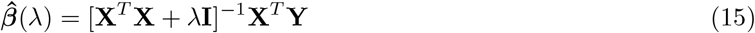

where *λ* is strictly positive and the solution or regularization path of the ridge estimate 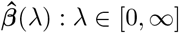 is the set of ridge estimates across the values of *λ*. The expectation of the ridge estimator

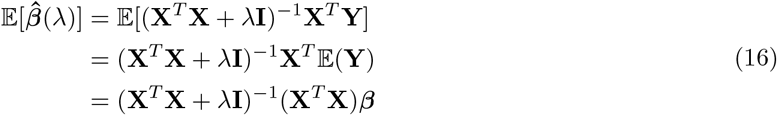

with 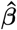 the maximum likelihood OLS estimator. If we consider an orthonormal design matrix **X**, with **X**^*T*^ **X** = **I** = (**X**^*T*^ **X**)^−1^ then we can express the relationship between 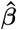, and the ridge estimator, 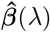, as

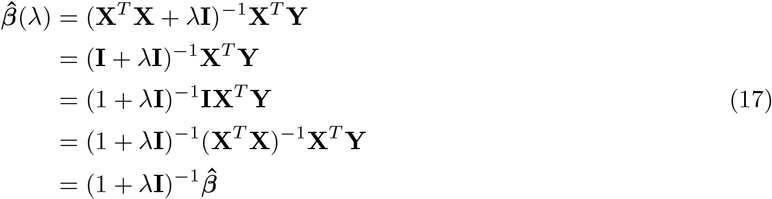

If we define **W**_*λ*_ = (**X**^*T*^ **X** + *λ***I**)^−1^(**X**^*T*^ **X**) then the ridge estimator 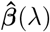 can be expressed as 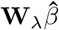 for

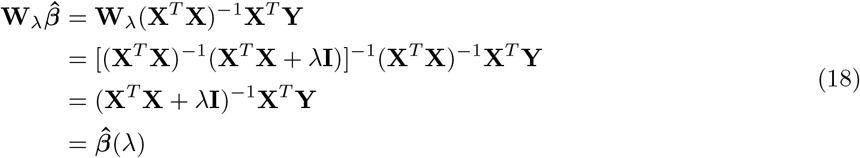

The variance of the ridge estimator is then

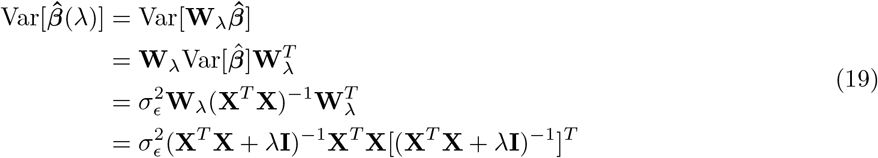

and the mean square error of 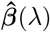 is

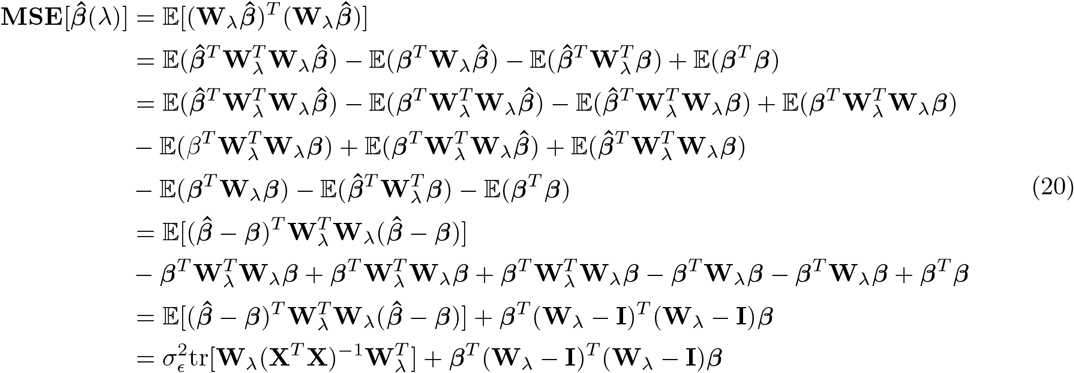

The first summand is the sum of the variances of the ridge estimator, while the second summand is the squared bias of the ridge estimator. With an orthonormal design matrix, **X**, Theorem 2 of Theobald 1974 [39] shows:

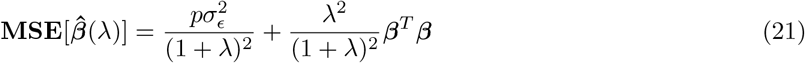

which achieves a minimum at 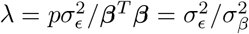, with 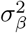 the variance of the *β* coefficients. This has been stated in the genetics literature as the optimal shrinkage parameter [40] for a ridge regression. However, this is derived under the assumption of uncorrelated covariates within the design matrix **X**.

To explore the effects of correlated covariates we use the ridge loss function, defined as

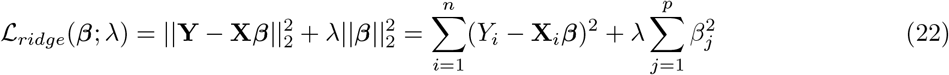

which is the sums-of-squares with a penalty, 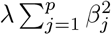, referred to as the ridge penalty, which shrinks the regression coefficients towards zero. The radius of the ridge constraint, the squared Euclidean norm of *β*, 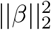, depends upon *λ*, **X** and **Y**, and taking its expectation

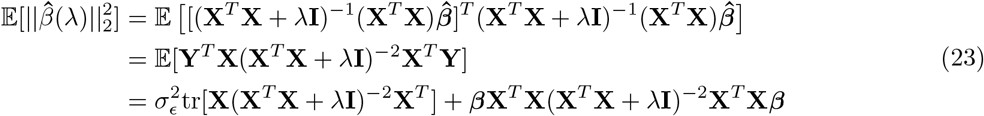

provides a measure that can be evaluated given different properties of the design matrix **X**. With the same *λ* and the same ***β***, Eq. (23) shows that the degree of collinearity among the covariates alters the variance of the estimated effects. Thus, in a ridge regression penalization does not remove collinearity but simply reduces its effects on the variance of the ridge estimator provided that the *λ* value is sufficiently large (and thus the 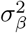 is small). We explore Eq. (23) in a simulation study described below and presented in Figure S1, Figure 1 and Figure 2. This theory is an extension of previous work [41] which showed that the inflation of the SNP heritability is proportional to a ratio of the average LD among causal variants and the markers and the average LD among all the markers, with inflation expected when causal variants are in higher LD with the markers than on average. Eq. (23) is a function of X’X, with the LD values the off-diagonal elements in X’X, but it suggests that inflation would be irrespective of the average LD across the genome, simply being expected if high-LD markers had strong effects and showing that inflation would occur only for the estimates of markers that are in LD with those causal variants. Thus, if SNP heritability is allocated across SNPs at random then estimation will on average be correct, irrespective of the LD among SNPs. If the effects of SNPs vary according to the MAF or LD of the SNP, and assumptions are made that all SNP effects are sampled from the same distribution, then this will lead to bias as the estimates at high-LD markers in strong LD with underlying causal variants will be inflated and this inflation will be sufficiently large and occur at a sufficient number of genomic locations so as to impact upon the global estimate of SNP heritability.

This issue has been detected, and demonstrated in simulation, in a number of recent papers [1–4]. However, to date it has remained little understood from a theoretical perspective. The LD-MAF corrections proposed in the literature all serve to alter the lambda value for SNPs, or sets of SNPs, so that it becomes proportional to the LD and MAF of the marker, in essence reducing the 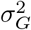, or making it more specific to the markers in question, and increasing the *λ* value for common, highly correlated covariates. The equivalence of ridge regression and mixed-linear models has been shown many times, using well-established results from prediction of random variables dating back to Henderson [42]. The model **Y** = **g** + ***E***, with **g** the genetic value of the individuals, and the model **Y** = **X*β*** + ***E***, with 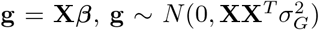 with marker effects thus ***β*** = **X**^*T*^ (**XX**^*T*^)^−1^**g**, are equivalent. Following Henderson [42], assuming 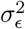 and 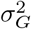 are known, with no fixed effect component, the log-likelihood can be shown to be proportional to:

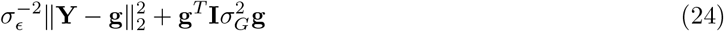

equating the partial derivatives of this mixed model loss function with respect to **g** to zero, yields the estimating equations known as Henderson’s mixed model equations. Returning to the mixed linear association model described in Eq.(13), using **u** to denote the marker effects estimated as random, *β* for the focal marker effect estimated as fixed, and assuming independent marker effects, Henderson’s mixed model equations (MME) take the form:

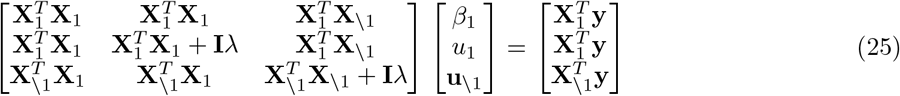

where 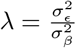. Subtracting the *u*_*1*_ from the *β* equations gives *u*_1_ = 0 and thus the MME reduce to:

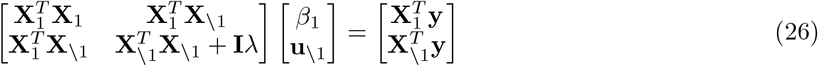

This has been derived previously [43], however there is an explicit assumption that the any estimation error of the random marker effect estimates go into the residual and does not influence the fixed estimate of the marker. For the random effect component, the equivalence with the ridge regression estimator of Eq.(15) is evident, as is the equivalence of Eq. (24) with Eq. (22) above. Thus an MLMAi model returns “ridge regression” estimate of the marker effects, and as we show above ridge regression estimates are inflated when effect sizes are higher for high LD markers. It then follows that mixed model effect size estimates could be biased when effect sizes are higher for high LD markers.

Seen in this light, we can now explore the influence of multicollinearity on the BayesR dirac spike and slab model described above and compare it to that of a ridge regression. If we denote a measure of fit, such as the ridge loss function described above, being composed of *l*(*β*) and a penalty function *pen*_*λ*_(*β*), then from a Bayesian perspective these correspond to the negative logarithms of the likelihood and the prior distribution, respectively. We can parameterize the BayesR dirac spike and slab model described above using the latent indicator of each SNP, *j, γ* = (*γ*_*j*_, …, *γ*_*p*_)^*T*^ with *γ*_*j,l*_ = 0 or 1, indicating whether or not the effect of SNP *j* follows a normal distribution with variance 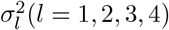. Then *p*(*γ*_*j,l*_ = 1 *π*_*l*_) = *π*_*l*_ and the prior distribution of each SNP effect *β*_*j*_ conditional on the indicator *γ*_*j,l*_ is

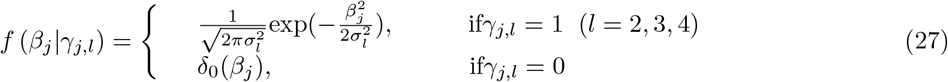

The joint distribution *p*(*β*_*j*_, *γ*_*j*_) conditional on *π*_*β*_ is

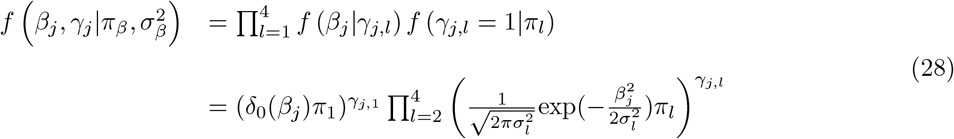

to simplify the following, we assume only a single normal distribution with *π*_1_ + *π*_2_ = 1 and we redefine the regression coefficient as *β*_*j*_ = *γ*_*j*_*α*_*j*_ with 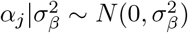. then:

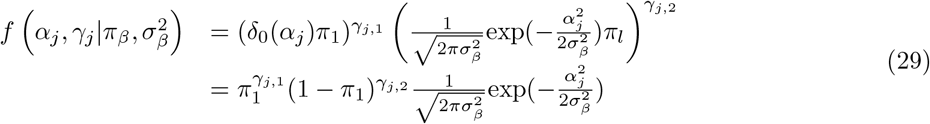

Now as above, if we define an active set of markers, **X**_*γ* ≠ 0_, as those columns of **X** where *β*_*γ* ≠ 0_, with an active set of *γ*, and 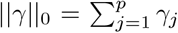 be its cardinality. The joint prior on the vector *γ, α* then factorizes across all the markers as

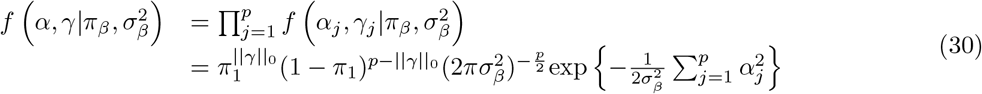

as above we can express the likelihood in terms of *γ, α* as

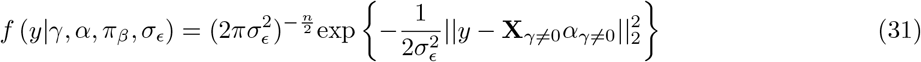

and then under this reparamterisation the posterior is given as

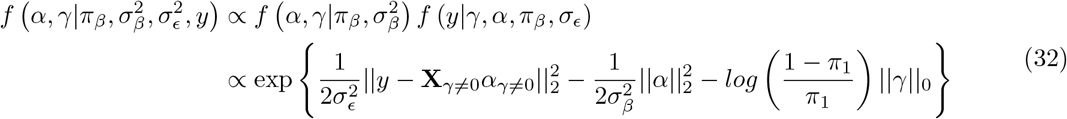

The regularized maximum a posterior estimator is equivalent to minimising over *γ, α* the least squares objective function as

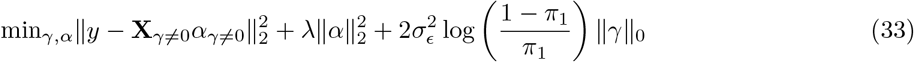

In comparison to the ridge loss function described above, the first two terms are very similar and the third term imposes a sparsity constraint on the model. The term 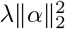 has the same expectation as in Eq. (23) but with **X** replaced with **X**_*γ*=0_. To give some insight into the influence of collinearity on E [*γ*]_0_ and on the active set, we explore a two SNP scenario.

In a single site updating scheme, the probability that the first marker enters the model is given by Eq. 12. We seek to derive the probability that the second marker enters the model conditional on the first marker being in the model. We consider a scenario where we observe our standardised outcome 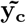 and two correlated predictors **X**_1_ and **X**_2_. We assume that 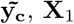 and **X**_2_ are scaled with zero mean and unit variance. We can then derive the partial least squares regression for 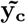 regressed on **X**_**2**_, adjusting for **X**_**1**_. If 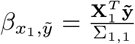, with 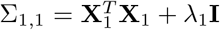, then a residual vector 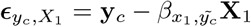 is the vector left after backfitting 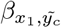 and we define 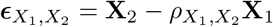 as the additional information in *X*_2_ left to fit 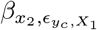, with 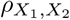 the correlation of *X*_1_ and *X*_2_. The correlation between the two residuals 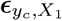 and 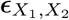 can be used to estimate 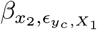, since 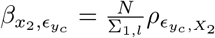. The correlation is a ratio between a covariance and a variance as

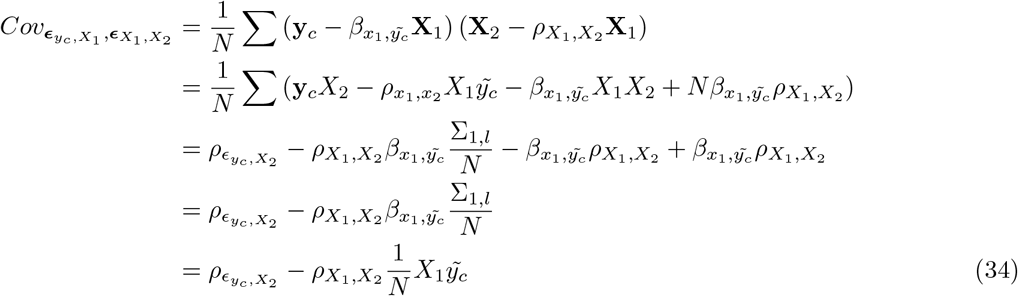

The variance in the correlation denominator is 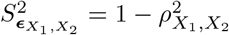 which gives

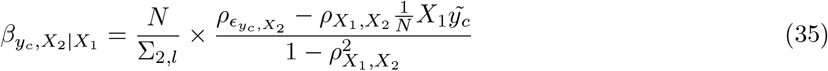

Eq. 35 can then be used in Eq. 11 and Eq. 12 to determine the posterior inclusion probability of the second covariate conditional on the first covariate being in the model. From this, the expectation, 𝔼 [‖ *γ* ‖_0_] for a two SNP scenario is then

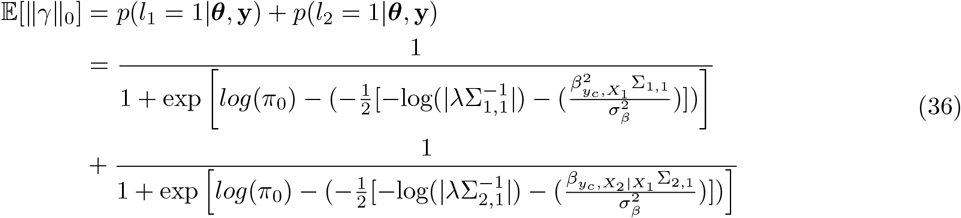

With the dirac spike and slab and ridge regression estimators minimizing the same sum-of-squares, the key difference with the constrained estimation formulation of ridge regression is not in the explicit form of *λ* but in what is bounded the domain of acceptable values for *α*. For the BayesR estimator the domain is specified by a bound on the *l*_0_*norm* of the regression parameter, while for its ridge counterpart the bound is applied to the squared *l*_2_*norm* of *β*. Multicollinearity will reduce the likelihood of the second covariate entering the model as it’s inclusion is dependent upon 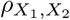 the correlation among covariates and 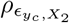 the correlation of the second marker and the residual vector after backfitting the first marker. This will limit the range of possible estimates to be lower than those obtained from ridge regression, reducing inflation of 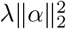 under high collinearity, but not entirely removing it. Due to the sampling of markers from a series of normal distributions collinearity will still inflate 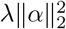, however, the degree to which this occurs will depend upon the number of correlated markers, the degree of correlation among them and the strength of the effects. Therefore, our aim here is not to derive a general solution predictive of all situations, merely it is to highlight that in order to make some inference as to the underlying distribution of genetic effects, it is required to extend the model as outlined in the following section.

### Extending the model to account for collinearity and genomic annotation

We extend the BayesR model to a BayesRR-RC model as follows

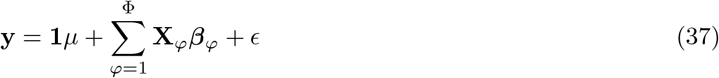

where there is a single intercept term **1***µ* and a single error term *ϵ* but now SNPs are allocated into groups (*ϕ*_1_, …, *ϕ*_Φ_), each of which having it’s own set of model parameters 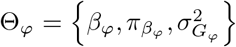. As such, each 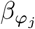 is distributed according to:

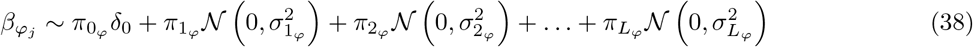

where for each SNP marker group 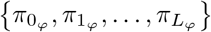 are the mixture proportions and 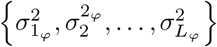 are the mixture-specific variances proportional to

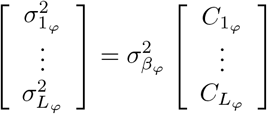

Thus the mixture proportions, variance explained by the SNP markers, and mixture constants are all unique and independent across SNP marker groups. This extends previous models (known as BayesRC [18] and BayesRS [19]), which have used additional mixtures for different SNP groups, but kept a single global variance component. Importantly, a single variance component with more mixtures serves only to change the amount of mass allocated at different sizes of the distribution, but does not alter the sizes of the effects themselves as there is still a single distribution. In contrast, the formulation presented here of having an independent variance parameter 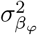 per group of markers, and independent mixture variance components, enables estimation of the amount of phenotypic variance attributable to the group-specific effects and enables differences in the distribution of effects among groups.

We can sketch the difference in the models by looking at the respective conditional posteriors, again, assuming a single component for simplification purposes. We have a BayesRC or BayesRS estimator by assuming different groups of effects in eq. 32, which yields:

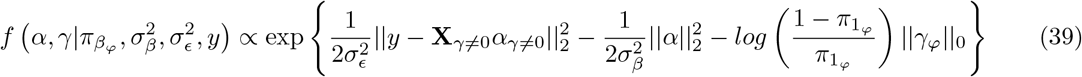

where 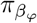 are the group-specific mixture proportions and ‖*γ*_*ϕ*_‖_0_ is the cardinality of the group. The corresponding MAP estimate would amount to adding extra penalisation on sparsity through the *π*_*ϕ*_ terms, while keeping the same level of shrinkage as the baseline BayesR.

In our model the conditional posterior is:

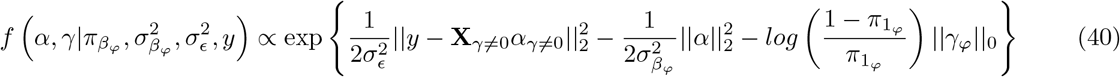

now each marker has a group-specific shrinkage 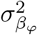, which translates to a specific *λ*_*ϕ*_ per group in the MAP estimate. This amounts to markers being shrunk according to the scale of the effects of their group, instead of the scale of all other markers. So instead of solving a single model selection and regularisation problem we are solving Φ model selection and regularisation problems, with shared information only through the residuals. If we subset by MAF and LD bins, the resulting groups of columns will have a correlation pattern similar to an exponential decay (LD decays with distance). If we take the whole genotype matrix, the pattern would be closer to a block diagonal matrix of correlations, in [17, 44] it is showed that the former case requires weaker conditions in order to recover the true vector *β* consistently than the latter. Although the sampling scheme was different, we have shown that a similar model with only two groups: genetic markers and epigenetic markers, is successful in identifying BMI and smoking epigenetic signatures [14].

### A Gibbs sampling scheme for biobank size data

For “*p* >> *n*” regimes, such as in genomics, where the number of covariates is greater than the number of individuals, hierarchical models controlling assumptions over the sparsity of the model are typically proposed, with examples of sparsity-inducing priors like the “spike and slab” prior [15, 25], the Bayesian LASSO [45] and the Horseshoe [46] prior. There are efficient tools to perform Bayesian regression analysis “out-of-the-box” using MCMC and variational inference [47–49], but these methods are limited to problems with explanatory variables in the low thousands of observations. Recent results show that Gibbs samplers for the Horseshoe prior [30], or for the Bayesian LASSO [50], offer a competitive advantage when combined with approximation schemes for problems of high dimensionality (over 100,000 covariates). These latter methods exchange the inversion of the coefficient matrix, for a matrix multiplication, thus reducing complexity from cubic to almost quadratic on the number of variables. However, despite these good properties, scaling these approaches up to a factor of millions of variables remains prohibitive.

We now describe an effective algorithmic implementation of our BayesRR-RC model that scales to millions of individuals, each genotyped at millions of genetic markers. We outline a Gibbs sampling algorithm that enables all sampling steps to utilize genetic data stored in mixed binary/sparse-index representation, reducing computational complexity of a single Gibbs step from 𝒪 (*n*) to 𝒪 (*n*_*z*_), with *n*_*z*_ the number of non-zero genotypes. We then outline a Bulk Synchronous Parallel Gibbs sampling scheme implemented based on a hybrid MPI + OpenMP model, distributing data across MPI tasks over as many compute nodes as required to hold all the data in memory. Uniquely, this enables large-scale genomic data to be split up into smaller manageable segments, whilst still conducting the analysis in the same way, estimated the marker effects jointly.

#### Algorithm 1

Serial Algorithm for sampling over the posterior distribution *p* (*µ, β, ϵ, σ*_ϵ_, *θ*). 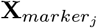 corresponding to the column *j* of the vector *marker*. Given that *marker* is shuffled before sampling the effects, this is equivalent to permuting the order of the effects to be sampled.

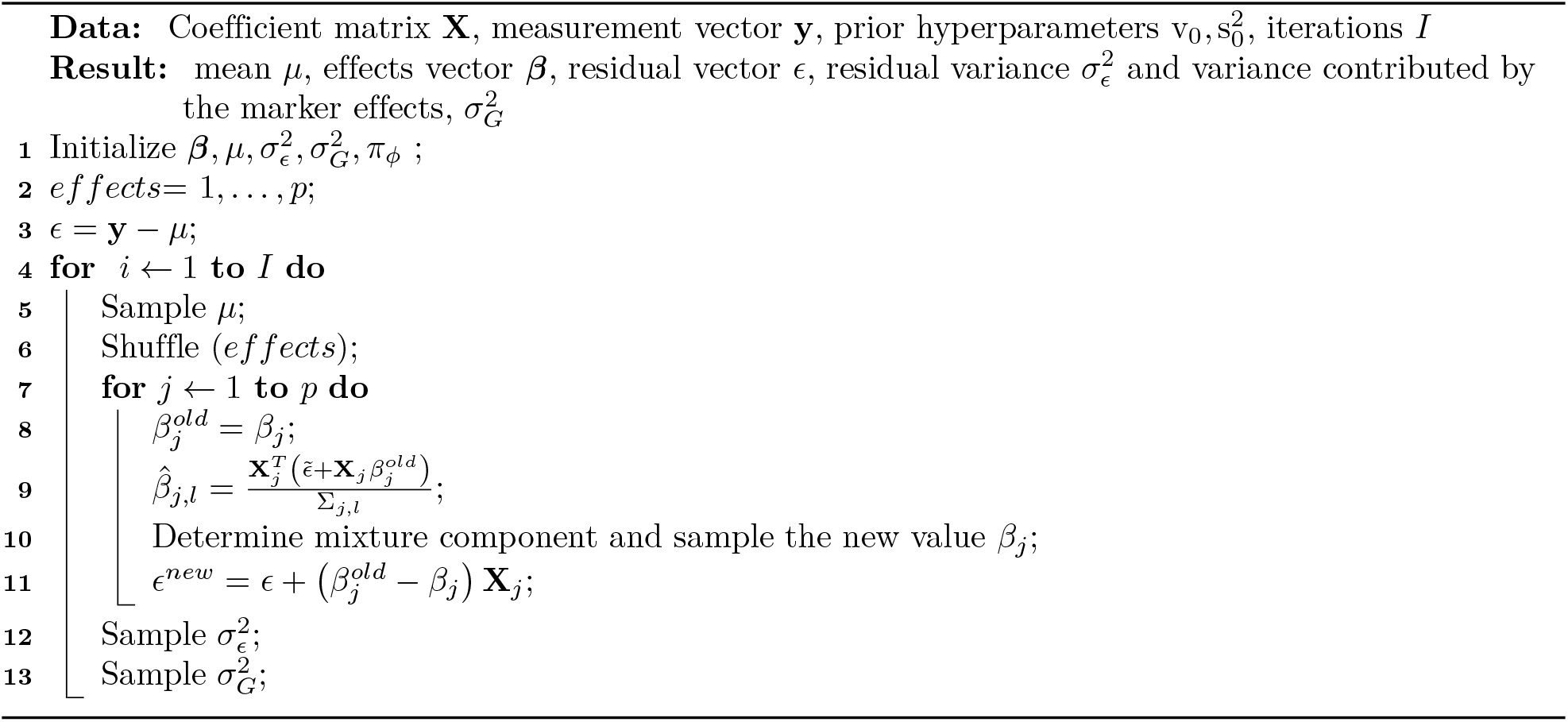

Algorithm 1 provides a full overview of the sampling scheme of the model as it has been previously implemented. For each marker *j*, we must compute 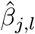 to determine which mixture a marker belongs to, before then sampling 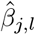 given the mixture group assigned. This quantity depends on the dot product 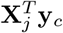, with **y**_*c*_ the centred phenotype. If we keep in memory the vector of residuals ϵ: = **y**_**c**_ − **X*β***_*γ*≠ **0**_, then we can compute efficiently 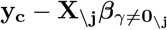 by the update 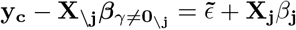, thus sampling from the joint distribution with a complexity 𝒪 (p). The most expensive operation in Algorithm 1 is computing the numerator in step 9: 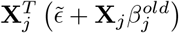. As the column vector **X**_*j*_ contains the centered and scaled genotypes, step 9 involves one sum of two dense vectors and a dot product of two dense vectors. However, if we store in memory the mean, *µ*_*j*_, and standard deviation *σ*_*j*_ of each column of the genotype matrix, we can express the numerator in step 9 with these quantities and the j-th column of the original genotype matrix **G** as (with 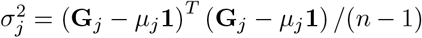 by definition):

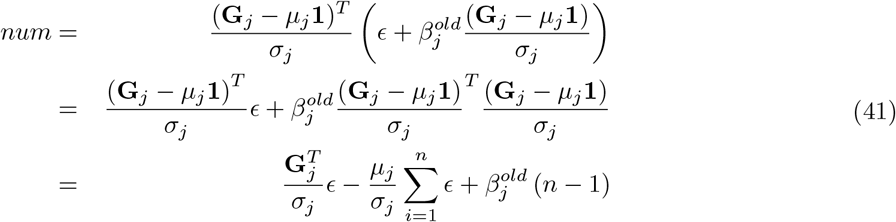

and we can do the same for the ϵ: update:

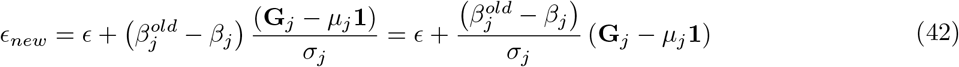

for which we only have to compute the difference of a sparse vector and a dense vector, and the sum of two dense vectors. Finally, to avoid computing 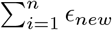 for each marker, we assign a variable to this quantity and update it after each *ϵ* update as follows (with 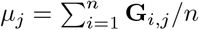 by definition):

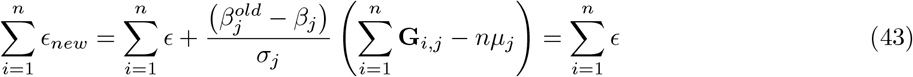

meaning that the sum of *ϵ* elements is constant during the algorithm execution (as expected as all involved vectors are zero-mean). Therefore, the only quantity to be computed per run (apart from the *ϵ* update) is the dot product 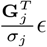 which can also be reduced, as the elements of **G**_*j*_ can only be either *{*0, 1, 2*}* with sequence data or hard-coded genotype. We call ℐ_1_ the indicator function such that 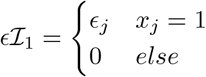 and similarly 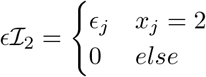 which then gives the dot product as 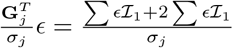 meaning that multiple 𝒪 (*n*) multiplications are now 𝒪 (*n*_*z*_) sums, and also that instead of storing in memory a sparse matrix of elements plus its indexes, we just need to store three ragged arrays of indexes, one for the “1” elements, a second one for the “2” elements, and a third one for the “M”issing elements. Those arrays contain information for all markers processed by a MPI task and are of unsigned integer type (32 bits). They store indices of the 1, 2 and M elements within the marker (i.e. ranging from 0 to *N* − 1). It corresponds to the smallest integer type that allows us to scale to hundreds of thousands or millions individuals. On top of those 3 ragged arrays there are two meta-data arrays for each element type which provide the starts and lengths of the 1, 2 and M elements for each marker in the ragged arrays. They are loaded in memory from reading sparse data files stemming from the conversion of the original Plink .bed file and accessed in parallel by the tasks with MPI I/O.

Even though the sparse representation is optimal in number of operations, performance may vary depending on hardware as a vectorised dot product may be faster than sparse dot product. Spatially, the sparse representation is optimal as long as the columns are sparse. In genotype data, even though the expected number of non-zeros per column is given by the average MAF (~20% in the UK Biobank data described below), the distribution is long tailed (Figure S7). These columns at the tail of the distribution can dominate the total size of the data structure in memory. Encoding a single column has a constant size of *N* × 2 bits in plink’s .bed file format (referred from now on as binary format), while in sparse representation a column has varying size of *n*_*z*_ × 32 bits. If we encode the columns with less than 6% of non-zeros as sparse and the rest in the original binary format, we can have a total memory occupancy of 60% the size of the original genotype matrix in Plink bed format. In (Figure S7) we represent on panel (b) the distribution of the proportion non-zeros per column of a genotype matrix for ~4 × 10^5^ individuals and ~ 1.5 × 10^7^ SNPs, solid line representing the mean of the distribution and slashed line the median. In panel (c) we show the total size of the data in memory as a function of the threshold used to split between binary and sparse format, in purple we see how the binary representations dominates the total size up until the mean of the distribution, after which, the size of the sparse data structure starts to dominate and ends up being around four times bigger than the original .bed file size (dotted horizontal line). We found the optimal threshold to be around 0.064 (6.4%, Figure S7).

Finally, we implement a vectorized dot product for genotype data stored in the raw binary format based on a couple of look-up tables, by writing the dot product as:

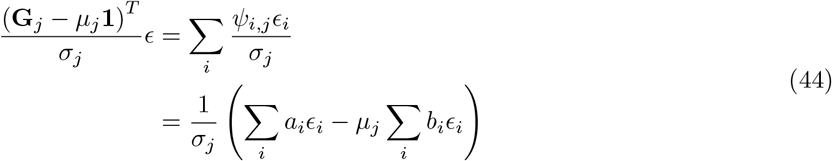

with coefficients *a*_*i*_ and *b*_*i*_ being 0.0, 1.0 or 2.0 depending on the value of **G**_*i,j*_ and following Table 4.

**Table 4.**
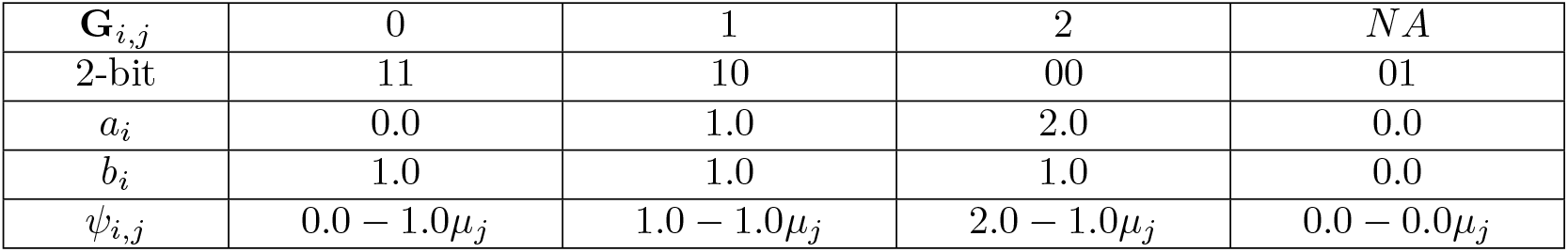
*a* and *b* coefficient values used for building up the two look-up tables needed for the vectorization of the dot product computation when processing binary data.

As 1 byte of plink’s .bed can contain 4^4^ = 256 different combinations of information for 4 individuals, we can setup two lookup tables with 256 × 4 entries each that will give for any byte the corresponding 4 *a*_*i*_ and *b*_*i*_ coefficients, hence allowing for vectorisation of Eq. 44 by performing *a*_*i*_*ϵ*_*i*_ and *b*_*i*_*ϵ*_*i*_ and accumulating them for 4 individuals at once. Additionally, we use OpenMP to parallelize the loop over the marker’s bytes. This greatly extends previously proposed sparse residual updating schemes and also facilitates the synchronous, fully parallel bulk-synchronous Gibbs sampling scheme that we describe in the next section below.

### Bulk-synchronous parallel Hogwild Gibbs sampling with sparse data

Bulk-synchronous parallel Hogwild Gibbs sampling [51] assigns block of columns from **X** to workers that then sample from *f* (*β*_*j*_|***β***_\*j*_, **y**) for each of the columns in their block. Workers can communicate between each other exchanging the current values of the variables they are sampling, or the whole state of variables for workers in particular. If we perform global synchronisation steps the algorithm is called Bulk-synchronous parallel Hogwild (BSP), if on the other hand, workers exchange messages without a global synchronisation, the algorithm is called Asynchronous parallel Hogwild (ASP) [52].

#### Algorithm 2: Hogwild Gibbs with ’Δ*ϵ*-exchange’.

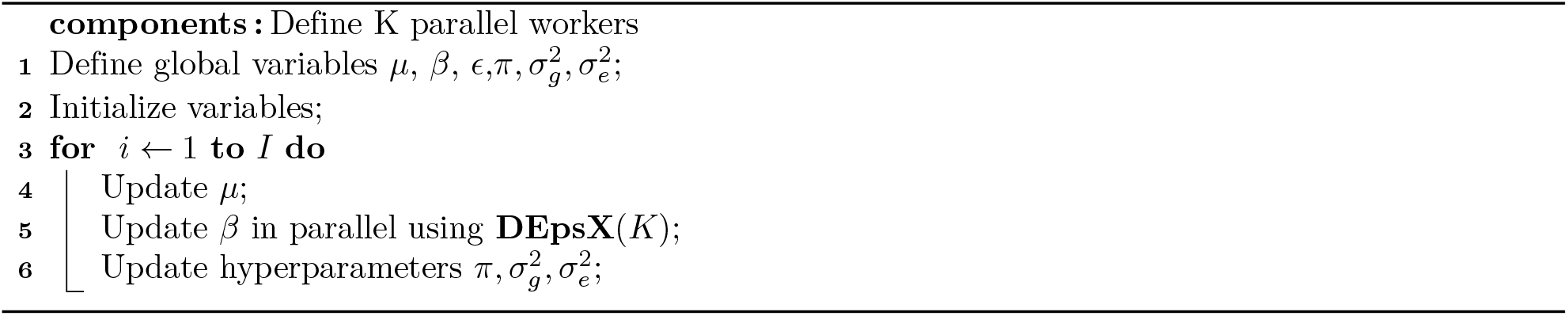

We propose Algorithm 2, which is a modification of a BSP algorithm where we sample the individual coefficients in parallel conditioned on the hyperparameters. We assign workers (MPI tasks) subsets of coefficients to sample, and each worker performs local Gibbs steps until a global synchronisation is triggered. This global synchronisation happens many times in each iteration, during the phase in which we sample the individual coefficients *β*_*j*_. For this algorithm, we developed a synchronisation scheme called ’Δϵ-exchange’as outlined in Algorithm 3. In this scheme each individual worker is assigned a block of columns from **X** and is in charge of sampling from *f (β*_*j*_ |***β***_\*j*_, **y)** for each of the columns in its block. We add an additional parameter for the synchronisation rate Ω. After Ω columns have been sampled in all workers (around 5-10 in practice to avoid divergence occurring), a synchronisation move is executed.

The purpose of the synchronisation move is to update all of the workers’ state based on the coefficients sampled from *t* = 1 until *t* = Ω in all workers. The sufficient statistic for this state is contained in the residual vector *ϵ*. Thus from *t* = 1 until *t* = *ω* each worker computes *f* (*β ϵ*) and keeps track of its local change in *ϵ* which we denote 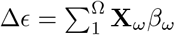 for *ω* in the set of indexes for the current batch of variables in the workers list of variables. For the synchronisation step, we use the MPI_Allreduce collective, meaning that each task will receive the sum of locally accumulated Δ*ϵ* from all tasks to update its 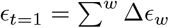 for *w* = (1…*W*) workers. With the new ϵ:_*t*=1_, the worker proceeds to sample the next Ω-sized batch of columns from its set of columns. This synchronisation scheme allows workers to exchange state information in compact form, as the total size of memory occupied in total by the messages is 𝒪 (*NW*).

#### Algorithm 3

’Δ*ϵ*-exchange’for synchronising changes in backfitted residuals in our BSP Gibbs sampling algorithm.

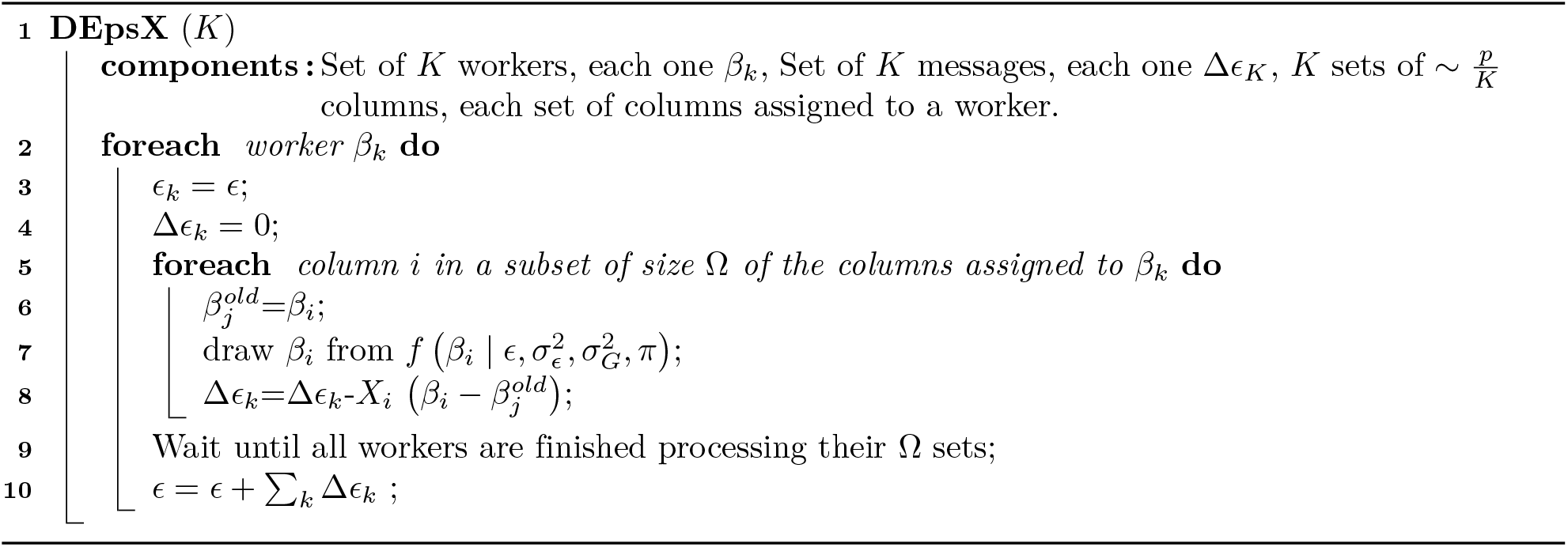

Previous results point to BSP Gibbs sampling for a multivariate Gaussian converging if the covariance matrix is strictly diagonal-dominant [52] with zero covariance of the markers split across workers. The risk for genomic data, is that two markers in LD get updated at the same time in parallel, double counting their effects, and leading to *ϵ* being mis-estimated after a synchronization has occurred. Suppose we have one fixed causal marker and two other markers *i* and *j* that are assigned to different MPI tasks. Suppose that the Pearson correlation between the causal marker and marker *i* or *j* is *ρ*_*i*_ and *ρ*_*j*_, respectively. Finally, let *ρ* denote the correlation between the markers *i* and *j*. For simplicity in this example suppose that the inclusion probability of the causal marker is *q* and we make an assumption that the inclusion probability of the marker *i* is then *P* (*β*_*i*_ ≠ 0) = *qρ*_*i*_ and for marker *j* it is *P* (*β*_*j*_ ≠ 0) = *qρ*_*j*_, that means that the inclusion probability is proportional to the correlation between causal and other markers. In reality, the effect size estimate is actually proportional to the causal effect: 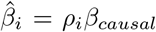 and the function between posterior inclusion probability and causal effect size *q*(*β*_*causal*_) is not linear for *β*_*causal*_ ≥ 0 as described in Eq.(12) and thus we cannot assume that *P* (*β*_*i*_ ≠ 0) = *qρ*_*i*_ in practice. In the case of parallelising the markers between two tasks we are interested in the probability that two markers from different tasks will absorb the effect of a same causal variant. Thus, we are interested in the probability *P* (*β*_*i*_ ≠ 0, *β*_*j*_ ≠ 0|*i, j* ∈ *U*), where *U* is the set of markers that are updated simultaneously in two different tasks. Thus, we can write:

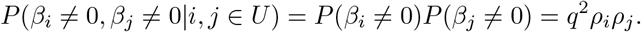

We see that the probability of making a mistake is dependant on the product *ρ*_*i*_*ρ*_*j*_. The correlation matrix *R* of the three markers

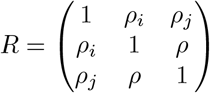

has to be positive semi-definite and thus we can examine what are the possible values for the product *ρ*_*i*_*ρ*_*j*_ given that we know *ρ*. Note that the value of *ρ* can be controlled by providing some blocking mechanism that would assign SNPs to the tasks so that the correlation for the markers from different tasks would be limited to *ρ* and this is what we advocate here, placing contiguous blocks of markers into different tasks, so as to maximise the LD within a block (MPI task), but minimise the LD across blocks. The maximum possible values for the product follow a linear function that depends on *ρ* as

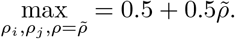

To get better estimates for the constraints for the product *ρ*_*i*_*ρ*_*j*_ then we need to make further assumptions about the distribution of *ρ*_*i*_ or *ρ*_*j*_. Therefore, we can say that *P* (*β*_*i*_ ≠ 0, *β*_*j*_ ≠ 0|*i, j* ∈ *U*) ≤ *q*^2^(0.5 + 0.5*ρ*). This result and inequality only holds per sampled pair (*i, j*). We then multiply this result with the probability of sampling the pair (*i, j*) that both have correlations *ρ*_*i*_, *ρ*_*j*_ > 0. Denoting a set of markers that have a positive correlation with one specific causal marker as the causal radius *C*, The probability of sampling any pair (*i, j*) is

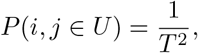

where *T* is the number of markers per one task. The probability of pair (*i, j*) belonging to *C* is *P* (*i, j* ∈ *C*) = *c*(<< 1), some reasonable values could be proposed or estimated for this (for example,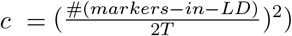. Combining the results together we get that the probability of making a mistake at one update of a pair (*i, j*):

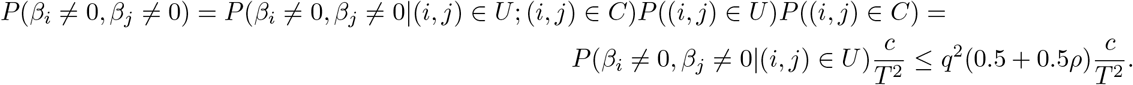

This result goes for one fixed causal marker and it also represents the expected number of mistakes per sampled pair (*i, j*) for one causal marker. If we want to find the expected number of mistakes per sampled pair, we should sum across the *P* causal markers:

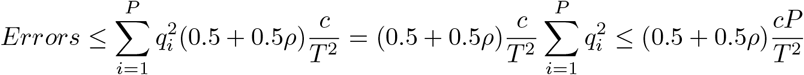

To provide some intuition, we can think of an extreme scenario and assume that there are 100,000 variants in the SNP marker data that would enter the model as they are in LD with underlying causal variants, that each of these variants has posterior inclusion probability of 1, and that for each variant there are two blocks with 30,000 markers in total of which 100 markers have LD = 1 with the causal variant, and that both blocks contain 30,000 markers. Placing these values into what we derive above and sampling over 10,000 iterations leads to probability of an error ~ 0.1 throughout the sampling for this extreme example. Having derived a stable highly parallel Gibbs sampling algorithm for large-scale genomics data, we then performed exhaustive empirical validation of our algorithm in simulation study as described below.

### Implementation and processing setup

We implement algorithms 2 and 3 in C++ as a pure CPU MPI + OpenMP hybrid solution. All data structures were properly aligned in memory to assist vectorization and assembly code was examined to ensure that the code was properly vectorized where expected. We utilize the scientific libraries eigen and boost (see Code Availability) and we profiled and benchmarked the code with Intel performance analysis tools such as Advisor and Ampflier. Current implementation requires to be compiled with Intel compiler on an architecture supporting at least AVX2 although support for AVX512 is recommended for performance. The code is freely available from our Github repository and we also provide a statically compiled binary (see Code Availability). All the results were generated on the cluster Helvetios from EPFL (see Code Availability) using 10 compute nodes and setting 8 MPI tasks per node and dedicating 4 (physical) cores to each task. 10 is the minimal number of nodes that was required to hold all the data in memory in its mixed-representation.

### Posterior summaries and discovery

The ability of the additive regression model outlined and applied here to infer the underlying distribution of genomic effects is limited unless an additive model with many 0 coefficients holds as approximately true and the true number of underlying nonzero coefficients is << *n*. Various ad hoc penalty functions in machine learning, and the range of proper priors employed by members of the Bayesian alphabet and beyond, all impose a restriction on the size of the regression coefficients, and while these restrictions differ, they all provide shrinkage estimators that by their definition are biased as they are shrunk toward zero (this true of mixed-linear association models also). In other words, the penalty function (prior) will be important and will influence the inference made here. Thus, the inference we obtain can only be made with respect to our *a priori* assumption that many marker effects are zero, and that the effects of those that are not zero can be reflected by a mixture of zero centred Gaussian distributions. Given this, we focused on comparing the posterior distributions of different traits obtained under the same model, focusing on the hyper-parameter estimates obtained for MAF-LD-annotation groups, and comparing these across traits. It has been shown in Bayesian penalized regression models that what is learned about *β* is a function of what is learned about **X*β*** and thus by placing separate hyper-parameters over different genomic groups we can obtain inference as to the variance contributed by each group [53]. As we show through theory and simulation study described below, MAF-LD-annotation specific hyper-parameters likely results in improved inference as to the distribution of genetic effects. However, with the exception of very rare variants with LD ~ 0, we cannot treat each *β*_*j*_ as independent and thus here we outline a strategy to identify associated genes, or genomic regions within a probabilistic framework.

For a simple example, consider two markers in LD that are correlated with a single causal variant, where either or both markers may be in the model at any one iteration and the expected posterior inclusion probability of each SNP is 0.5. In this scenario, we cannot use the posterior inclusion probability of each marker to assess association and thus instead, we take an approach of assessing the contribution of different genomic regions to trait variation whilst controlling the posterior type I error rate (PER), which is more suitable controlling for false positives, than controlling the genome-wide error rate (GER). Many papers have discussed the advantages of controlling the false discovery rate (FDR), and related measures rather than controlling GER [54] and here we follow [22] where the posterior probability that *β*_*j*_ is nonzero for at least one SNP *j* in a window or genomic segment is used to make inferences on the presence of an association in that segment.

Briefly, following [22], we will refer to this probability as the window posterior probability of association (WPPA). The underlying assumption is that if a genomic window contains a marker in LD with a causal variant, one or more SNPs in that window will have nonzero *β*_*j*_. Thus, WPPA, which is estimated by counting the number of MCMC samples in which *β*_*j*_ is nonzero for at least one SNP *j* in the window, can be used as a proxy for the posterior probability that the genomic region contains a causal variant. Because WPPA for a given window is a partial association conditional on all other SNPs in the model, including those flanking the region, the influence of flanking markers on the WPPA signal for any given window will be inversely related to the distance *k* of the flanking markers. Thus, as the number of markers between a causal variant and the focal window increases, the influence of the causal variant on the WPPA signal will decrease and so WPPA computed for a given window can be used to locate associations for that given window [22].

This measure can be shown to control the PER, which in frequentest statistics would be associated with the test of a hypothesis. The null hypothesis in this case is that the genomic region does not contain any SNPs associated with the trait. Using this notation, WPPA is the conditional probability that the null is false given the observed data, while PER is the conditional probability that the null hypothesis is true given that it has been rejected based on some statistical test. Suppose the test is based on WPPA and the null is rejected whenever WPPA is larger than some value *t*. Then, PER is the probability that that the null hypothesis is true given WPPA is larger than *t*, and it can be written as:

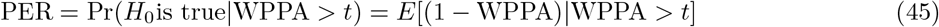

Thus, for any interval with WPPA > *t* the proportion of false positives among significant results will be ≤ (1 − *t*). Here, we are interested in detecting genes and genomic regions that explain more than some proportion *v* of the total phenotypic variance attributable to the SNP markers (genetic variance). The genomic segment variance is defined as the sum of the squared partial regression coefficient estimates at each iteration and these are divided by the sum of all the squared partial regression coefficient estimates genome-wide to give a proportion for each genomic region at each iteration. Then we simply count the proportion of MCMC samples where the proportion of genetic variance is greater than a thresholds of 0.001% and we denote this metric as the posterior probability of window variance (PPWV).

We extend this PPWV approach to develop an association metric for LD blocks of the genome. Currently, association studies predominantly estimate SNP effects and test for association one marker at a time, which does not control for local LD among SNP markers. Thus, the level of association determined is at the regional level as results are reduced, using LD patterns, to a subset of the strongest associated LD-independent variables. We can solve the problem of having a correlated posterior distribution by applying our PPWV approach the LD blocks of the genome providing a Bayesian probabilistic metric of association that is equivalent to selecting the number of independent associated SNP markers. We define LD blocks as a group of SNPs that have squared correlation greater that 0.15 and then for each iteration, we sum the squared partial regression coefficient estimates for all the SNPs within the block, divide this by the sum of all the squared partial regression coefficient estimates genome-wide to give a proportion for each genomic region at each iteration. Then we simply count the proportion of MCMC samples where the proportion of genetic variance is greater than a thresholds of 0.001% providing a probabilistic association metric for each LD block that controls the FDR genome-wide. Within each associated region the individual SNP posterior inclusion probabilities can then also be used to “fine-map” the associations, in order to select the base-pair position that is most likely to be closest to the true underlying causal variant in imputed SNP data.

### Simulation study

#### Large-scale simulation study

We wished to highlight the performance of BayesRR-RC for the following tasks: (i) estimation of the phenotypic variance attributable to genetic markers, both genome-wide, and for segments of the genome; (ii) “discovery” of associated genomic regions, including the identification of candidate SNPs; and (iii) phenotypic prediction. We developed a large-scale simulation study, randomly selecting 40,000 unrelated individuals and all 596,741 imputed (version 3) genetic markers from chromosomes 19 through 22 from the UK Biobank. Using this data, we simulated a wide-range of different possible underlying genetic effect size distributions as follows:

- We chose either 5,000 or 10,000 imputed SNP markers for which to assign a genetic effect size, providing two different levels of polygenicity.
- We selected these 5,000 or 10,000 markers in two different ways. Either, we selected SNPs at random, or we selected the marker of highest minor allele frequency per LD block of the genome, with an LD block defined as a group of SNP markers with absolute LD of at least 0.05. Randomly allocating markers creates a set of associated variants with the same distribution of LD and MAF as the SNP data, which is composed of predominantly low frequency variants. Selecting only the highest frequency marker per LD block creates a simulation setting where for each set of markers in LD with each other, there is only one causal genetic variant, and where the distribution of associated markers differs to that of the SNP markers as a whole.
- Having created four different ways of selecting associated markers (5,000 or 10,000 and high-MAF or random) we then created five different ways of assigning effect sizes to them:
  – We simulated effect sizes from a normal distribution with zero mean and variance 0.6 divided by the number of markers (5,000 or 10,000) with no relationship to the LD or MAF of the markers. Thus, effects had variance *α N* (0, *w*^0^[*p*(1 − *p*)]^0^) with *w* the LD score of the marker and *p* the allele frequency.
  – We simulated effect sizes from a normal distribution with zero mean and variance 0.6 divided by the number of markers (5,000 or 10,000) *α N* (0, *w*^−0.25^[*p*(1 − *p*)]^−0.25^). This simulates stronger effect sizes for rare variants and those in low LD.
  – We simulated effect sizes from a normal distribution with zero mean and variance 0.6 divided by the number of markers (5,000 or 10,000) *α N* (0, *w*^0.25^[*p*(1 − *p*)]^−0.25^). This simulates stronger effect sizes for rare variants and those in high LD.
  – We simulated effect sizes from a normal distribution with zero mean and variance 0.6 divided by number of markers (5,000 or 10,000) *α N* (0, *w*^−0.25^[*p*(1 − *p*)]^0.75^). This simulates equivalent effect sizes for common and rare variants, and greater effects for markers in low LD.
  – We simulated effect sizes from a normal distribution with zero mean and variance 0.6 divided by the number of markers (5,000 or 10,000) *α N* (0, *w*^0.25^[*p*(1 − *p*)]^0.75^). This simulates equivalent effect sizes for common and rare variants, and greater effects for markers in high LD.
- For each of the four different sets of markers, each with 5 different effect size sampling schemes, we then created two additional settings. In the first setting markers were sampled from the various normal distribution, as described above, for the 5 different effect size sampling schemes. In the second setting, for each of the 5 different effect size sampling schemes we simulated effects from 13 different distributions, one for each of 13 different genomic annotation groups with different proportions of total SNP heritability 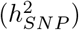. So, for each of the 5 different effect size sampling schemes the relationship to LD and MAF remained the same, but the total variance attributed to the SNP markers was partitioned across annotation groups as follows for exonic variants 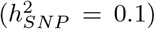, intronic variants 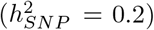, 1kb promotor variants 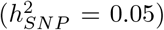, 1-10kb enhancer variants (0.025), 1-10kb transcription factor binding sites 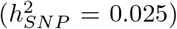, 1-10kb other variants 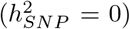, 10-500kb enhancers 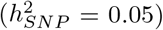, 10-500kb transcription factor binding sites 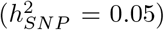, 10-500kb other variants 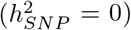, 500kb-1Mb enhancers 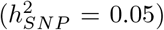, 500kb-1Mb transcription factor binding sites 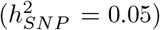, 500kb-1Mb other variants 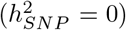,and other non-annotated SNPs 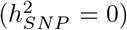. Four of these distributions had zero variance indicating that no associations were created for these groups. In the first setting, this simulates variance explained by annotation groups that is on average proportional to the number of SNPs within each annotation. In the second scheme, the variance and average effect size differs across annotation groups. We refer to these as 2 different enrichment settings: “random”, or “enriched”.
- This created four different sets of associated markers (5,000 or 10,000 and high-MAF or random), each with 5 different marker effect size sampling schemes, which we refer to in the main text as the 20 different generative genetic models (Table 1), each of which has 2 enrichment settings. This gave 40 different sampling schemes for the genetic effects and we simulated 10 replicates for each setting, giving a total set of 400 simulated phenotypes.
- For each generative model the total genetic variance was 0.6 and we sampled individual-level environmental (residual) variance from a normal distribution with zero mean and variance 0.4 to give phenotypes with zero mean and unit variance.

This range covers generative genetic models discussed in the literature and provides models that both fit and violate the assumptions of the range of variance component statistical models, both individual-level and summary statistic approaches, that are currently applied in the literature for estimation of the variance attributable to the SNP markers, for testing association of genetic markers with phenotypes genome-wide, and for genomic prediction. Therefore, this provides a range of different simulation scenarios for which we can explore the model performance of BayesRR-RC and then to compare to existing approaches.

We began by comparing BayesRR-RC to BayesR. For each of the 400 phenotypes, we ran both models for 5,000 iterations with burnin of 2000 iterations. To assess performance for estimation of the phenotypic variance attributable to genetic markers, we calculated the posterior mean of the phenotypic variance attributed to the SNP markers and for BayesRR-RC, we calculated the posterior mean of the phenotypic variance attributed to the SNP markers within each annotation group.

To assess performance for “discovery” of associated genomic regions, we first wished to show that the effect sizes are accurately estimated. For each generative model, we first calculated the correlation of the posterior mean effect size of the SNP markers within each annotation group to the true mean effect size of the SNP markers simulated as being trait-associated. We summed the squared regression coefficient estimates of all SNPs in the model at each iteration for each LD block of the genome, with an LD-block defined as markers in LD *R*^2^ ≥ 0.1 within each simulation replicate and took the posterior mean across iterations. We then calculated the z-score of the LD block squared regression coefficient estimate from the simulated value and we use this metric to assess estimation accuracy across the allele frequency spectrum of the markers assigned as being trait-associated. Together, with the correlation of the average effect size obtained, this provides an assessment of effect size estimation accuracy of our proposed model.

We then test our PPWV approach described above for LD blocks of the genome. Our proposed PPWV approach provides a probabilistic determination of associated of a given LD block, genomic window, gene, upstream region etc. and while our theory outlined above suggests that the FDR should be well controlled we wished to confirm model performance in identifying associated regions of the genome. We calculated precision-recall curves, where associations are defined as LD blocks with PPWV of ≥ 95% at 0.001% proportion of variance explained. True positives were the number of identified 5,000 or 10,000 LD blocks that contained a causal variant. False positives were the number of identified LD blocks that did not contain a causal variant. Precision was defined as the ratio of true positives to the sum of true positives and false positives. Recall was defined as the ratio of true positives to the sum of true positives plus false negatives. The FDR was defined as the proportion of LD blocks with PPWV of ≥ 95% at 0.001% proportion of variance explained that did not contain a causal variant. In this way, we are able to determine the discovery performance and FDR control of our proposed PPWV LD block approach across a range of generative genetic models for both BayesR and BayesRR-RC.

Finally, we wished to determine the out-of-sample phenotypic prediction performance of BayesRR-RC. We randomly selected 10,000 individuals from the UK Biobank that were unrelated to those used in the simulation. Using the same SNP markers and the simulated marker effects we calculated a simulated genetic value for each individual across the replicates. Then, using the effects generated by BayesR and BayesRR-RC, we calculated the predicted genetic value and determined the correlation with the simulated genetic value.

### Benchmarking against existing approaches

Comparing frequentist and Bayesian model performance is generally not advisable, but benchmarking BayesRR-RC against existing approaches for the tasks (i) through (iii) described above is required to convey the model performance improvements possible under our sampling scheme as compared to current approaches. Thus, we ran the following individual-level models for each of the 400 phenotypes:

- A restricted maximum likelihood model implemented in the software GCTA with a single relationship matrix providing an estimate of the variance attributable to SNPs genome-wide.
- A restricted maximum likelihood model implemented in the software BoltREML [20]. Here, we used a 78 MAF-LD-annotation group model using the non-overlapping genomic annotation groups described below in the UK Biobank analysis providing an estimate of the variance attributable to SNPs genome-wide and an estimate of the variance attributable to SNP markers of each annotation group.
- A Haseman-Elston regression using the same 78 group model implemented in the software RHEmc [21], providing an estimate of the variance attributable to SNPs genome-wide and an estimate of the variance attributable to SNP markers of each annotation group.
- Mixed linear association model (MLMA), which is a two-stage approach where the variance attributable to the SNP markers genome-wide is estimated and this estimate is then used in a second generalized least squares step to test for SNP-phenotype associations one marker at a time. There are two forms of this model. In the first, the SNP is fitted twice as it is included in both the fixed and random terms (MLMAi). In the second, the SNP to be tested as fixed is removed from the random term alongside those on the same chromosome (MLMA). We used the software BoltLMM, Regenie, and GCTA to fit these models. These approaches provided estimates of the SNP regression coefficients (marker effect sizes).

For each of the 400 phenotypes, we also estimated the regression coefficients of each marker using plink2 [55], whilst fitting 20 principal components of the marker data as covariates. We then ran the following summary-statistic models:

- Linkage disequilibrium score regression (LDSC [11]), with LD scores calculated using the same data, and the same 78 non-overlapping annotations in a 78 component LDSC annotation model. We included SNPs with MAF>1% following the software instructions. This model is intended to approximate an individual-level REML analysis with 78 annotations and provides an estimate of the variance attributable to SNPs genome-wide and an estimate of the variance attributable to SNP markers of each annotation group.
- We used the software SumHer [6]. We calculated marker taggings under the same 78 component annotation model. We ignored the LD weights when calculating the taggings as we found this gave the best estimates we could obtain from the simulated data across all scenarios. We set the relationship of effect size and minor allele frequency to be −0.25 as suggested by the authors. This matches some settings, but differs to others, and this is important as in real data one does not know the true underlying model and we wished to test how this specification influences model performance. This model is intended to approximate an individual-level REML analysis with 78 annotations, but using a different scaling of the relationship matrix, and provides an estimate of the variance attributable to SNPs genome-wide and an estimate of the variance attributable to SNP markers of each annotation group.

We ran the following prediction models focusing on the models proposed in a recent paper [24], as the methods contain two approximations to our BayesRR-RC model and the authors claim to outperform all other existing methods, including the individual-level model:

- An individual-level bayesR model using genomic annotation SNP variance estimates from the SumHer models as implemented in the software MegaPRS [24]. This provides estimates of the SNP marker effects for creating a genetic risk predictor.
- An individual-level boltREML model using genomic annotation SNP variance estimates from the SumHer models as implemented in the software MegaPRS [24]. This provides estimates of the SNP marker effects for creating a genetic risk predictor.
- A summary statistic bayesR model using genomic annotation SNP variance estimates from the SumHer models as implemented in the software MegaPRS [24]. This provides estimates of the SNP marker effects for creating a genetic risk predictor.
- A summary statistic boltREML model using genomic annotation SNP variance estimates from the SumHer models as implemented in the software MegaPRS [24]. This provides estimates of the SNP marker effects for creating a genetic risk predictor.

We compared the estimated phenotypic variance attributable to the SNP markers across approaches to the simulated value both genome-wide and for each annotation group. This represents a direct comparison of individual-level and summary statistic approaches, across different generative genetic models for the same genomic annotations. We determined the ability of the approaches to correctly identify enriched regions of the DNA by estimating the correlation of the estimated and simulated average effect size for each annotation group. For LDSC, BoltREML, RHEmc and SumHer we calculated the average effect size as the variance explained by SNP markers of an annotation divided by the number of SNPs of the annotation as proposed in the literature (Figure 1).

We compared the accuracy of the effect size estimation across approaches by estimating the z-score of the estimated from the true simulated value for each SNP marker effect. This provides a single comparable metric of effect size estimation accuracy across Bayesian and frequentist approaches, and we then plotted these z-score against the minor allele frequency of the causal variant of the block. Our theory predicts that these z-score should be positively inflated at higher minor allele frequencies for the MLMA approaches when many causal variants are high-MAF SNPs (Figure 2a).

Then, to compare the Bayesian and frequentist approaches in their discovery performance, we used precision-recall curves and we also compared the false discovery rate. For the MLMA methods, following standard practice we clumped the marker effect estimates using Plink, as local LD is not controlled for, selecting LD independent markers (LD *R*^2^ ≤ 0.01 with other markers) across the genome. True associations were defined as clumped selected SNPs that were in LD with a simulated causal variant (LD *R*^2^ ≥ 0.01). False associations were defined as selected SNPs that were not in LD (LD *R*^2^ ≤ 0.01) with a simulated causal variant. Precision and recall were calculated across thresholds of the chi-squared statistics of the selected markers, and the area under the curve was calculated using the trapezoid rule for calculating the integrals, assuming the curve is linear between the points. FDR is then calculated as the proportion of markers with p-value ≤ 5×10^−8^ that were not in LD with a causal variant (LD *R*^2^ ≥ 0.01). This provides a way to directly compare model performance for the discovery of associated genomic regions across Bayesian and frequentist approaches and tests our hypothesis that a PPWV approach controls FDR well in comparison with Bonferroni p-value correction (Figure 2b,c). For both MLMA and Bayesian approaches our definition of FDR is not strictly the FDR. Markers in LD *R*^2^ ≤ 0.01 with the clumped selected markers may still show a weak correlation with the simulated causal variants, and likewise blocks of SNPs in LD *R*^2^ ≤ 0.1 may still be in weak LD with the causal variants. Our approach instead captures the ability of MLMA and Bayesian approaches to localise an effect to within *R*^2^ ≥ 0.01 and *R*^2^ ≥ 0.1 respectively.

We also focused on the ability of our approach to fine-map associated regions to identify candidate SNPs and to provide a probabilistic assessment of the most likely associated set of SNP markers. To do this we used our large-scale simulation data and focused on seven focal regions within a blocks of chromosome 22. We allocated effect sizes to the following SNPs: rs131529 with MAF 0.32 which had LD *R*^2^ ≥ 0.15 with 348 other SNPs, rs2096537 with MAF 0.14 which had LD *R*^2^ ≥ 0.15 with 295 other SNPs, rs131538 with MAF 0.05 which had LD *R*^2^ ≥ 0.15 with 82 other SNPs, rs141962840 with MAF 0.007 which had LD *R*^2^ 0.15 with 11 other SNPs, rs117873986 with MAF 0.02 which had LD *R*^2^ ≥ 0.15 with 12 other SNPs, rs9606483 with MAF 0.005 which had LD *R*^2^ ≥ 0.15 with 1 other SNP, and rs78881648 with MAF 0.009 which had LD *R*^2^ ≥ 0.15 with 1 other SNP. To these seven SNPs, we assigned the same effect sizes in four different scenarios, either 0.05, 0.025, 0.0125, or 0.01 on the SD scale. On the remainder of chromosomes 19,20,21 and 22, we randomly selected 1000 SNPs as causal variants to give a polygenic background, sampling their effects from a normal distribution with zero mean and variance 0.5/1000. We repeated each of the four scenarios 20 times. We selected these regions as we wished to compare the performance of BayesRR-RC to the fine-mapping approach SuSiE as outlined in a recent paper [23]. For BayesRR-RC, we calculate the PPWV of the LD blocks containing the seven focal SNPs, and then prune these blocks based on the LD among the markers in the block (described as “purity” in the SuSiE paper [23]) to identify a credible set with LD *R*^2^ 0.9. We then count the proportion of times across the simulations that each causal variant was contained with one of the credible sets. For SuSiE, we ran the model from the individual-level data of the whole block of chromosome 22 using the suggested settings and setting K=10. We then calculate the proportion of times that the credible sets identified contained one of the seven causal variants. We present these results in Figure S4c.

Finally, we took the marker effect estimates from the MLMA approaches and conducted LD clumping with p-value thresholding using Plink to find the set of markers that gave the highest correlation of the genetic predictor and the simulated genetic value within the 10,000 UK Biobank individual selected for out-of-sample prediction. We also used the MegaPRS methods implemented in the software LDAK running the four different models described above. We compared the correlation of predicted and simulated genetic value across approaches for each of the 400 simulated phenotypes (Figure 2d).

### Smaller-scale simulation examples

While we assess the performance of our model in the large-scale simulation work, smaller-scale focused simulation work was also conducted to support and test the inference made. Our theory and our large-scale simulation study suggests that there will be increased variance of the regression coefficient estimates and, as a result, an inflated estimate of the phenotypic variance attributable to SNP markers under high multicollinearity for both mixed linear model approaches and a Dirac spike and slab mixture model. To create a toy example of this, we conducted a simulation study where for each of 50 replicates, we simulated 50 independent genomic regions, each containing two SNP markers. In each simulation replicate, we simulated values for 5,000 individuals at each of the 50 SNP marker pairs, by first simulating from a standard multivariate normal distribution with correlation set to either 0 or 0.99. From this, we obtained the integral from − inf to *q* of the probability density function, where *q* is the z-score of the values obtained for each individual from the multivariate normal. From these integrals, we then made two draws from the inverse of the cumulative density function of the binomial distribution to obtain the marker value for each individual, with frequency 0.3. This gave marker values (0, 1, or 2), with the pairs of SNPs having either all LD = 0, or all LD = 0.99. For each of the 50 pairs of SNPs, we assigned effect size 0 to the first marker and 0.1 to the second marker. We then scaled the SNP markers to zero mean and unit variance and multiplied the markers by the effect sizes to obtain the genetic values for the 5,000 individuals, with variance 0.5. We then simulated the environmental component of the phenotype from a normal distribution with zero mean and variance 0.5 and then created a phenotype as the sum of the genetic values and the environmental values, with zero mean and unit variance.

We then analysed these 50 data sets using different methods of single-marker OLS regression (OLS), mixed-linear model association (MLMA), ridge regression (Ridge), and a Dirac spike and slab mixture of regressions model (BayesR), all of which are described above. For the frequentist approaches, we directly solved the estimation equations, scaling the SNP markers to have zero mean and unit variance. For BayesR we sampled the effects for 5000 iterations, with burn-in period of 2000 iterations to obtain the posterior mean effect sizes, again scaling the SNP markers to zero mean and unit variance. We repeated these analyses many times, each time fixing the estimated phenotypic variance attributable to the markers 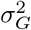 to be a different value. We selected (2, 1, 0.5, 0.1, and 0.01) and fixed the residual variance 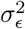 to be 0.5, to give different lambda values 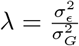, giving *λ* = 0.25, 0.5, 1, 5, and 50. Our aim here was to explore the pattern of effect sizes that we obtain under these *λ* values. So first, we plotted the effect sizes obtained for each of the 50 SNP pairs obtained across the 50 simulation replicates in Figure S2, to show the differences in the variance of the estimates obtained across approaches when the pairs of SNP markers were orthogonal (LD=0), or collinear (LD=0.99), under different lambda values. Second, we then plot the distribution of the sum of the squared regression coefficients in Figure S1c across approaches, when the pairs of SNP markers were orthogonal (LD=0), or collinear (LD=0.99), under different lambda values, where the expectation is 0.5 (sum of the 50 squared 0.1 SD effect sizes). This simulation confirmed, that regression coefficient estimates have higher variance under multicollinearity, resulting in inflation of the sum of the squared coefficient estimates for all approaches when the variation attributable to SNP markers is overestimated, resulting in a reduction in the lambda values.

We then further explored the performance of the MLMA and BayesR models under multicollnearity to (i) better understand the interplay between the fixed GLS estimate obtained and the random marker effects, and (ii) to better understand how the prior of the BayesR model changes with lambda and how this constrains the inclusion probabilities of correlated markers. We first examined the influence of varying lambda and varying the collinearity of markers on the variation of the effect size estimates obtained from the Henderson’s mixed model equations, where one focal marker is estimated as fixed, and a further five markers are estimated as random, with LD between the markers estimated as fixed and random. To do this, we simulated five markers in the same manner as described above that were either (i) entirely orthogonal with LD = 0, or (ii) had LD = 0.99 among the first three markers, with the final two markers having LD = 0 with all others. We assigned effect sizes to the five markers as *beta* = (0.25, 0, 0, 0.25, 0.25), multiplied these effect sizes by the simulated marker values scaled to zero mean and unit variance to create the genetic values, and then added an environmental component simulated from a normal distribution with mean zero and variance 1 minus the variance of the genetic values (0.1875) to give a phenotype with zero mean and unit variance. We directly solved the Henderson’s mixed model equations, fixing the lambda value at different levels (the appropriate lambda from theory assuming orthogonal covariate would = (1− 0.1875)*/*0.1875 = 4.333). We find that even with high shrinkage, a lambda value of almost 20 times greater than the theoretical orthogonal expectation is required to produce effect sizes under collinearity, with similar variance to those obtained under orthogonality (Figure S2).

For BayesR, we first explored the density of the posterior distribution by simulating draws from the prior as we change the variance attributable to the SNP markers. Figure S2c shows these densities, revealing how the prior becomes strongly centred on zero and almost exponentially distributed as the variance becomes small. This is in contrast to the almost flat prior observed with high variance, which will do little to constrain effect size estimates toward zero. We then conducted 1000 simulation replicates of paired SNP markers for 10 different scenarios of variance attributable to the SNP markers of 0.01, 0.05, 0.1, 0.2, and 0.5, for pairs of SNPs with correlation of either 0 or 0.99. For each of these 10,000 data sets we simulate a pair of SNPs for 5000 individuals, assuming error variance of 0.5, effect size for the first marker of 0.01 SD and then we simulated a sequence of 1000 different effect sizes from −0.05 to 0.05. Of these 10 million phenotypes and pairs of SNPs obtained, we then determine the posterior inclusion probability of the second marker, given that the first marker is in the model, with the effect size correctly estimated as 0.01, from the BayesR model derivations presented above. The lines presented in Figure S2d go through the mean posterior inclusion probability of the second SNP marker across the 1000 simulation replicates, for each of the 1000 different effect sizes from −0.05 to 0.05 for marker 2, with a different colour for each scenario of the variance attributable to the SNP markers. The plot shows a reduction in the posterior inclusion probability of the second SNP marker as the variance attributable to the SNP markers decreases under multicollinearity. Thus, if the hyperparameter estimates of the variance contributed by markers is kept small, by having different hyperparameters for different groups of markers, then the BayesR model acts to constrain the inclusion of any additional correlated markers in the model.

Having confirmed our theory, we then conducted a further simulation study to further replicate these observations using real genomic data. We randomly selected 50,000 individuals from the UK Biobank study data and used the imputed SNP data from chromosome 22 as supplied in the data release. We simulated phenotypes under contrasting generative models:

- We chose markers of high LD with other SNPs to be the causal variants and we assigned effects proportional to the LD score of those markers and their minor allele frequency. To do this, we first grouped the SNPs using the clumping procedure in Plink (see Code Availability) based on 1 - MAF, selecting the highest frequency variants and removing any variants with LD < 0.01, to obtain 4988 independent SNPs. For these 4988 SNPs we calculated the LD score of the markers. We then assigned effect sizes to these selected SNPs, drawing them from a single normal distribution with variance ~ LD_score^1^MAF^−1^. We multiplied these effect sizes by the simulated marker values scaled to zero mean and unit variance to create the genetic values with variance 0.5, and then added an environmental component simulated from a normal distribution with mean zero and variance 1 minus the variance of the genetic values to give a phenotype with zero mean and unit variance.
- We then took the same 4988 SNPs but assigned effect sizes to the markers at random from a normal distribution with zero mean and variance 0.5*/*4988. We multiplied these effect sizes by the simulated marker values scaled to zero mean and unit variance to create the genetic values with variance 0.5, and then added an environmental component simulated from a normal distribution with mean zero and variance 1 minus the variance of the genetic values to give a phenotype with zero mean and unit variance.
- We then sampled randomly 4988 evenly spaced markers as causal variants, but assigned effect sizes proportional to the LD score and minor allele frequency of the markers as described above. We multiplied these effect sizes by the simulated marker values scaled to zero mean and unit variance to create the genetic values with variance 0.6, and then added an environmental component simulated from a normal distribution with mean zero and variance 1 minus the variance of the genetic values to give a phenotype with zero mean and unit variance.
- Finally, we then sampled randomly 4988 evenly spaced markers as causal variants and randomly assigned the effect sizes from a normal distribution with zero mean and variance 0.5*/*4988. We multiplied these effect sizes by the simulated marker values scaled to zero mean and unit variance to create the genetic values with variance ~ 0.5, and then added an environmental component simulated from a normal distribution with mean zero and variance 1 minus the variance of the genetic values to give a phenotype with zero mean and unit variance.

This replicates our main simulation study, but creates a situation where there is an association at every LD block on chromosome 22 and thus the results seen in the main simulation study should be magnified here. We analysed 50 simulation replicates of each of the four scenarios with BayesR, BayesRR-RC with 20 MAF-LD groups (deciles of MAF, each split into two groups based on median LD score within each MAF decile), and a MLMAi model implemented in software GCTA. For the Bayesian methods we ran three chains with different starting values for each of the 200 simulation replicates for 3000 iterations, removing the first 1500 iterations as burn-in and taking the posterior mean across the three chains. In Figure S1a we plot the distribution of the posterior mean for BayesR and BayesRR-RC, and the MLMA point estimates, of the proportion of variance attributable to the SNP markers minus the true simulated value obtained across the 50 simulation replicates for each of the four scenarios, showing inflation of the MLMA estimates when selecting high LD variants, and inflation of the BayesR estimates with high LD and random effect size estimates. In contrast, estimates obtained from BayesRR-RC were unbiased across all scenarios. By simulating an effect size MAF relationship LD_score^1^MAF^−1^, we are assigning the smallest absolute effect size values to the most common SNPs, which appears to limit the inflation of the estimates for BayesR, when selecting high LD SNPs as causal variants (Figure S1a). We then examined the effect size estimates obtained from these three approaches across the MAF spectrum under the second scenario of high LD causal variant selection, but random effect size allocation, to show using z-scores calculated as the estimated effects minus the simulated effects, divided by the SD of the simulated effects. We find overestimation of common variant effect sizes under BayesR, and dramatic inflation of effect size estimates under MLMA showing poor recovery of the underlying effect size distribution (Figure S1b). Grouping effects by MAF and LD in a BayesRR-RC model resolved this overestimation issue (Figure S1b) as seen in our original large-scale simulation study.

We then explore the ability of the model to recover a different set of annotation-specific effect sizes using the same set of 50,000 randomly selected UK Biobank individuals and imputed genotype data for chromosome 22 grouped by chromatin state annotations (15-state ChromHMM model) from the epigenome of primary mononuclear cells from peripheral blood (E062) of the Epigenome Roadmap Project [56]. We simulated the genetic architecture as follows :

- We first mapped SNPs to active and inactive chromatin states from the mnemonic bed files for E062 (see Code availability). 37,187 SNPs mapped to active chromatin states including transcription start site (TSS) and their flanking regions, genic and other enhancers, untranslated transcribed regions (UTR) and actively transcribed regions and zinc finger genes states. 27,224 SNPs mapped to inactive states including heterochromatin, bivalent/poised TSS and their flanking regions, bivalent enhancers and repressed polycomb states. The remaining 47,018 SNPs were grouped and labelled as Other SNPs (Figure 1d).
- To simulate enrichment in both chromatin states, we randomly sampled 2000 SNPs as causal variants from variants mapped to active chromatin states and another 2000 SNPs from variants mapped to inactive chromatin states. We then assigned effect sizes to these 4000 selected SNPs, drawing them from a normal distribution with zero mean and variance 0.35*/*2000 for active states and 0.15*/*2000 for inactive states.
- We multiplied annotation-specific effect sizes by the simulated marker values scaled to zero mean and unit variance to create the annotation-specific genetic values with variance 0.35 for active states, 0.15 for inactive states and 0 for other SNPs. We finally added an environmental component simulated from a normal distribution with mean zero and variance 1 minus 0.5 (the sum of the genetic values) to give a phenotype with zero mean and unit variance.

We analyzed 20 simulation replicates with our BayesRR-RC software specfiying annotations (active states, inactive states and other SNPs) with 2 LD groups based on median LD score within each annotation. We compared our software to BoltREML [20] and RHEmc [21] both multi-variance component methods that also use individual-level data but provide single heritability estimates per genetic component. For BayesRR-RC we ran three chains with different starting values for each of the 20 simulations replicates for 3000 iterations, removing the first 1000 iterations as burn-in and taking the posterior mean across the three chains. We then performed the same analysis but randomly assigning SNPs to each annotation resulting in mis-specification of the underlying genetic architecture. In Figure S1d, we plot the estimated sum of the squared regression coefficients that is evenly split across the three annotations when misspecifying the underlying genetic architecture (labelled : Misspecification of groups) and shows enrichment when we properly assign SNPs to annotation (labelled : Multiple group enrichment). We find that BayesRR-RC performs as boltREML and RHEmc, with RHEmc estimates showing higher variability, supporting our main simulation results.

We also further examined the ability of BayesRR-RC to recover effect sizes compared to BayesR by comparing 10 simulations of 5 chains with different starting values where each simulation has two groups in high LD with an interdigitated structure where one in two SNPs is assigned to group 1 (Figure S3). We then simulated phenotypes as previously described, randomly selecting 1000 causal variants in group 1 only, using 20,000 randomly selected UK Biobank individuals and imputed genotype data for chromosome 2 (with MAF > 0.05). In Figure S3, we compare the proportion of markers entering the model in group 1 and group 2 at different posterior inclusion probability thresholds. Annotation-specific estimates for BayesR are calculated post-analysis for each group. We also compare the correlation of estimated genetic values with the truth when using BayesRR and BayesR. For this, we conducted estimation of marker effects in an independent data set to compare prediction accuracy. We simulated 10 new phenotypes and computed the genetic value 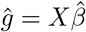 where *X* is the genotype matrix and 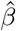 is a vector of estimated marker effects for each individual. Figure S3 shows BayesRR-RC has improved model performance over BayesR to recover effect sizes and infer underlying genetic architectures.

Finally, we also extend our validation the use of PPWV in simulation study, first simulating 500 replicate data sets of 10,000 SNP markers for 5,000 individuals for each of two scenarios. In the first scenario, 1000 SNPs are randomly selected to be causal variants and all 10,000 SNP markers are LD independent. In the second, the 1000 causal variants are each in LD with four other variants with LD = 0.95, with the remaining 5000 variants having zero effect size and LD = 0. For each scenario, we simulate effect sizes as an equally spaced sequence from an effect size of −0.04 SD, to 0.04 SD giving genetic variance of 0.55, and we simulate residual variance from a normal distribution with zero mean and variance 0.45, to give a phenotype with zero mean and unit variance. For the first scenario, we calculate the posterior inclusion probability of each causal SNP. For the second scenario, we calculate the PPWV for each 5-SNP group. Across the 500 replicates of each scenario, we take the mean PPWV and mean PIP for each of the 1000 different effect sizes and compare these in Figure S4a. Additionally, we grouped SNPs in 50kb regions and selected the number of regions that explain at least 0.1%, 0.01% and 0.001% of the variance attributed to all SNP markers in 0.8% to 100% of the iterations using the simulated data described above for the multiple group enrichment scenario for chromosome 22 in the UK Biobank. We then calculated the false discovery rate (FDR), defined as the proportion of 50kb regions identified that do not contain a causal variant, at PPWV thresholds ranging from 0.8% to 100%. We compare these in Figure S4b where as we lower the PPWV variance threshold, the number of false discoveries in the model increases but remains at ≤ 5% when the PPWV is ≥ 95%. This further demonstrates that our proposed PPWV approach is an appropriate metric of summarising the posterior distribution to identify associated genomic regions, irrespective of the genomic region used.

### The influence of population structure and relatedness

We then investigated the importance of controlling for multicollinearity for the control of population genetic and data structure effects. In principle, a MLMA approach will control for bias with correlated markers (either local or long-range LD) fitted as random when testing for the effects of a focal SNP. For two markers, **X**_1_and**X**_2_ in LD correlation 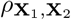, with *β*_2_ = 0 we can express the MLMA fixed effect solution as a partial regression coefficient of the phenotype regressed onto the focal SNP after adjusting for **X**_2_ estimated as 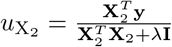. Following our derivation above for a shrinkage estimator of a partial regression coefficient the effect size of **X**_**1**_ is estimated as 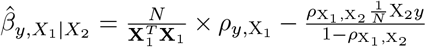 and in this two-SNP example the bias is accounted for in the term 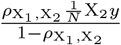 when the fixed effect is estimated. Multicollinearity acts to increase the *σ*_*G*_ term of *λ*, reducing the denominator 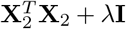 in the estimation of 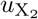, and increasing the variance of the estimates of common markers in high LD, those with the highest average *F*_*ST*_.

We conducted a simulation study using real genomic data from chromosome 22 where 10,000 individuals were selected from 2 UK Biobank assessment centres (Glasgow and Croydon). First, causal variants were allocated to 5000 high-LD SNPs with effect sizes simulated from a normal distribution with variance proportional to the *F*_*ST*_ among the two populations at each SNP. Second, we selected the same high-LD SNPs as the causal variants, but simulated effect sizes to have correlation 0.5 with the allele frequency differences of the SNPs among the two populations, and thus not only is the effect size proportional to the *F*_*ST*_, but there is also directional differentiation (trait increasing loci tend to be those with higher allele frequency in Croydon, trait decreasing alleles have lower frequency in Croydon). For each of these two scenarios, we simulated 50 replicate phenotypes where the phenotypic variance attributable to the causal SNPs is 0.5, there is a phenotypic difference where Croydon individuals have a phenotype that is 0.5 SD higher than Glasgow individuals (contributing variance 0.05), and residual variance was simulated from a normal with variance 0.45, to give a phenotype with mean of zero and variance of 1. The data were then analysed using a mixed-linear model association (MLMAi implemented in GCTA) and a grouped Bayesian dirac spike and slab models (BayesR implemented in hydra). In the analysis, we either adjusted the phenotype by the first 20 PCs of the genetic data used in the simulation study, or we did not adjust the phenotype for the PCs, to examine the effects of this common methods of population stratification control. In a two-population scenario the leading eigenvector encapsulates the allele frequency differentiation between the populations and thus the expectation is that this should adjust for these differences when estimating the marker associations. The results are presented in Figure S5a, where we find that an MLMA approach overestimates the variance attributable to the SNPs under all scenarios, both with and without adjustment for PCs. BayesRR returns accurate estimates when the variance of the marker effects is proportional to *F*_*ST*_ and underestimates the variance when there is a directional associations, with this underestimation being less severe with PC adjustment.

Finally, we also assess the influence of relatedness on the estimates obtained from a bayesR model using real genomic data from chromosome 21 and 22 (226,662 SNP markers) and 10,000 families randomly selected from the UK Biobank (26,034 individuals). Here, we selected 2000 LD blocks with a single causal SNP per block at random, with an LD block defined as a group of SNP markers with absolute LD of at least 0.01. We assigned effect sizes to these 2000 selected SNPs, drawing them from a normal distribution with zero mean and variance 0.5/2000. We then multiplied effect sizes by the simulated marker values scaled to zero mean and unit variance to create the genetic values with variance 0.5. In addition to the genetic component, we added a common environment component to simulate effects coming from shared familial environment. We simulated four scenarios where each family was assigned the same common environment effect drawn from a normal distribution with variance 0 (no common environment), 0.1, 0.2 and 0.3. Finally, we added an environmental component simulated from a normal distribution with mean zero and variance 1 minus the genetic variance and minus the common environment variance. We analysed 20 replicates of each of the four scenarios with BayesRR-RC with 6 MAF-LD groups (terciles of MAF, each split into two groups based on median LD score within each MAF tercile). In Figure S5, we summarise 800 samples of the posterior distribution from 5000 iterations with a thin of 5 and removing the first 1000 iterations as burn-in. We find that the variance attributable to the SNPs, the regression coefficients and the posterior probability of window variance (PPWV) remain unchanged with relatedness and with increasing family effects.

### Testing algorithm performance and parallelism

We then explored the influence of increasing parallelism in our algorithm. We used the simulated data described above for the randomly sampled 50,000 UK Biobank individuals with imputed genotype data for chromosome 22, where we sampled randomly 4988 evenly spaced markers as causal variants and randomly assigned the effect sizes from a normal distribution with zero mean and variance 0.6*/*4988 (the fourth scenario). For each of the 50 simulation replicates, we compared the three chains obtained by running the BayesRR-RC model (with 20 MAF-LD groups) in serial, with a single MPI task and synchronisation rate of 1 (residual updating after sampling each SNP), to three chains obtained by increasing the number of MPI tasks to 4 and then to 8, with synchronisation rates of 10 and 20 sampling steps before residual updating. For each simulation, we ran three chains of our BayesRR-RC model with different starting values for 3000 iterations. Like with all MCMC chains of regression models, convergence and sampling properties will be problem specific and dependent upon the LD of the markers, LD among the causal variants, the phenotypic variation attributable to the SNP markers across the MAF and LD spectrum, the study sample size, the degree of data parallelism per total marker number, and the synchronisation rate. Thus, the aim here is to simply show a series of diagnostic tests that can be utilized to explore the properties of the posterior to highlight how the different metrics can be used to identify convergence issues. We use the distribution, across simulations, of the proportion of effective samples obtained for the hyperparameter estimate of the proportion of phenotypic variance attributable to the markers of each group. This shows that for all ranges of parallelism, we achieve more effective samples for low MAF and low LD variants. As high MAF SNPs are interchangeable in the model to a large degree, their entry and exit from the model is correlated across iterations, and thus this is entirely expected and is actually a consequence of the model mixing. With high synchronisation rates, where many marker updates occur before residual updating by message passing a reduction in effective sample sizes occurs. We also use the distribution of the Gelman-Rubin test statistic for the three chains, a general metric to monitor convergence that compares within- and among-chain variance, as the number of iterations increases. Finally, a Geweke statistic value can be used to test the equality of the means of the first and last part of the Markov chains. We present the results of this simulation in Figure S6 also including the distribution of z-scores of the posterior distribution of the phenotypic variance attributable to the markers for each MAF-LD group from the simulated values, which show stability of the estimates obtained with increasing data parallelism (tasks), but that a very high synchronisation rate with high parallelism can lead to poor convergence rates, meaning that the chains would have to be run for longer (Figure S6).

### UK Biobank data

We restricted our discovery analysis of the UK Biobank to a sample of European-ancestry individuals. To infer ancestry, we used both self-reported ethnic background (UK Biobank data code 21000-0) selecting coding 1 and genetic ethnicity (UK Biobank data code 22006-0) selecting coding 1. We also took the 488,377 genotyped participants and projected them onto the first two genotypic principal components (PC) calculated from 2,504 individuals of the 1,000 Genomes project with known ancestries. Using the obtained PC loadings, we then assigned each participant to the closest population in the 1000 Genomes data: European, African, East-Asian, South-Asian or Admixed, selecting individuals with PC1 projection < absolute value 4 and PC 2 projection < absolute value 3. This gave a sample size of 456,426 individuals.

To facilitate contrasting the genetic basis of different phenotypes, we then removed closely related individuals as identified in the UK Biobank data release. While the BayesRR model can accommodate relatedness similar to mixed linear models, we wished to simply compare phenotypes at markers that enter the model due to LD with underlying causal variants. Relatedness leads to the addition of markers within the model to capture the phenotypic covariance of closely related individuals, and this will vary across traits in accordance with the genetic and environmental covariance for each phenotype. For these unrelated individuals, we used the imputed autosomal genotype data of the UK Biobank provided as part of the data release. We used the genotype probabilities to hard-call the genotypes for variants with an imputation quality score above 0.3. The hard-call-threshold was 0.1, setting the genotypes with probability ≤ 0.9 as missing. From the good quality markers (with missingness less than 5% and p-value for Hardy-Weinberg test larger than 10-6, as determined in the set of unrelated Europeans) were selected those with minor allele frequency (MAF) > 0.0002 and rs identifier, in the set of European-ancestry participants, providing a data set 9,144,511 SNPs, short indels and large structural variants. From this we took the overlap with the Estonian Genome centre data to give a final set of 8,430,446 markers. From the UK Biobank European data set, samples were excluded if in the UKB quality control procedures they (i) were identified as extreme heterozygosity or missing genotype outliers; (ii) had a genetically inferred gender that did not match the self-reported gender; were identified to have putative sex chromosome aneuploidy; (iv) were excluded from kinship inference. Information on individuals who had withdrawn their consent for their data to be used was also removed. These filters resulted in a data set with 382,466 individuals.

We then selected the recorded measures of BMI (UK Biobank variable identifier 21001-0.0) and height (variable identifier 50-0.0) collected during initial assessment visit (year 2006-2010). BMI and height phenotypes 6 standard deviations (SD) away from the mean were not included in the analyses. For Type 2 Diabetes (T2D) in UKB we selected as cases very broadly as individuals who have main or secondary diagnosis (UKB fields 41202-0.0 − 41202-0.379 and 41204-0.0 − 41204-0.434) of “non-insulin-dependent diabetes mellitus” (ICD 10 code E11) or self-reported non-cancer illness (UKB field 20002-0.0 − 20002-2.28) “type 2 diabetes” (code 1223). From respondents self-reporting just “diabetes” (code 1220), we selected as cases those who did not self-report “type 1 diabetes” (code 1222) and had no Type 1 Diabetes (ICD code E10) diagnosis. Individuals with self-reported “diabetes” and “type 1 diabetes”/E10 were also left out from controls. We also defined coronary artery disease (CAD) cases broadly as participants with one of the following primary or secondary diagnoses or cause of death: ICD 10 codes I20 to I28; self-reported angina (code 1074) or self-reported heart attack/myocardial infarction (code 1075). Participants with self-reported “heart/cardiac problem” (code 1066) were not included as cases but also excluded from controls. This gave a sample size for each trait of 25,773 T2D cases and 359,730 T2D controls, 39,766 CAD cases and 344,054 CAD controls, 382,402 measures of height and 381,899 measures of BMI.

All phenotypes were adjusted for age of attending assessment centre (UKB code 21003-0.0, factor with levels for each age), year of birth (UKB field 34-0.0, factor with levels for each year), UK Biobank recruitment centre (UKB field 54-0.0, factor with levels for each centre), Genotype batch (UKB field 22000, factor with levels for each batch) and final 20 leading principal components of 1.2 million LD clumped markers from the 8,430,446 markers included in the analysis, calculated using flashPCA (see Code Availability). The residuals were then converted to z-scores with 0 mean and variance of 1. Similarly as for relatedness, population stratification is also accounted for within the BayesRR model through the addition of a background of marker effects entering the model, however we also wished to account for this in the standard manner by adjusting for the leading 20 PCs of the SNP data to get as close as possible to the inclusion of markers in the model that reflect LD with the causal variants. We note that as with any association model, while we take steps to adjust for known spatial (UKB centre), batch, and ancestry effects, and that the effects of each SNP is estimated jointly (and thus conditionally on the effects of all the other SNPs) environmentally induced covariance between SNP markers and a phenotype is still possible.

We partition SNP markers into 7 location annotations using the knownGene table from the UCSC browser data (see Code Availability), preferentially assigned SNPs to coding (exonic) regions first, then in the remaining SNPs we preferentially assigned them to intronic regions, then to 1kb upstream regions, then to 1-10kb regions, then to 10-500kb regions, then to 500-1Mb regions. Remaining SNPs were grouped in a category labelled “others” and also included in the model so that variance is partitioned relative to these also. Thus, we assigned SNPs to their closest upstream region, for example if a SNP is 1kb upstream of gene X, but also 10-500kb upstream of gene Y and 5kb downstream for gene Z, then it was assigned to be a 1kb region SNP. This means that SNPs 10-500kb and 500kb-1Mb upstream are distal from any known nearby genes. We further partition upstream regions to experimentally validated promoters, transcription factor binding sites (tfbs) and enhancers (enh) using the HACER, snp2tfbs databases (see Code Availability). All SNP markers assigned to 1kb regions map to promoters; 1-10kb SNPs, 10-500kb SNPs, 500kb-1Mb SNPs are split into enh, tfbs and others (un-mapped SNPs) extending the model to 13 annotation groups. Within each of these annotations, we have three minor allele frequency groups (MAF<0.01, 0.01>MAF>0.05, and MAF>0.05), and then each MAF group is further split into 2 based on median LD score. This gives 78 non-overlapping groups for which our BayesRR-RC model jointly estimates the phenotypic variation attributable to, and the SNP marker effects within, each group. For each of the 78 groups, SNPs were modelled using five mixture groups with variance equal to the phenotypic variance attributable to the group multiplied by constants (mixture 0 = 0, mixture 1 = 0.0001, 2 = 0.001, 3 = 0.01, 4 = 0.1). We conducted a series of convergence diagnostic analyses of the posterior distributions to ensure we obtained estimates from a converged set of four Gibbs chains, each run for 6,000 iterations with a thin of 5 for each trait (Figure S8, S9, S10, S11).

We calculate PPWV for LD blocks of the genome, by first calculating the minor allele frequency of each SNP (*p*) and using 1− *p* in a Plink clumping procedure to select LD independent (*correlation*^2^ ≤ 0.1) blocks of SNPs. We then repeat the estimation of the PPWV of 50kb regions across the genome, then map SNPs to the coding region of genes, and to the closest gene +/− 50kb from the SNP position labelling them as located in a coding region, an intron, 1kb upstream of a gene using our functional annotations. Remaining SNPs are labelled as located in a cis-region (up to +/− 50kb from a gene). Finally, we mapped SNPs with greater than 50% posterior inclusion probability (PIP) across all 4 chains labelling them using our 7 location annotations (Figure S14). We report SNPs with PIP > 95% and their corresponding p-value from reported GWAS summary statistics (fastGWA, see Code Availability) with ‘body mass index’ entry for BMI, ‘standing height’ for HT, ‘angina / heart attack’ for CAD and ‘diabetes’ for T2D (Supplementary Table S6).

We then compared our BayesRR-RC estimates for height, BMI, T2D and CAD to RHEmc [21] which also relies on individual level data. We ran RHEmc with 10 independent random vectors and 100 jackknife blocks on the 382,466 individuals and 8,430,446 SNP markers assigned to our 78 non-overlapping groups. SNP heritability estimates, enrichment and standard errors per genetic component are reported in Supplementary Table 8. We intended to include SNP heritability estimates from Bolt-REML [20] in the method comparison but the run time and memory usage exceeded 7 days and 900 GB which is the limiting run-time and memory for our HPC system. Among the summary statistic methods, we ran sLDSC [11] and SumHer [6]. To do so, we created summary statistics containing marginal associations for each of the 8,430,446 markers using plink2 [55] for height, BMI, T2D and CAD. For sLDSC, we computed univariate LD scores and annotation-specific LD scores for the 78 non-overlapping groups using a window size of 10,000 kb and a subset of 20,000 individuals randomly selected from the full data set. We then partitioned heritability with our annotations and no restriction on MAF. SNP heritability estimates, proportions of heritability, enrichment and standard errors per genetic component are reported in Supplementary Table 9. For SumHer, we computed LDAK weightings and created tagging files separately by chromosomes using the full data set (M=8,430,446 and N=382,466) as reference and a window size of 1,000 kb. Because SNPs included in groups *others* and *rare 1Mb tfbs* are not present in all chromosomes, tagging files are constructed using 70 non-overlapping annotations only. The remaining SNPs are modeled together in an extra partition. Finally, we merged the tagging files and regressed the summary statistics onto this file assuming the LDAK model. SNP heritability estimates, proportions of heritability, enrichment and standard errors per genetic component are reported in Supplementary Table 10. The proportion of genetic variance estimated genome-wide with RHE-mc, sLDSC and SumHer are shown in Table2. We also report the proportion of genetic variance attributed to SNPs located in exons, introns, 1kb, 1-10kb and 10-500kb regions and the proportion of common SNPs located in exons, introns and 10-500kb regions computed from the single heritability estimates observed (Table2).

In addition to plink2 [55] summary statistics, we also applied Bolt-LMM [8] and Regenie [9] to height, BMI, T2D and CAD. In the first step, we pruned SNPs using plink [57] with a pairwise r2 threshold of 0.5 and a window size of 1,000 kb, resulting in a subset of 1,362,013 SNPs markers. We restricted the random effects in the mixed model for bolt-LMM and the ridge regression predictors for Regenie to this subset of pruned SNPs. In the second step, all 8,430,446 SNPs from the full genotype data were then tested for association in both methods. Following recommendations, we used the provided hg19 genetic map file and 1000 Genomes LD scores reference for Bolt-LMM and performed the default mixed linear model association test. For Regenie, the 1,362,013 SNP markers are split in blocks of 1,000 consecutive SNP markers and ridge regression predictors are computed for a range of 5 shrinkage parameters within each block. For the association testing, we split the 8,430,446 SNP markers in blocks of 400 consecutive SNP markers and set the Firth correction p-value threshold to 0.01. We then applied an approximate and joint association analysis called GCTA-COJO [58] to the summary statistics obtained with Bolt-LMM, Regenie and plink2. We ran GCTA-COJO using a subset of 20,000 individuals randomly selected from the 382,466 individuals as reference with a window size of 10,000 kb and a r2 cutoff value of 0.5 for the LD among the SNPs in the data. Finally, we set a p-value threshold to 5*e* − 8 to report significant SNPs associated with height, BMI, CAD an T2D in Table 3.

### Estonian Genome Centre data

For the Estonian Genome Centre Data, 32,594 individuals were genotyped on Illumina Global Screening (GSA) arrays and we imputed the data set to an Estonian reference, created from the whole genome sequence data of 2,244 participants [59]. From 11,130,313 markers with imputation quality score > 0.3, we selected SNPs that overlapped with the UK Biobank, resulting in a set of 8,433,421 markers.

We selected height and BMI measures from the Estonian Genome Centre data, in 32,594 individuals genotyped on GSA array and converted them to sex-specific z-scores after applying the same outlier removal procedure as in UKB and adjusting for the age at agreement. Prevalent cases of CAD and T2D in the Estonian Biobank cohort were first identified on the basis of the baseline data collected at recruitment, where the information on prevalent diseases was either retrieved from medical records or self-reported by the participant. The cohort was subsequently linked to the Estonian Health Insurance database that provided additional information on prevalent cases (diagnoses confirmed before the date of recruitment) as well as on incident cases during the follow-up.

As the UK Biobank marker effects are estimated from traits that were standardized to a z-score prior to analysis, all effect sizes obtained are on the SD scale. Thus when we create a genomic predictor, for say coding SNPs, by multiplying SNPs mapped to coding regions genotyped in Estonia to the effect sizes obtained in the UK Biobank for each iteration, to obtain a genetic predictor for each iteration, providing a posterior predictive distribution that is also on the SD scale. For each trait, we created 2000 genomic predictors for each individual in the Estonian Biobank, at each of the 13 annotation groups, by selecting effect size estimates obtained every tenth iteration from the last 3000 iterations of each of the four Gibbs chains and combining them together in a single posterior.

We calculated prediction accuracy as the proportion of phenotypic variation explained by the genomic predictor, and area under the receiver operator curve (AUC) for T2D and CAD using each individual’s mean genetic predictor. For each of the 13 annotation groups, we calculated the partial correlation of the genetic predictor of each of the 2000 iterations and the phenotype, and then used this to estimate the independent proportional contribution of each group to the total prediction accuracy, providing a metric of replication for our UK Biobank enrichment results.

For height and BMI, we determined the probability that each Estonian individual’s predictor accurately reflected their phenotypic value. To do this, we calculated the proportion of posterior samples with abs(*ĝ* −*y*) of less than 1 for each individual, which gives a measure of the degree to which each posterior predictive distribution overlaps with the phenotype within +/− 1 SD.

For T2D and CAD, we extended the PCF metric, typically defined as the proportion of cases with larger estimated risk the then top *p*^*th*^ percentile of the distribution of genetic risk in the general population. We calculated the proportion of posterior samples for each individual with values in the top 25% of the distribution of genomic predictors for each trait. Thus for each individual, we calculate the probability that the posterior predictive distribution is in the top 25% of the distribution of genetic risk in the general population.

As a comparison, we also estimated a boltLMM prediction model using MegaPRS [24] as recommended by the authors and as shown to have the best prediction performance out of the MegaPRS approaches in our simulation study. We clumped SNPs with r2 threshold of 0.5 resulting in 1,508,624 SNP markers to be included in the analysis and randomly selected 20,000 individuals to compute the LDAK weights. We then computed the tagging file using the same data set as reference and the 64 BLD-LDAK annotations. Here, weights are models as an extra annotation and we save the heritability matrix. We then regress the plink2 [55] summary statistics for height, BMI, CAD and T2D onto the tagging file, saving the per-predictor heritabilities. We then created 4 reference panels with the same 1,508,624 SNP markers but randomly selecting different 5,000 related individuals from the UK Biobank and we used these to: (i) calculate predictor-predictor correlations with a window size of 3,000 kb to estimate the LD structure; (ii) compute pseudo summaries from the plink2 summary statistics including ambiguous alleles, which creates pseudo training and test summary statistics to be used in the construction of the prediction model; (iii) estimate effect sizes specifying a bolt-LMM model for height, BMI, CAD and T2D, using the predictor-predictor correlations, the per-predictor heritabilities, the plink2 summary statistics and training pseudo summary statistics, whilst including ambiguous allele and specifying a 1000 kb window; (iv) test prior distributions to determine the most accurate model and obtain the best effect sizes. These steps resulted in 1,397,514 predictors for height, 1,471,586 for BMI, 1,397,514 for CAD and 1,389,364 for T2D and we ensured at no point was the Estonian genome centre data used, nor was the any overlapping individuals in the UK Biobank subsets used to train the models and the data used to generate the summary statistics. Finally, we then calculated genomic predictors for each individual in the Estonian Biobank using the best effect sizes. We report the squared correlations between the genomic predictor and phenotypes.

## Data availability

This project uses UK Biobank data under project 35520. The Estonian Genome Centre data are available upon request from the cohort authors with appropriate research agreements. Summaries of all posterior distributions obtained are provided in Supplementary data sets. Full posterior distributions of the SNP marker effects sizes for each trait are deposited on Dryad https://datadryad.org/

## Data Availability

This project uses UK Biobank data under project 35520. The Estonian Biobank data are available upon request from the cohort authors with appropriate research agreements.

## Code availability

Our BayesRR-RC model is implemented within the software Hydra, with full open source code available at: https://github.com/medical-genomics-group/hydra.

UCSC Table Browser https://genome.ucsc.edu/cgi-bin/hgTables

flashPCA https://github.com/gabraham/flashpca

Plink1.90 https://www.cog-genomics.org/plink2/

GCTA https://cnsgenomics.com/content/software

HACER database http://bioinfo.vanderbilt.edu/AE/HACER/

snp2tfbs database https://ccg.epfl.ch//snp2tfbs/

fastGWA database http://fastgwa.info/ukbimp/phenotypes/

Computing environment https://www.epfl.ch/research/facilities/scitas/hardware/helvetios/

## Author contributions

MRR conceived and designed the study. MP, DTB, and AK contributed to the study design. MP and MRR conducted the experiments and analyses with input from DTB, AK, SEO, AH, JS, PMV, RM and LR. MRR, DTB, SEO, and LR derived the equations and the algorithm. EJO and DTB developed the software, with contributions from MRR, MP, SEO, AK, and GM. MRR, MP, and DTB wrote the paper. RM provided study oversight and contributed data to the analysis. All authors approved the final manuscript prior to submission.

## Author competing interests

The authors declare no competing interests.

## Acknowledgements

This project was funded by an SNSF Eccellenza Grant to MRR (PCEGP3-181181), and by core funding from the Institute of Science and Technology Austria. We would like to thank the participants of the cohort studies, and the Ecole Polytechnique Federal Lausanne (EPFL) SCITAS for their excellent compute resources, their generosity with their time and the kindness of their support. PMV acknowledges funding from the Australian National Health and Medical Research Council (1113400) and the Australian Research Council (FL180100072). LR acknowledges funding from the Kjell Märta Beijer Foundation (Stockholm, Sweden). We also would like to acknowledge Simone Rubinacci, Oliver Delanau, Alexander Terenin, Eleonora Porcu, and Mike Goddard for their useful comments and suggestions.

## Supplementary Online Material

**Figure S1.**
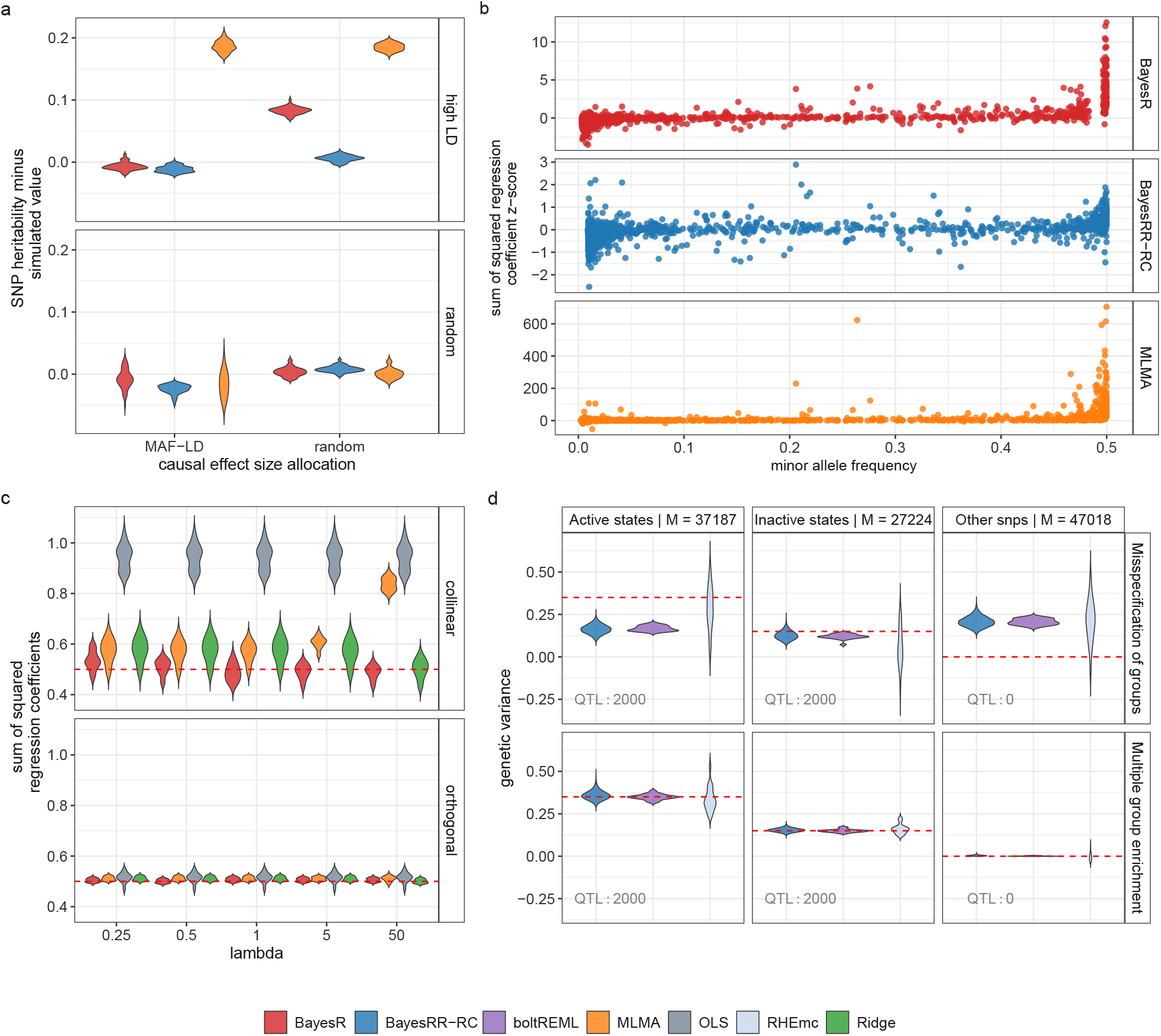
Theory and simulation study for genetic penalized regression models under multicollinearity. (a) Smaller-scale simulation study then that presented in the main text using real genomic data from chromosome 22 where 50 replicate phenotypes were generated by either allocating 5000 LD-independent causal variants to high LD SNPs (y-axis panel: high LD), or randomly allocating 5000 SNPs as causal variants (y-axis panel: random), and then either randomly allocating effect sizes to those SNPs (x-axis: random), or allocating effect sizes proportional to their LD and MAF (x-axis: MAF-LD, see Methods). In this simulation every LD block of chromosome 22 contributes to the trait variance. SNP heritability estimation error is plotted as the difference of the estimate and the true simulated value across the 50 replicates. (b) We then investigated this further for the scenario where causal variants are allocated to high-LD SNPs. While the 5000 causal variants are LD-independent, they are each correlated with a large number of SNPs of simulated effect size 0. For each causal variant, we took all the markers in LD ≥0.05 and summed the squared estimated regression coefficients of these markers. The true simulated value is simply the square of the effect size allocated to the causal variant, and we subtracted this from the sum of the squared regression coefficients divided by the SD of the simulated genetic effects, to give a z-score for each causal variant and this is plotted on the y-axis for MLMA, BayesR, and BayesRR-RC. (c) Our theory outlines how this overestimation is the result of the effect of multicollinearity (see Methods) and an example is shown here, where 50 pairs of SNP markers with LD = 0.9 were simulated for each of 50 simulation replicates, where only one marker of each pair has an effect (0,0.1 SD), giving the sum of the squared regression coefficients as 0.5 for each simulation (dotted red line). lambda is the shrinkage parameter, the ratio of the error variance and the variance attributable to the SNP markers, used for MLMA, ridge regression (Ridge) and the BayesR model to estimate the effects. (d) Simulation of a genetic architecture (dotted red line) using real annotations from the Epigenome Roadmap Project [56] (active states, inactive states, other snps). We compared BayesRR-RC to other recent approaches providing annotation-specific variance component estimates in individual-level data when SNPs are randomly assigned to an annotation (labelled: misspecification of groups), or when specifying enrichment using prior knowledge (labelled: multiple group enrichment)

**Figure S2.**
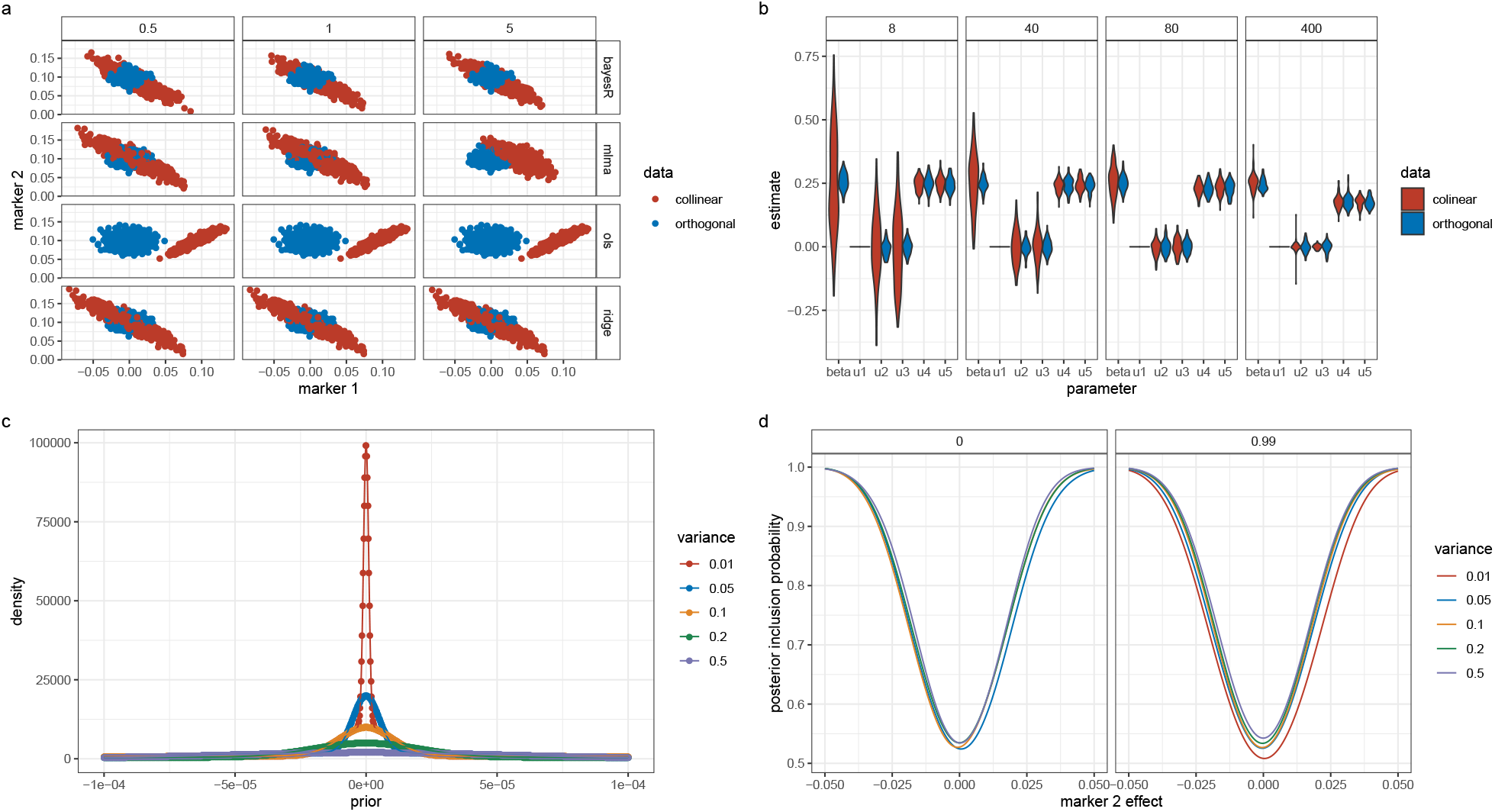
Theory and simulation study of SNP marker model parameters. (a) accompanies Eq. (23) and shows the distribution of the point estimates of the effect sizes of two correlated markers of effect size (0,0.1) under orthogonality (LD = 0) and collinearity (LD = 0.99) across 2500 replicates (50 independent genomic regions for 5,000 individuals within each of 50 replicates) for a range of different models: a dirac spike and slab mixture of regressions model (bayesR), a mixed linear association model (MLMA), single-marker ordinary least squares (OLS), and ridge regression (Ridge). Panels give the lambda shrinkage parameter of the model, the error variance divided by the phenotypic variance attributable to the SNP markers, showing that as lambda decreases the variation of the estimates increases under multicollinearity. (b) accompanies Eq.(26) and shows the marker estimates obtained from Henderson’s mixed model equations for a MLMA with the focal marker as fixed (beta) and random (u1), with four other markers in the model. Markers were either uncorrelated (orthogonal, LD=0) or the focal marker was correlated with the first two out of the four other markers (collinear, LD=0.99). Panels give the lambda shrinkage parameter, showing that as lambda decreases the variation of the estimates increases under multicollinearity. (c) shows the prior density of the BayesR model for different hyperparameter values of the phenotypic variance attributable to genetic effects (variance), showing that as the variance attributable to the markers decreases, the prior has higher mass around zero. Thus, with a grouped mixture of regressions model (BayesRR-RC), each hyperparameter estimate will be smaller and thus there will be higher prior density around zero. This then has consequences for marker inclusion in the BayesRR model. Higher prior mass around zero makes little difference for the inclusion of uncorrelated markers, but it results in reduced posterior inclusion probability for correlated markers as shown in (d). For (d), we calculated the inclusion probability (PIP) of two markers with LD = 0 and LD = 0.99, as the variance attributable to the SNP markers, and thus the prior distribution, changes assuming a background inclusion probability of 0.1, a sample size of 5000, and an effect size of 0.01 SD for marker 1 (see Methods). (d) shows that the PIP of the second marker is reduced across a range of possible effect size values (the average of 1000 replicated simulations for 1000 marker 2 effect values for each line) as the hyperparameter estimate decreases, and thus the smaller hyperparameter estimates in a BayesRR model means that correlated markers are less likely to enter the model, controlling better for the effects of multicollinearity.

**Figure S3.**
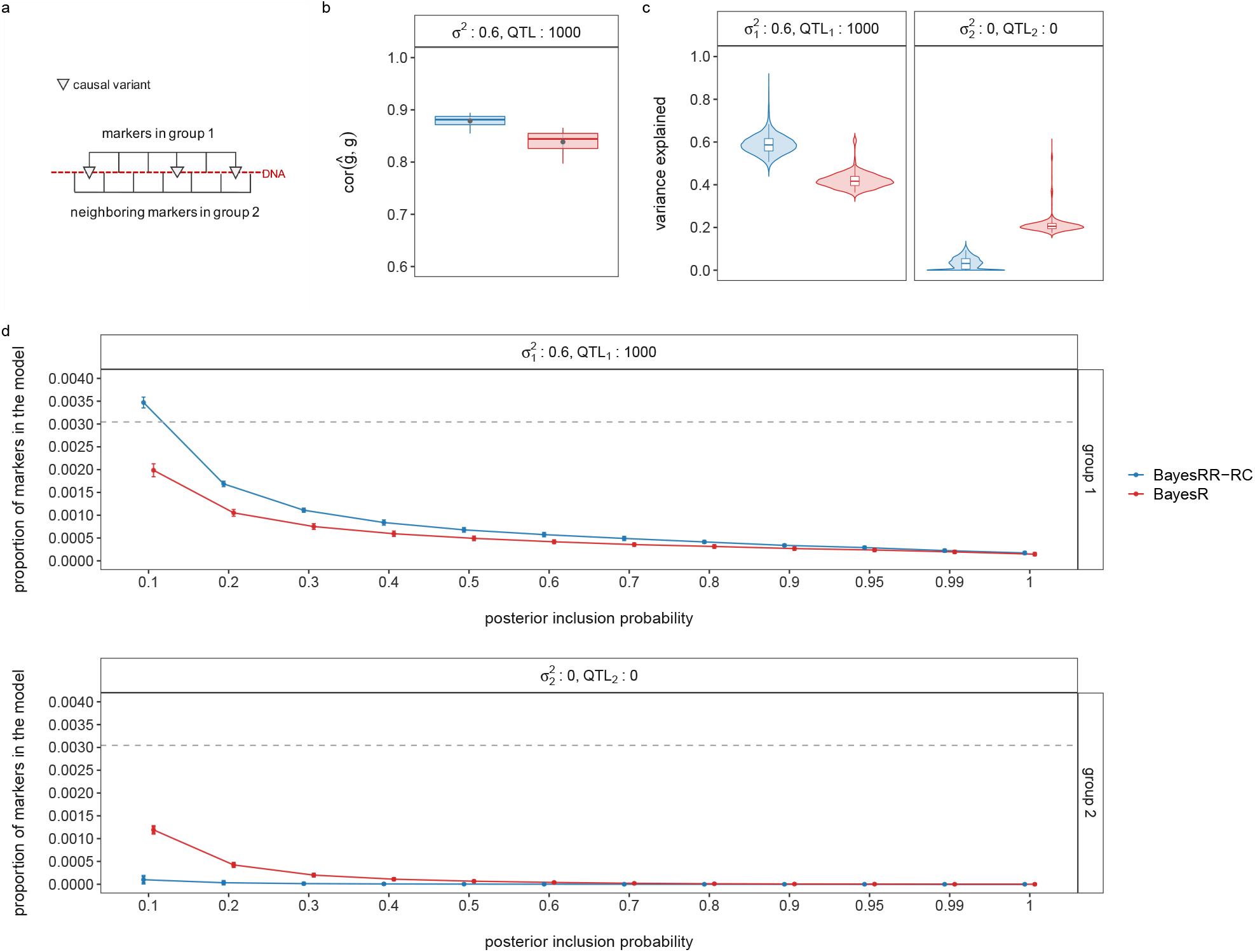
Classification power of BayesRR-RC. Grouping effects in a BayesRR-RC model improves the power of BayesR to estimate effect sizes and infer the genetic architecture of common complex traits and diseases. This setting compares 10 simulations of 5 chains with different starting values (chain length : 2500, burn-in : 500, thin : 5) executed using BayesRR-RC. (a) Each simulation has two groups in high LD with an interdigitated structure where one in two SNPs is assigned to group 1 and all genetic variance is assigned to group 1 with 1000 QTL. Annotation-specific estimates for BayesR are calculated post-analysis for each group. (b) Estimation of markers effects in an independent data set. BayesRR-RC improves on correlation between predicted and simulated genetic values. This increase in prediction implies that adding functional information to BayesR better fits the data and improves prediction accuracy. (c) Genetic variance and (d) proportion of markers entering the model at posterior inclusion probability (pip) thresholds summarized across 10 simulations for group 1 and group 2. The proportion of markers included in the model is closer to the truth (dotted grey line) when using BayesRR-RC compared to BayesR. Effects are thus more likely attributed to the correct group using our approach, which also explains why we estimate more accurately the group genetic variance compared to the baseline. Simulation setting: *N* = 20, 000 unrelated European individuals from the UK Biobank, *M* = 328, 385 markers (chromosome 2). Dots in box plots show the mean of the correlation between predicted and simulated genetic values.

**Figure S4.**
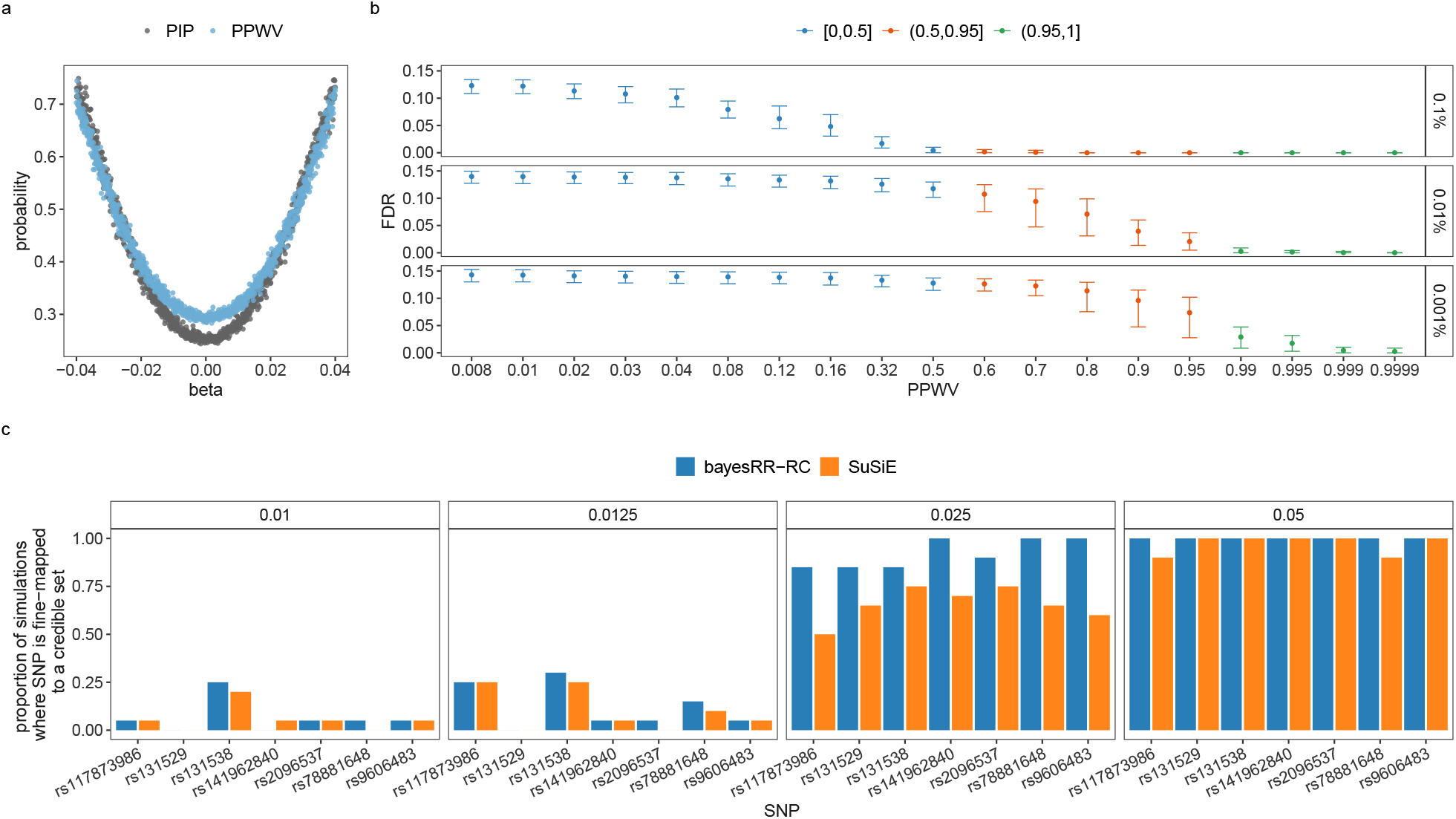
Posterior inclusion probability (PIP) and posterior probability of window variance (PPWV). (a) We validate the use of PPWV in simulation study, first simulating 500 replicate data sets of 10,000 SNP markers for 5,000 individuals for each of two scenarios. In the first scenario, 1000 SNPs are randomly selected to be causal variants and all 10,000 SNP markers are LD independent. In the second, the 1000 causal variants are each in LD with four other variants with LD = 0.95, with the remaining 5000 variants having zero effect size and LD = 0. For each scenario, we simulate effect sizes as an equally spaced sequence from an effect size of −0.04 SD, to 0.04 SD giving genetic variance of 0.55, and we simulate residual variance from a normal distribution with zero mean and variance 0.45, to give a phenotype with zero mean and unit variance. For the first scenario, we calculate the posterior inclusion probability of each causal SNP. For the second scenario, we calculate the PPWV for each 5-SNP group. Across the 500 replicates, we take the mean PIP for each SNP of the 1000 different effect sizes for the first scenario and the mean PPWV of each of the 1000 5-SNP windows for the second scenario, and these are the points on the figure. (b) Shows mean and 95% credible interval of the false discovery rate (FDR), defined as the proportion of regions identified that do not contain a causal variant, at PPWV thresholds ranging from 0.8% to 100%. Here, we grouped SNPs in 50kb regions and selected the number of regions that explain at least 0.1%, 0.01% and 0.001% of the variance attributed to all SNP markers in 0.8% to 100% of the iterations using simulated data for chromosome 22 in the UK Biobank (see Methods). We compare the FDR at these different PPWV thresholds and as we lower the PPWV variance, the number of false discoveries in the model increases, but remains at ≤ 5% at PPWV ≥95%. (c) A comparison of BayesRR-RC and SuSiE where we assigned effect sizes of either 0.05, 0.025, 0.0125, or 0.01 on the SD scale to seven SNPs. For BayesRR-RC, we calculate the PPWV of the LD blocks containing the seven focal SNPs, and then prune these blocks based on the LD among the markers in the block to identify a credible set with LD *R*^2^ ≥0.9. We then count the proportion of times across 20 simulation replicates that each causal variant was contained with one of the credible sets. For SuSiE, we calculate the proportion of times that the credible sets identified contained one of the seven causal variants.

**Figure S5.**
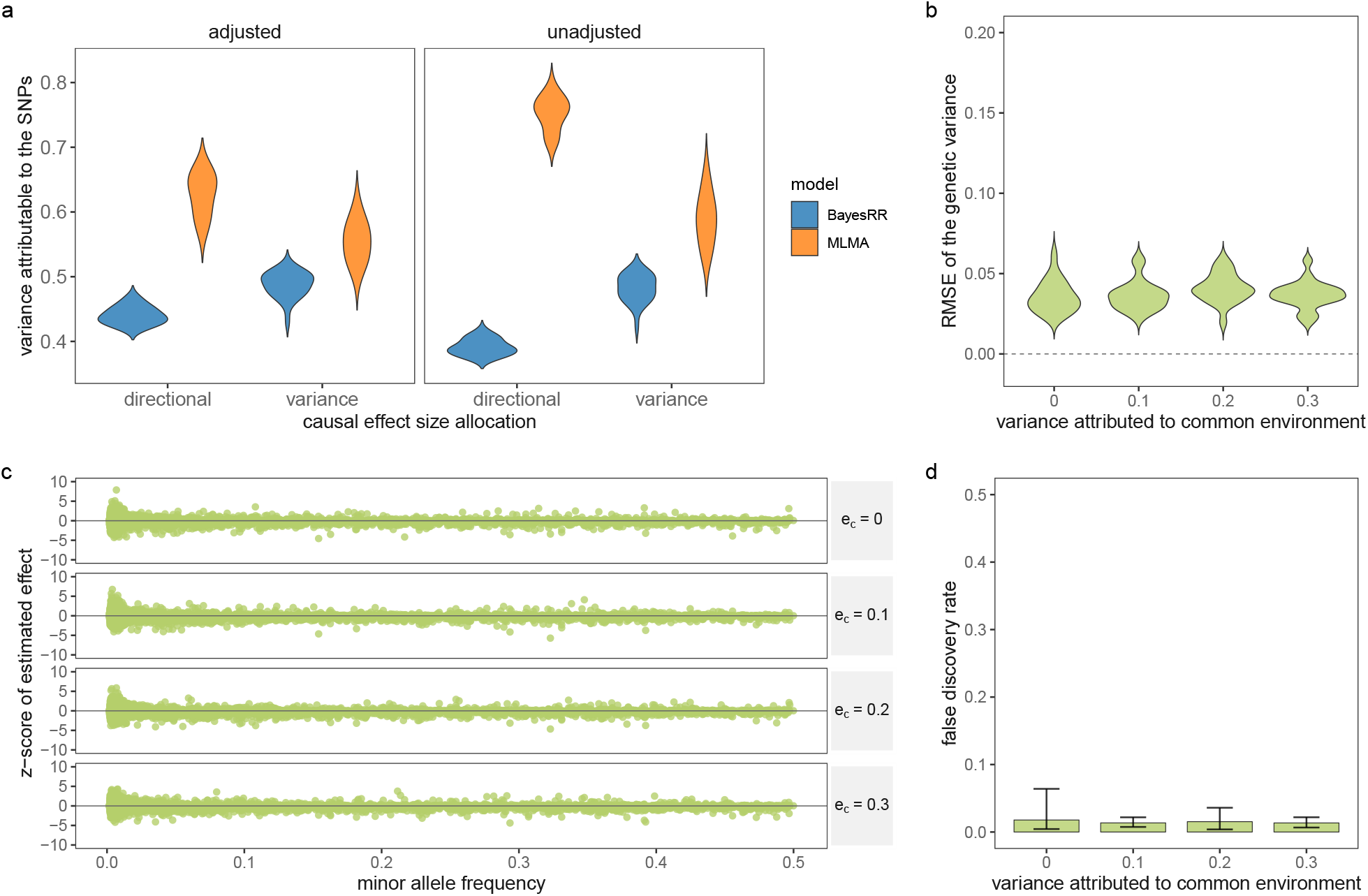
Exploring effects of population stratification and relatedness among samples. (a) Simulation study using real genomic data from chromosome 22 where 10,000 individuals were selected from 2 UK Biobank assessment centres (Glasgow and Croydon). First, causal variants were allocated to 5000 high-LD SNPs with effect sizes simulated from a normal distribution with variance proportional to the *F*_*ST*_ among the two populations at each SNP (labelled ‘variance’, see Methods). Second, we selected the same high-LD SNPs as the causal variants, but simulated effect sizes to have correlation 0.5 with the allele frequency differences of the SNPs among the two populations, and thus not only is the effect size proportional to the *F*_*ST*_, but there is also directional differentiation (trait increasing loci tend to be those with higher allele frequency in Croydon, trait decreasing alleles have lower frequency in Croydon). For each of these two scenarios, we simulated 50 replicate phenotypes where the phenotypic variance attributable to the causal SNPs is 0.5, there is a phenotypic difference where Croydon individuals have a phenotype that is on average 0.5 SD higher than Glasgow individuals (contributing variance 0.05), and residual variance was simulated from a normal with variance 0.45, to give a phenotype with mean of zero and variance of 1. The distribution across simulations of the estimated phenotypic variance attributable to the SNP markers is shown for each of the two causal effect size allocation scenarios when the data was analysed using a mixed-linear model association (MLMA, distribution of the point estimates) and a grouped Bayesian dirac spike and slab models (BayesRR-RC, distribution of the posterior means). In the analysis, we either adjusted the phenotype by the first 20 PCs of the genetic data used in the simulation study (“adjusted”) or we did not adjust the phenotype for the PCs (“unadjusted”). (b), (c) and (d) show bayesRR-RC simulation results using real genomic data from chromosome 21 and 22 and 10,000 families randomly selected from the UK Biobank. We simulated 20 replicates where we selected 2000 LD blocks at random, with an LD block defined as a group of SNP markers with squared LD correlation of at least 0.15. We assigned a causal SNP per LD block and for each replicate, we simulated 4 phenotypes increasing the variance attributed to family effects from 0 (no common environment) to 0.3 (see Methods). (b) Violin-plot of the root mean square error (RMSE) of the SNP-heritability estimates across simulation replicates. For each LD block of each simulation replicate, we summed the squared regression coefficient estimates of all SNPs in the block and took the posterior mean. We then calculated the z-score of the LD block and plotted it against the minor allele frequency of the causal variant of the block. (d) Shows mean and 95% credible intervals of the false discovery rate defined as the posterior probability of window variance (PPWV), of ≥ 95% at 0.001% variance threshold that did not contain a causal variant.

**Figure S6.**
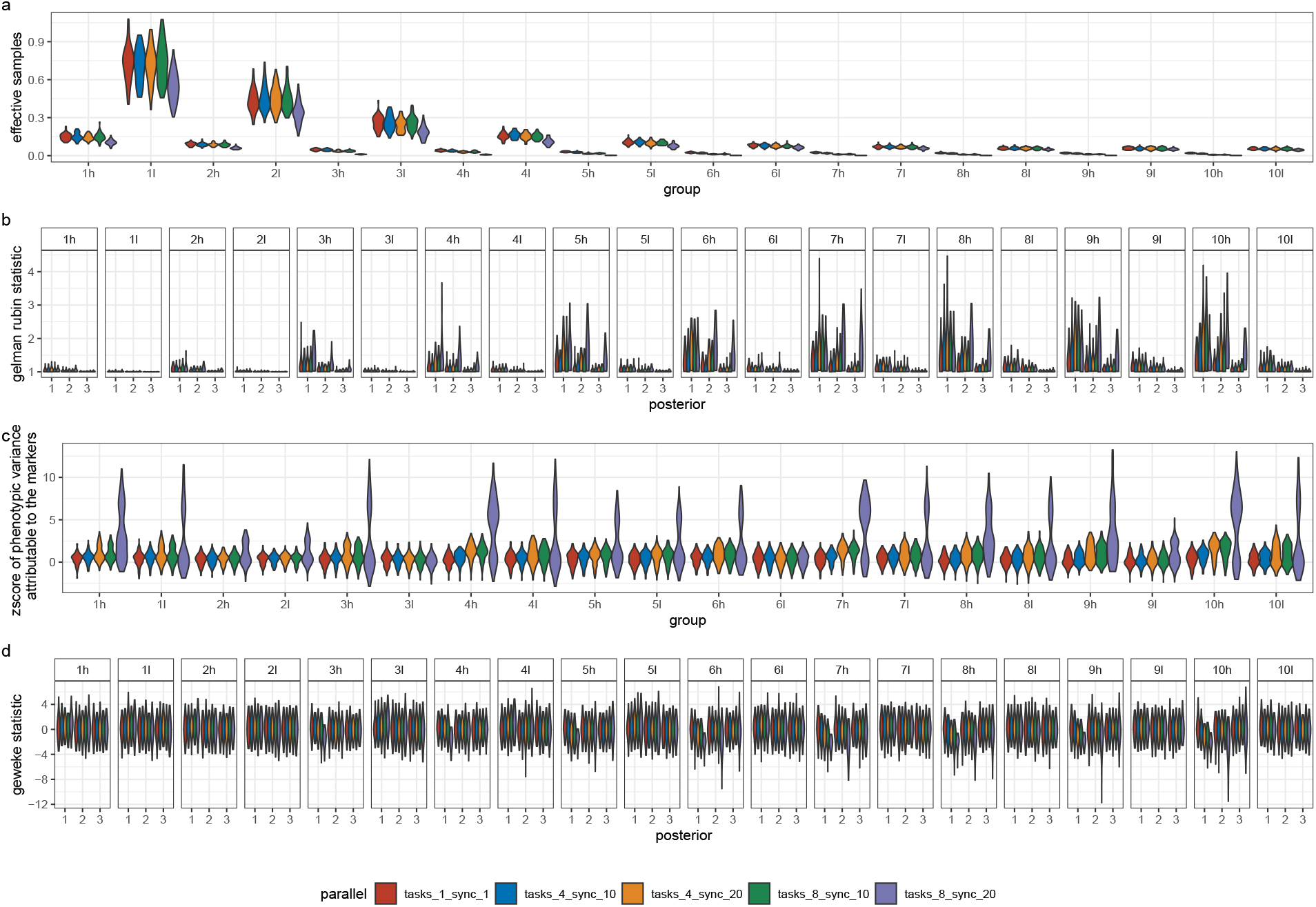
Simulation study of increasing task parallelism and increasing message passing rate for our hybrid-parallel sampling scheme. We aimed to compare (a) the effective samples obtained, (b) the convergence rate of the algorithm, (c) the accuracy of the estimation, and (d) the stability of the estimates obtained as data parallelism increases within a burn-in period of the initial 3000 iterations. For 50,000 randomly selected UK Biobank individuals, and 111,425 imputed SNP markers of chromosome 22, we simulated 50 replicate phenotypes by randomly selecting 4,988 SNPs as causal variants and randomly allocating effect sizes from a normal distribution, with SNP heritability of 0.5. For each simulation, we ran three chains of our BayesRR model with different starting values for 3000 iterations. The SNP marker data was grouped into deciles of the distribution of minor allele frequency (MAF) and within each decile the markers were further grouped these into two groups based on the distribution of linkage disequilibrium (LD), giving twenty groups in total (1l = MAF decile 1, low LD; 1h = MAF decile 1, high LD; …; 10l = MAF decile 10, low LD; 10h = MAF decile 10, high LD). We repeated the three chains, but with increasing data parallelism: (1) in serial where one MPI task is used and the residual is updated after each marker is sampled (tasks_1_sync_1); (2) where the markers were split across four MPI processes with synchronisation occurring by message passing after 10 markers have been updated (task_4_sync_10); (3) where the markers were split across four MPI processes with synchronisation occurring after 20 markers have been updated (task_4_sync_20); (4) with 8 MPI processes and synchronisation of 10 (task_8_sync_10); and (5) with 8 MPI processes and synchronisation of 20 (task_8_sync_20). (a) shows the distribution across simulations of the proportion of effective samples obtained for the hyperparamter estimate of the proportion of phenotypic variance attributable to the markers of each group. For all ranges of parallelism, we achieve more effective samples for low MAF and low LD variants. With high synchronisation rates, where many marker updates occur before residual updating by message passing a reduction in effective sample sizes occurs. (b) gives the distribution of the Gelman-Rubin test statistic for the three chains, a general metric to monitor convergence that compares within- and among-chain variance, as the number of iterations increases. On the x-axis, 1 gives the distribution of the statistic across chains and MAF-LD groups for the first 500 iterations showing divergence of the chains (y-axis value >> 1) across all MAF-LD groups, 2 gives the distribution for the first 1000 iterations, and 3 gives the distribution for the whole chain showing convergence of the chains by the end of this initial 3000 iteration sampling period irrespective of the data parallelism, with the exception of a few groups with infrequent synchronisation and high data parallelism which have yet to converge within this burn-in phase. (c) gives the distribution of z-scores of the posterior distribution of the phenotypic variance attributable to the markers for each MAF-LD group from the simulated values, showing stability of the estimates with increasing data parallelism (tasks), but not with infrequent synchronisation within the 3000 iterations run here. (d) shows the distribution of the Geweke statistic value which is a test of the equality of the means of the first and last part of the Markov chains. On the x-axis, 1 gives the distribution of the statistic calculated using all iterations across all MAF-LD groups, 2 gives the distribution discarding the first 500 iterations, and 3 gives the distribution discarding the first 1000 iterations. (a) - (d) suggest that our hybrid-parallelism sampling scheme achieves the same accuracy and convergence rates as a serial sampling scheme, provided that frequent synchronisation occurs and data parallelism is kept moderate. At high data parallelism and infrequent synchronisation, our theory shows that we are more likely to make a sampling mistake, preventing chains from converging and requiring longer sampling times. Convergence and accuracy of the MCMC Gibbs sampling chain will be problem specific and dependent upon the LD of the markers, LD among the causal variants, the phenotypic variation attributable to the SNP markers across the MAF and LD spectrum, the study sample size, the degree of data parallelism per total marker number, and the synchronisation rate. Therefore, like with all MCMC chains, a series of diagnostic tests can be utilized to explore the properties of the posterior and here we show how different metrics can be used to identify convergence issues.

**Figure S7.**
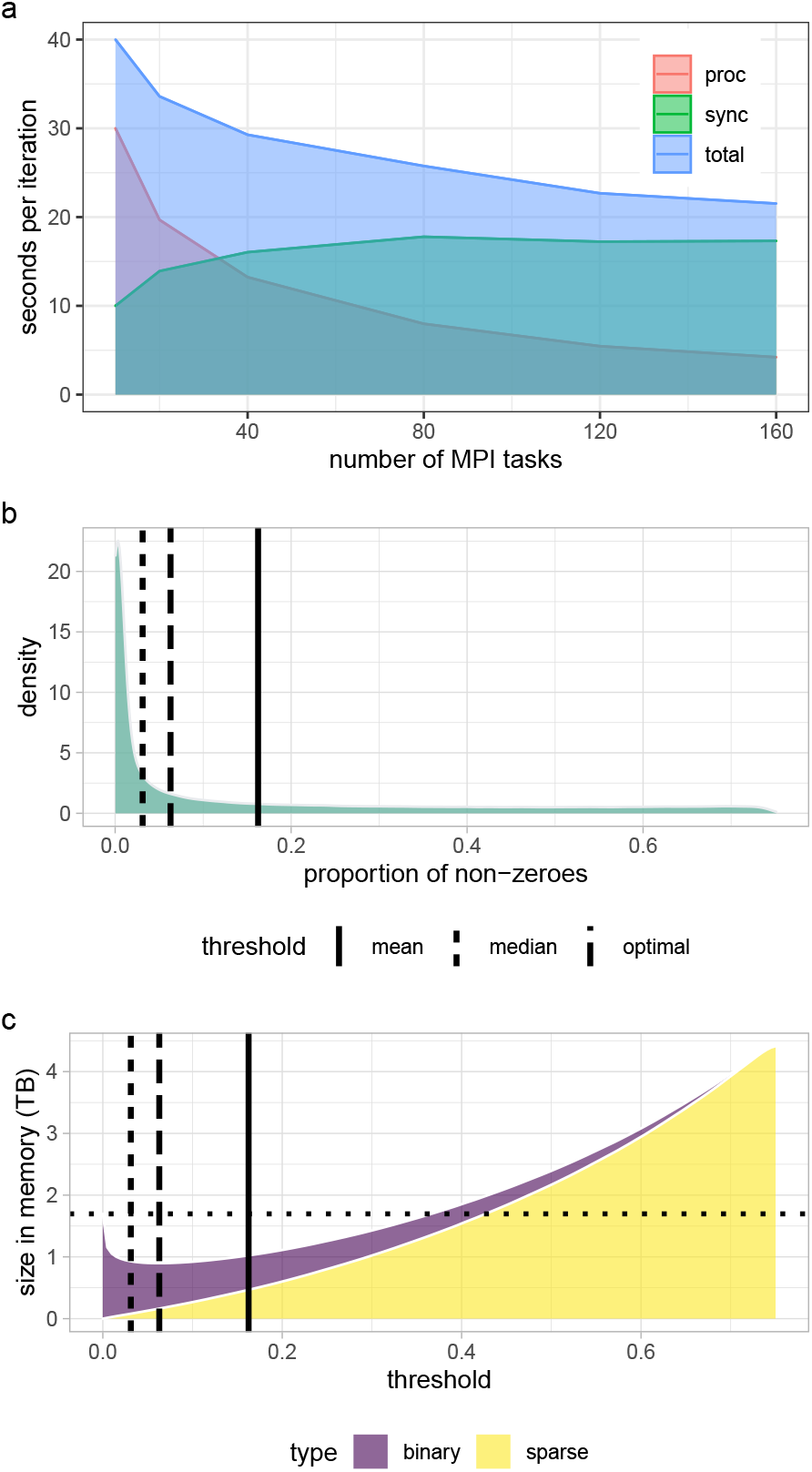
A mixed representation bulk synchronous hybrid-parallel Gibbs sampling scheme for genomic data. (a) The minimum seconds per iteration achieved for 382,466 unrelated individuals from the UK Biobank data genotyped at 8,430,466 markers, with an increasing number of message-passing interface (MPI) tasks used. The total seconds is given in blue and this is subset into (i) the time taken to process the markers and estimate all of the 8,433,421 marker effects and hyper-parameters (proc), and (ii) the time taken to synchronise the estimates as they are being obtained (sync). With increasing data parallelism parameter estimation times drop quickly to less than 5 seconds with 160 MPI tasks, however the time taken to synchronise the estimates increases as the number of tasks increases. The SD was 1 second, with variation in sampling times induced by fluctuations in networking speed that influenced the synchronisation times. Each MPI task was able to used 4 CPUs. (b) the distribution of the proportion non-zeros per column of a genotype matrix for ~4 ×10^5^ individuals and ~1.5× 10^7^ SNPs taken from UKB, with solid line representing the mean of the distribution and dashed line the median. (c) the size in memory in TB of the data as the coding of the SNP markers moves from binary to the sparse indexed format, the optimal threshold is achieved between mean and median of the distribution of non-zeros in the genotype matrix. Above this threshold columns are coded in binary format below in sparse index. Through a combination of a mixed data representation and highly vectorized look-up tables, memory usage is reduced while maintaining fast computational speed.

**Figure S8.**
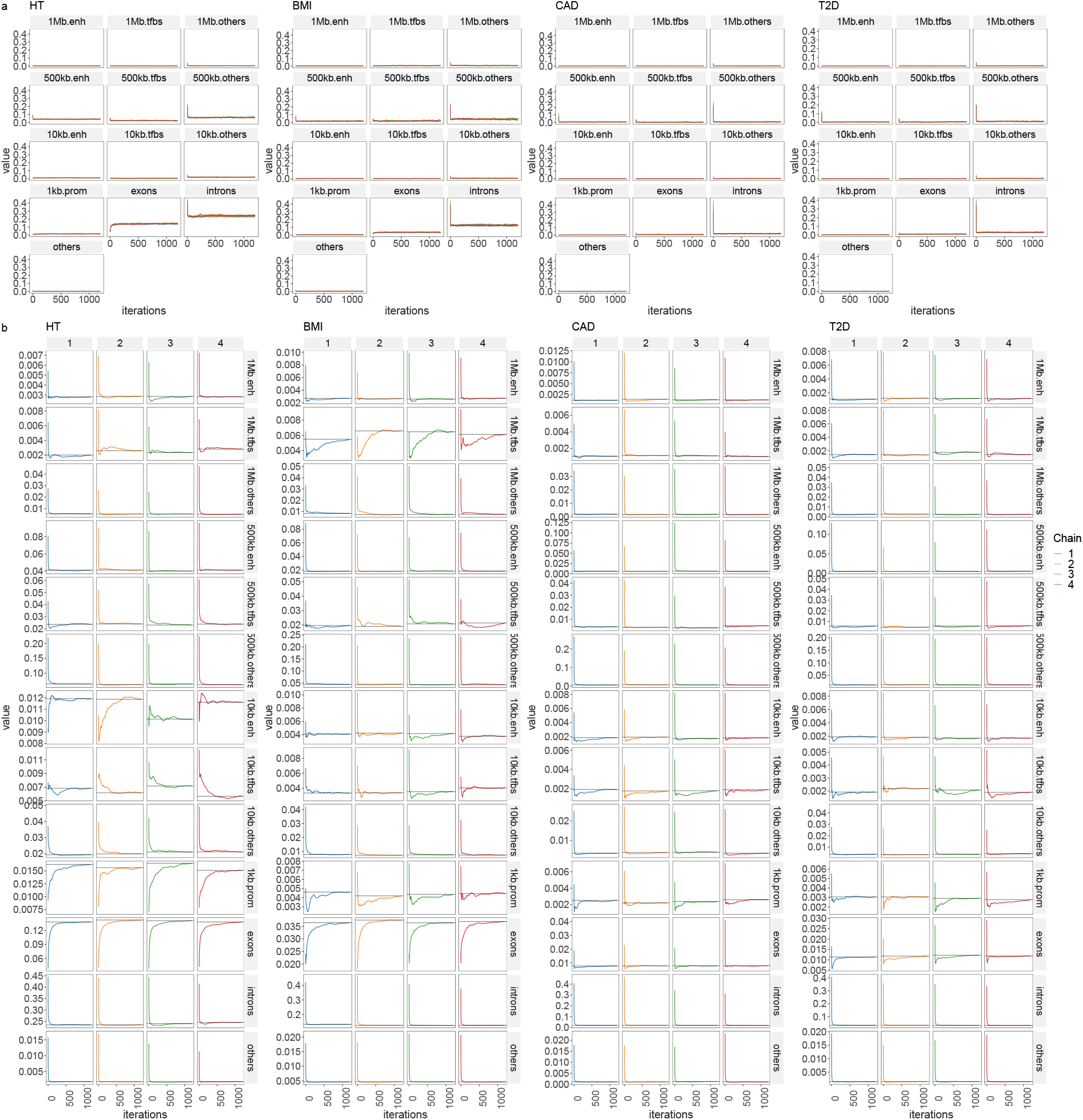
Convergence diagnostics of model chains for UK Biobank analysis. (a) traceplot of the phenotypic variance attributable to SNP markers for each trait across functional annotation of exonic regions, intronic regions, promoters (prom) 1kb upstream of coding regions, enhancers (enh) 1kb to 10kb upstream of coding regions, transcription factor binding sites (tfbs) 1kb to 10kb upstream of coding regions, other snps 1kb to 10kb upstream of coding regions, enh 10kb to 500kb upstream, tfbs 10kb to 500kb upstream, other snps 10kb to 500kb upstream, enh 500kb to 1MB upstream,tfbs 500kb to 1Mb upstream, other snps 500kb to 1Mb upstream and SNP markers elsewhere in the genome (other), with colours representing the different chains. (b) a time series of the running mean of each chain, for each annotation group and each trait showing all chains approach the same mean value for each parameter.

**Figure S9.**
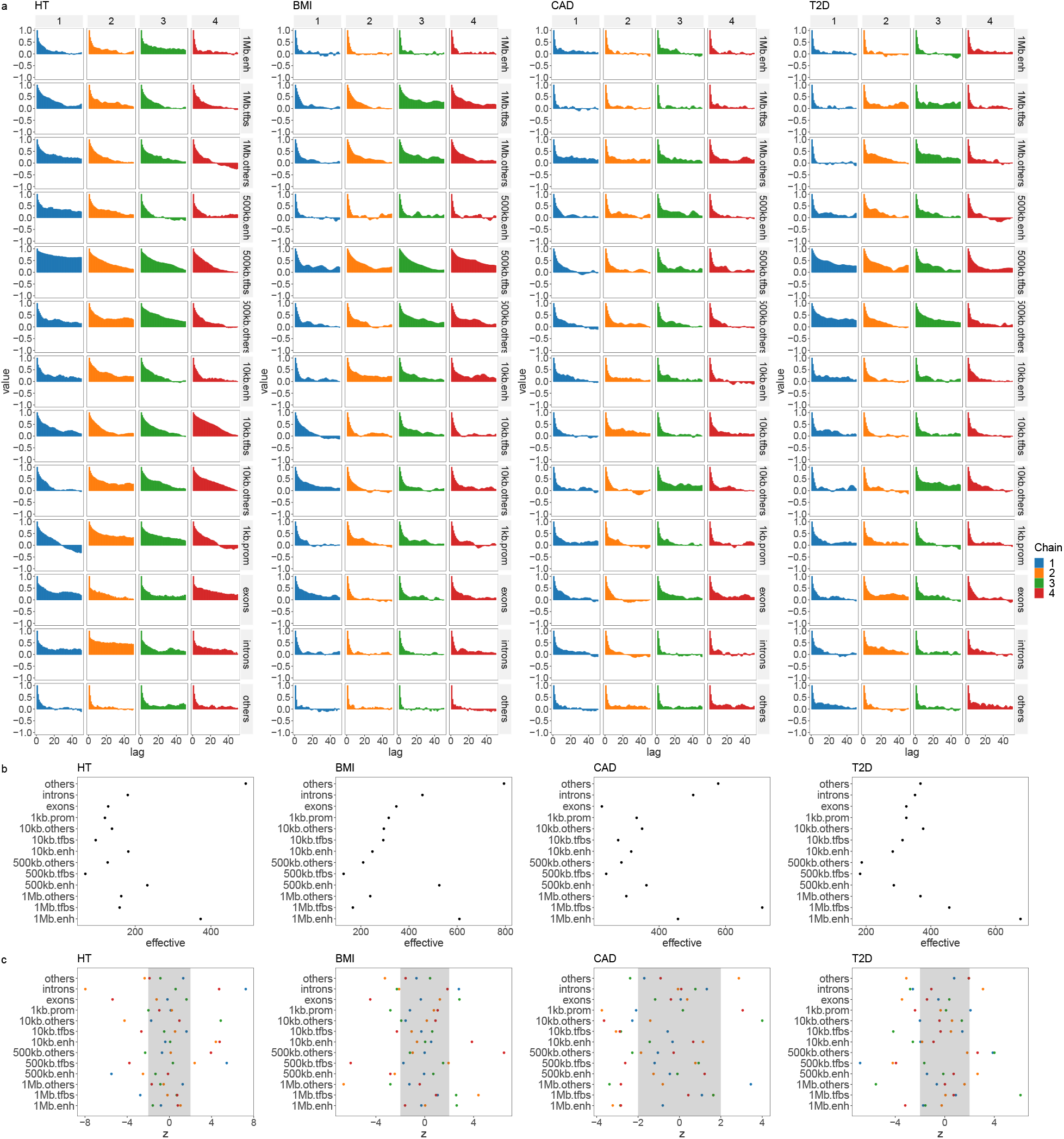
Convergence diagnostics of model chains for UK Biobank analysis. (a) lagged autocorrelation plot of each chain, for each annotation group and each trait and (b) effective number of uncorrelated sampled obtained for each annotation group and each trait. As phenotypic variance is being partitioned it is not expected that posterior estimates obtained are entirely uncorrelated. (c) Geweke z-score statistic comparing the initial part of the chain to the final part, for each annotation group and each trait.

**Figure S10.**
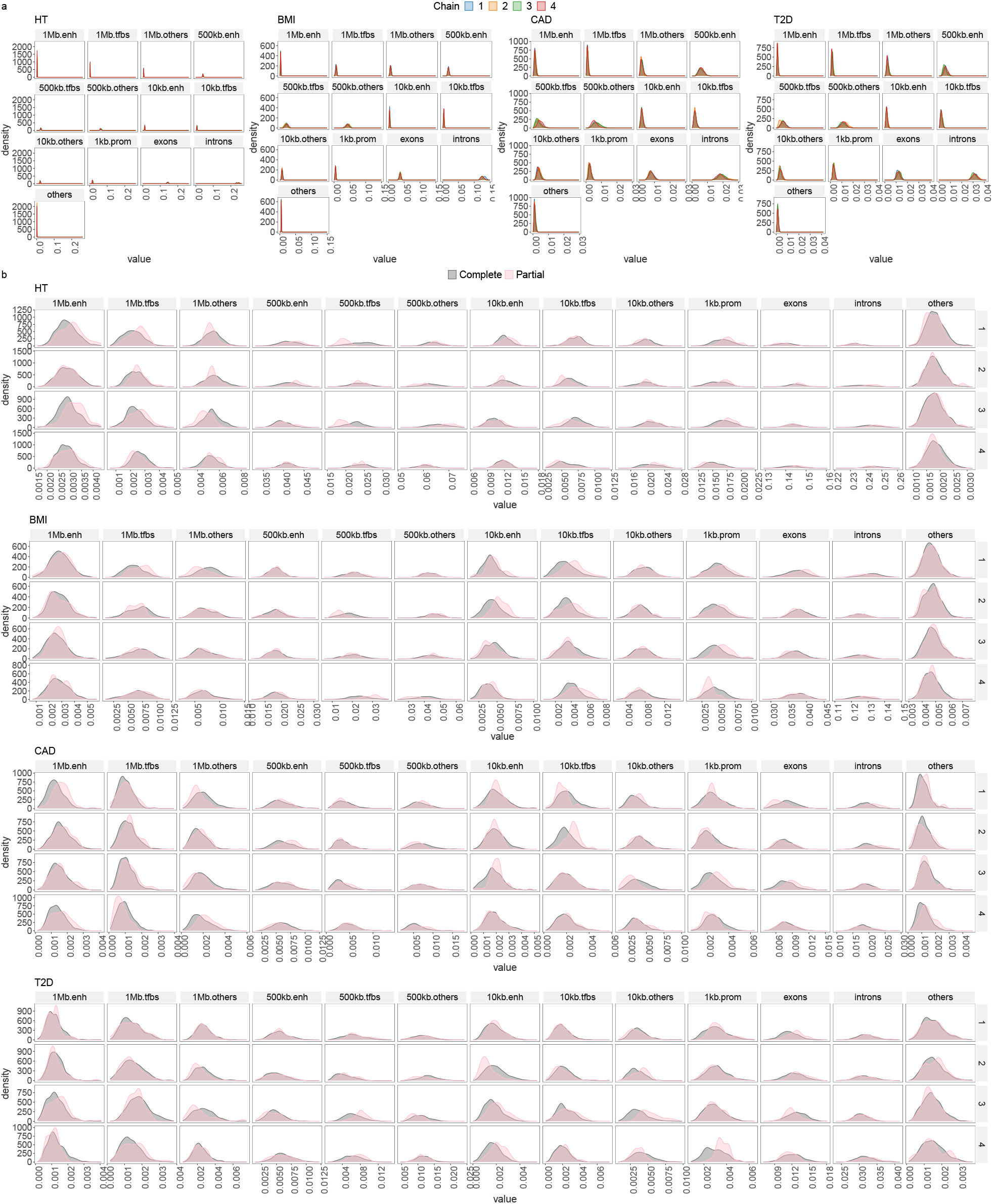
Convergence diagnostics of model chains for UK Biobank analysis. (a) overlapped density plots to compare the target distribution by chain showing each chain has converged in a similar space, for each annotation group and each trait. (b) overlapped density plots comparing the last 10 percent of the chain (green), with the whole chain (pink), showing that the initial and final parts of the chain are sampling the same target distribution for each annotation group and each trait.

**Figure S11.**
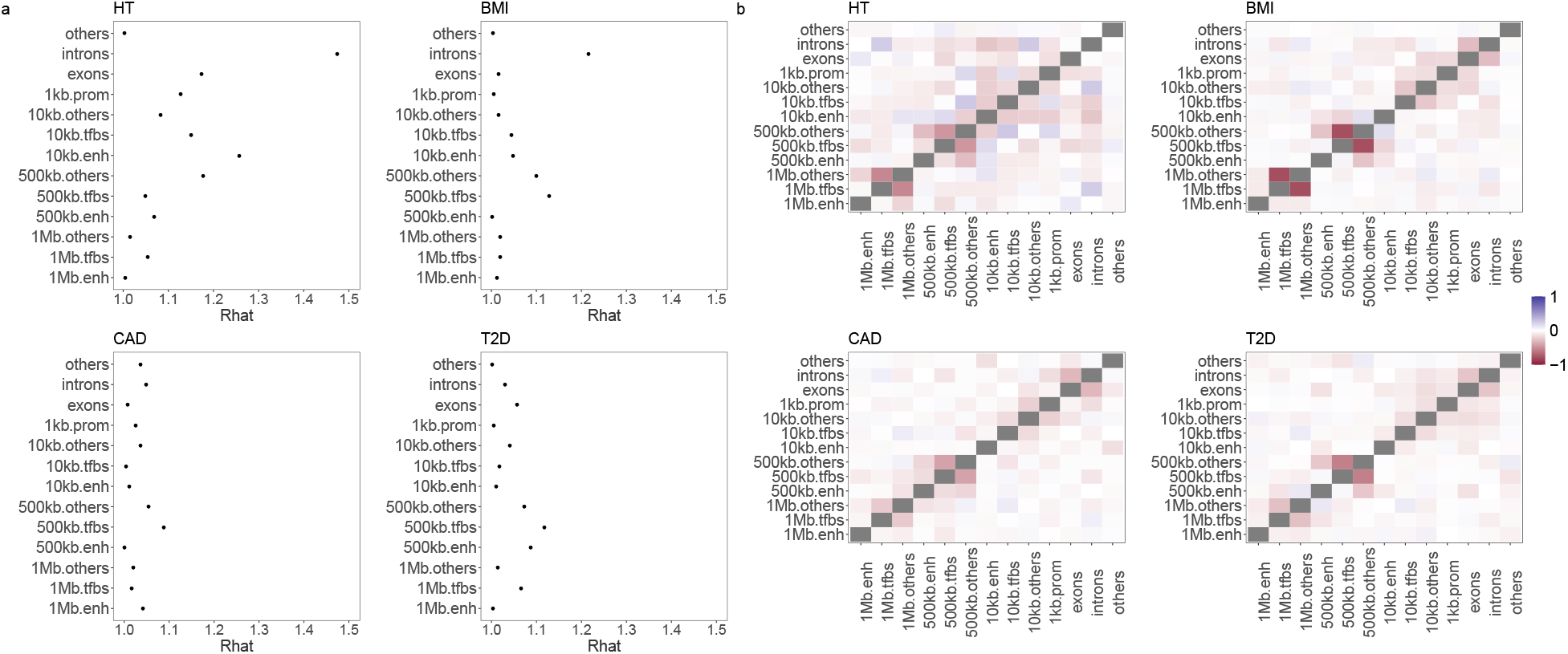
Convergence diagnostics of model chains for UK Biobank analysis. (a) the potential scale reduction factor comparing the among- and within-chain variance for each annotation group and each trait. (b) the cross-correlation between all parameters for each annotation group and each trait.

**Figure S12.**
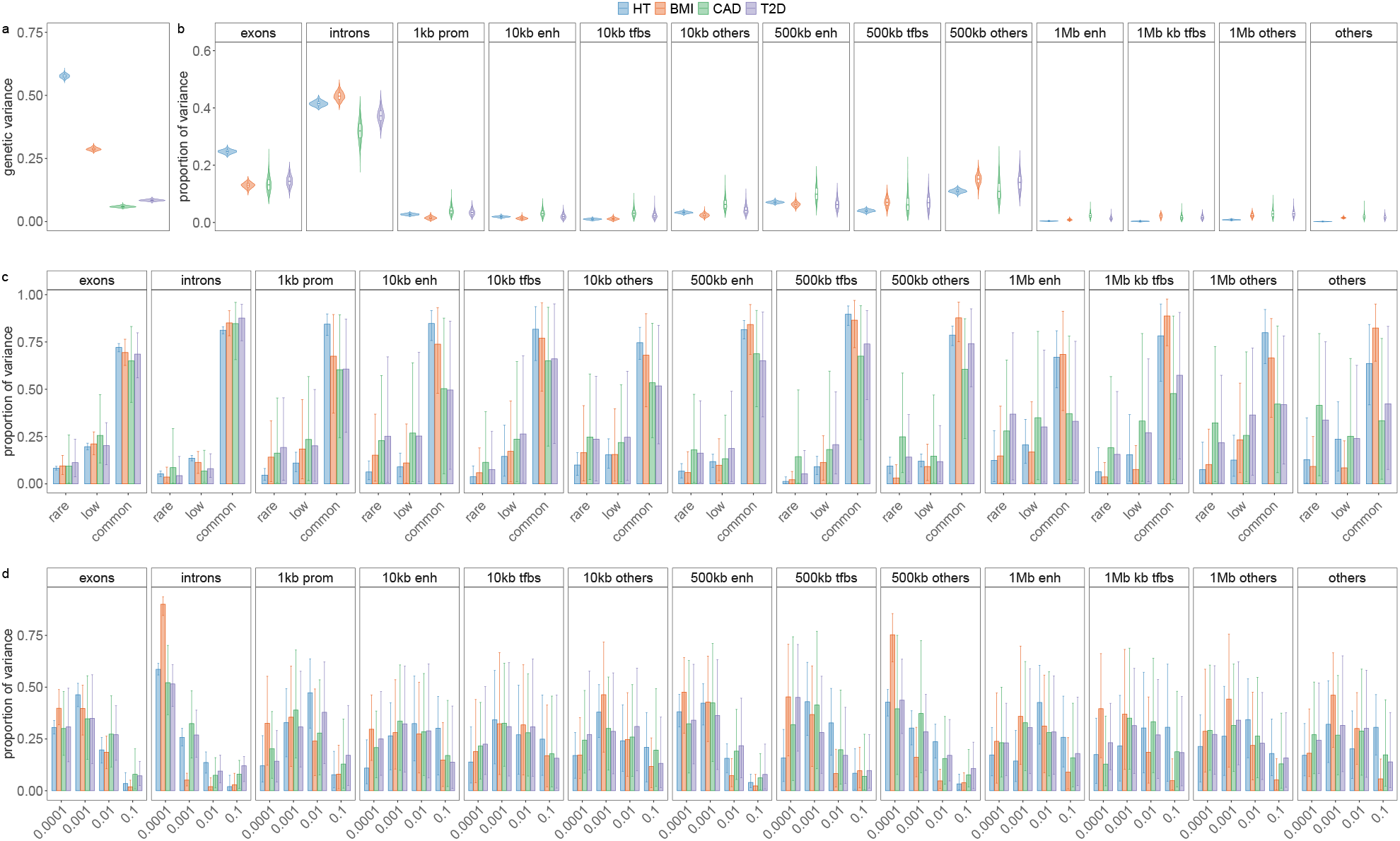
Genetic architecture of height, body-mass-index (BMI), cardiovascular disease (CAD) and type-2-diabetes (T2D). (a) Shows violin plots with boxplots giving the 95% credible intervals for the posterior mean of the phenotypic variance attributable to the SNP markers in each trait. We find that SNPs contribute 57.66% (95%CI 56.09, 59.14) for height, 28.74% (95%CI 27.62, 30.00) for BMI, 5.94% (95%CI 5.30, 6.67) for CAD and 8.45% (95%CI 7.83, 9.18) for T2D. Values are summed over annotation, MAF and LD groups. (b) Violin plot with boxplots giving the 95% credible intervals of the proportion of the total genetic variance attributable to each annotation group. Values are summed over MAF and LD groups. All four traits show the same pattern of annotation-specific genetic variance, with main contributions from intronic regions, exonic regions, and SNPs located 10kb to 500kb upstream of genes to the genetic variance in the population. (c) Bar plots with error bars giving the 95% credible intervals for the proportion of variance of each annotation group that is attributable to each of the four non-zero mixtures for each trait. Values are summed over MAF and LD groups. (d) Bar plots with error bars giving the 95% credible intervals for the proportion of variance of each annotation group that is attributable to each of the three MAF groups for each trait. Values are summed over LD groups. Within each annotation, variation is (c) attributable predominantly to variants with MAF>0.05 and (d) attributable predominantly to small (0.0001) to moderate (0.001) effect sizes variants with little differences across traits, except for BMI which has higher polygenicity compared to height, CAD and T2D.

**Figure S13.**
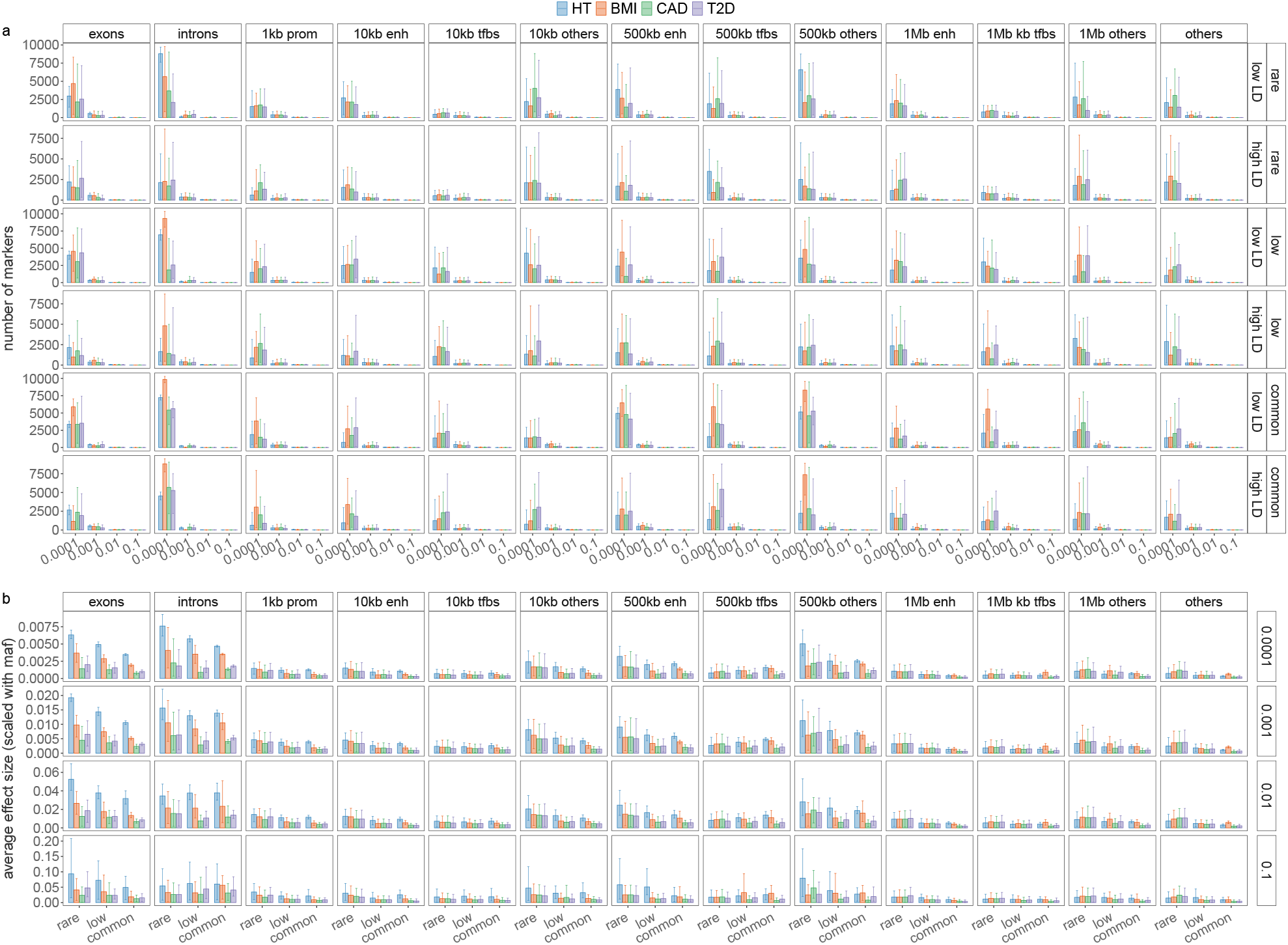
Marker inclusion and effect estimate overview. (a) Barplots of the number of markers entering the model for each mixture group (x-axis), within each MAF-LD group (y-axis facets, with top row MAF and bottom row LD), within each annotation (x-axis facets). Mixture 1 = 0.0001, 2 = 0.001, 3 = 0.01, 4 = 0.1. (b) Boxplots of the posterior distribution of the average effect size of markers in the model for each annotation group, scaling the effects to their frequency and split by mixture.

**Figure S14.**
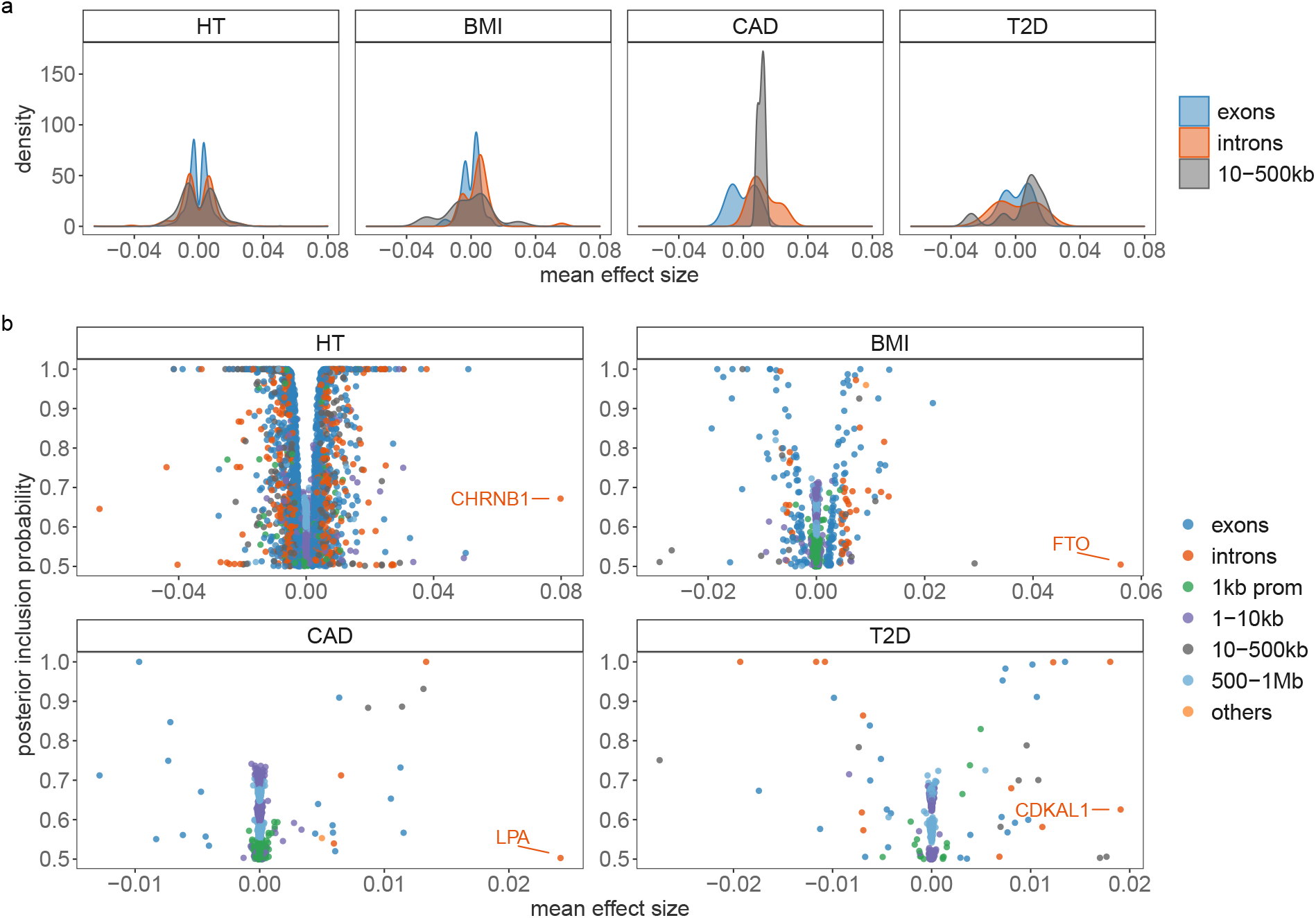
Contribution of SNPs with posterior inclusion probability (PIP) > 0.5 to height, body-mass-index (BMI), cardiovascular disease (CAD) and type-2-diabetes (T2D). (a) Shows the distribution of mean effect sizes for SNPs with PIP > 0.5 attributed to exons, introns and 500kb upstream of genes in each trait. (b) We then plot the relationship between mean effect size and posterior inclusion probability for SNPs with PIP > 0.5 attributed to the annotation groups (exons, introns, SNPs located 1kb, 1-10kb, 10-500kb and 500-1Mb upstream of genes and other un-mapped SNPs). We labelled the closest gene to the SNP with the highest mean effect size in each trait.

**Figure S15.**
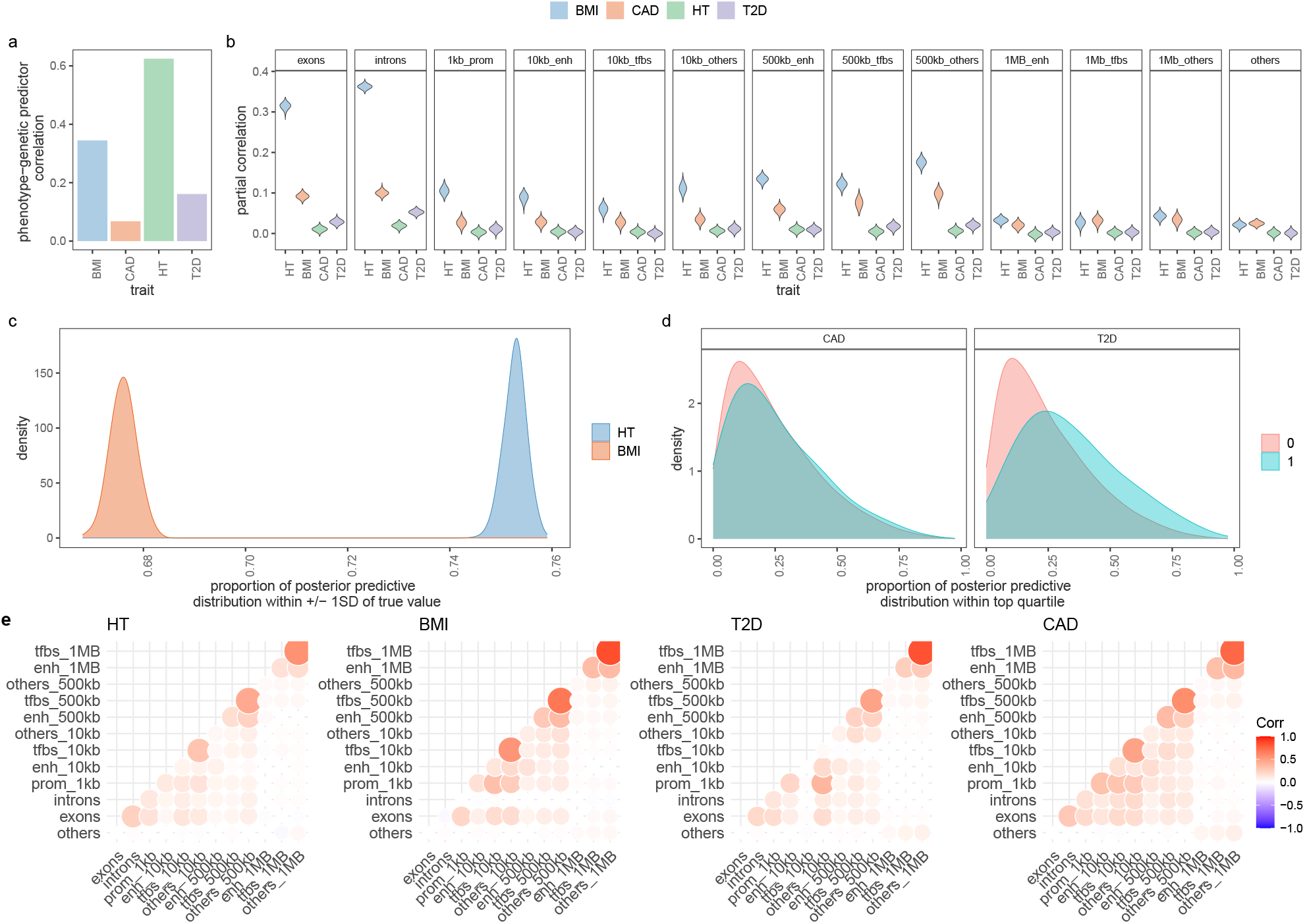
Cross-cohort prediction accuracy and the posterior predictive distribution. (a) Correlation of the posterior mean predictor and height (HT), body mass index (BMI), type-2 diabetes (T2D), and cardiovascular disease (CAD). (b) the partial correlations of the phenotype and genomic predictors specific to different genomic annotations. (c) For height and BMI, we calculate the probability that the distribution of genomic predictors obtained for each individual is within 1 SD of the true phenotypic value. The density of these probabilities is shown. (d) Correlation of genetic predictors obtained across annotation groups.

## Notes

### Competing Interest Statement

The authors have declared no competing interest.

### Author Declarations

Canton Vaud ethics committee.

### Summary of Updates

A large number of additional simulations were conducted to benchmark the method against a number of alternative approaches.

